# Epidemiological, Clinical Characteristics and Risk Factors for Severe Condition and ICU Admission of COVID-19 Patients of Early Stage in China: A review

**DOI:** 10.1101/2021.02.15.21249676

**Authors:** Qinwei Fu, Wenjuan Wu, Yang Liu, Heyin Huang, Peihai Zhang, Xinrong Li, Ying Liu, Lanzhi Zhang, Hui Yang, Xin Zhang, Qinxiu Zhang, Chaolin Huang

**Author notes:** Contributed equally (Qinwei Fu, Wenjuan Wu, Yang Liu, Heyin Huang, Peihai Zhang, Xinrong Li, Ying Liu). **Correspondence to:** 1. Prof. Qinxiu Zhang, School of Medical and Life Sciences, Chengdu University of Traditional Chinese Medicine, Chengdu, 610041, China; Hospital of Chengdu University of Traditional Chinese Medicine, Chengdu, 610072, China., 2.Prof. Chaolin Huang, Jin Yin-tan Hospital, Wuhan, 430022, China. **Funding** None.

## Abstract

**Objective:** Some retrospective studies reporting epidemiological, clinical characteristics of COVID-19 patients of early stage were published. We aim to provide an overview of epidemiological and clinical characteristics of the COVID-19 patients, and identification, treatment of early stage, especially for the patients with poor prognosis.

**Data Sources:** PubMed, CNKI and Google Scholar.

**Study Selection:** We searched for published retrospective studies that described epidemiological and clinical characteristics of confirmed COVID-19 patients in China by April 14^th^, 2020 with search terms. Some studies were excluded according to criteria. Finally, 53 studies were included.

**Data Extraction:** Characteristics of the COVID-19 patients available from included articles were extracted, reorganized and recorded into electronic data forms.

**Data Synthesis:** Characteristics of patients in the included studies were summarized and analyzed for median-interquartile ranges and univariable odds ratio.

**Conclusion:** This study summarized, analyzed and compared epidemiological, clinical characteristics and estimated univariable risk factors among confirmed COVID-19 patients either in former epicenter, in severe condition, with ICU admission or not of early stage. Higher proportions of patients were found to have older age and more comorbidities, typical characteristics on admission and complications either in former epicenter, with severe condition or ICU admission. No evidence showed that patients who were male or had smoking history had higher susceptibility, but they were significant risk factors for severe condition. Some self-implementable traditional Chinese medicine therapies conducted for immunity improvement, control of comorbidities and reduction of some medicine intake. Limited evidence revealed that some characteristics of the disease might be changing with human-to-human transmission, and more research, especially international collaboration, is needed.

**Copyright form disclosure:** The authors have disclosed that they do not have any potential conflicts of interest.

## INTRODUCTION

In early December 2019, some cases of pneumonia with unknown cause, identified and named as SARS-Cov-2 later, occurred in Wuhan City, the capital city of Hubei Province (hereinafter former epicenter), China [1]. Results of gene sequence of the virus collected from infected patients showed a high similarity to virus in bats [2], and through haplotype analyses of 93 complete genomes of SARS-CoV-2, the source was identified as not from Huanan Seafood Market but from some other origin(s) [3]. As of April 26^th^, 2020, a total of 2,804,796 people were confirmed infected with SARS-CoV-2, including 193,710 deaths worldwide [4].

Till April 14^th^, 2020, over 80 retrospective case series had reported epidemiological and clinical characteristics of COVID-19 patients in or outside of former epicenter at different stages of transmission and scale of location in China. Typical signs and symptoms of COVID-19 patients includes fever, dry cough, myalgia and fatigue, which were found similar to severe acute respiratory syndrome (SARS) with high mortality [5]. Clustering onset of the disease and higher susceptibility among older males with comorbidities were reported [6], and evidence from Li et al indicated that human-to-human transmission had occurred since mid-December 2019 [7]. Some studies reported characteristics of the patients outside former epicenter, indicating relatively mild symptoms among them than in Hubei Province [8,9]. In addition, the confirmed patients sometimes assumed no typical symptoms and radiologic abnormalities [10], and more attention should be paid to hospital-related transmission [12].

In this study, we summarized, analyzed and compared confirmed cases from 53 retrospective studies of early stage in China, involving epidemiological characteristics, clinical outcomes, comorbidities, signs and symptoms on admission, complications, radiologic and laboratory findings on admission and treatment. Based on the 53 retrospective studies, more research was done concerning patients in/outside former epicenter, with/without severe condition and with/without ICU admission in terms of epidemiological and clinical characteristics, and risk factors of patients. Our study could be helpful for researchers and medical workers to have a more comprehensive and dynamic view of the characteristics of the disease of early stage in China.

## MATERIALS AND METHODS

### earch strategy and selection criteria

We searched PubMed, CNKI and Google Scholar for published retrospective studies that described epidemiological and clinical characteristics of confirmed COVID-19 patients in China by April 14^th^, 2020. Search terms included “coronavirus”, “COVID” and “2019-nCov” from December 2019 with no language restrictions. We also extensively searched the Internet for other published studies. Case report, research lacking key epidemiological information (e.g., gender or age) or radiological and laboratory findings, report on specific cases (e.g., newborn, familial cluster infections or selected for certain examination), research or data of patients outside China and of suspected were excluded. Finally, 53 studies were included.

### Data analysis

Epidemiological and clinical characteristics were summarized, analyzed and compared in/outside former epicenter, with/without severe condition and with/without ICU admission. Categorical variables summarized as percentages were expressed as median and interquartile ranges (IQR), and differences were compared with the Mann-Whitney U test between patients in/outside former epicenter and with/without severe condition. We also explored univariable risk factors of the patients with severe condition or ICU admission in included studies, and Chi-squared test or Fisher’s exact probability test and one-samples T test were used as appropriate. Statistical analyses were performed by SPSS Version 25.0 Software, and P value <0.05 was considered statistically significant with all tests 2-sided.

## RESULTS

### Overview of studies, epidemiological and clinical characteristics

This study systematically analyzed 53 published retrospective researches since the onset of SARS-CoV-2 as of April 14^th^, 2020 [5,6,8–58]. Among them, 20 studies were in Chinese [20,23,29–31,44–58], and 33 were in English [5–19,21,22,24–28,32–43]. Details of hospital(s), city/cities, province(s), admission period and number of patients in each study are in table S2.

Epidemiological and clinical characteristics of the 53 studies are in figure 1,2 and table S1 and table S3-S36. Proportions of male were higher than 50% in 34 studies, ranging from 30% to 77% [median=53.45%(48.48%-58.03%,IQR)] in 50 studies compared with other three nationwide studies (51.4%, 57.3% and 58.2%). [median=51.6%(34.65%-62%)] of confirmed patients were aging>45-60 years old, and smoking history was reported in 13 studies [median=7% (6.55%-24%, IQR)], but no evidence shows higher susceptibility for current smokers (table S1). Case fatality rates (CFRs) were [median=0.75%(0%-9.53%,IQR)] in total, and [median=18%(9.6%-32.2%,IQR)] of the confirmed patients were in severe condition.

**Figure 1:**
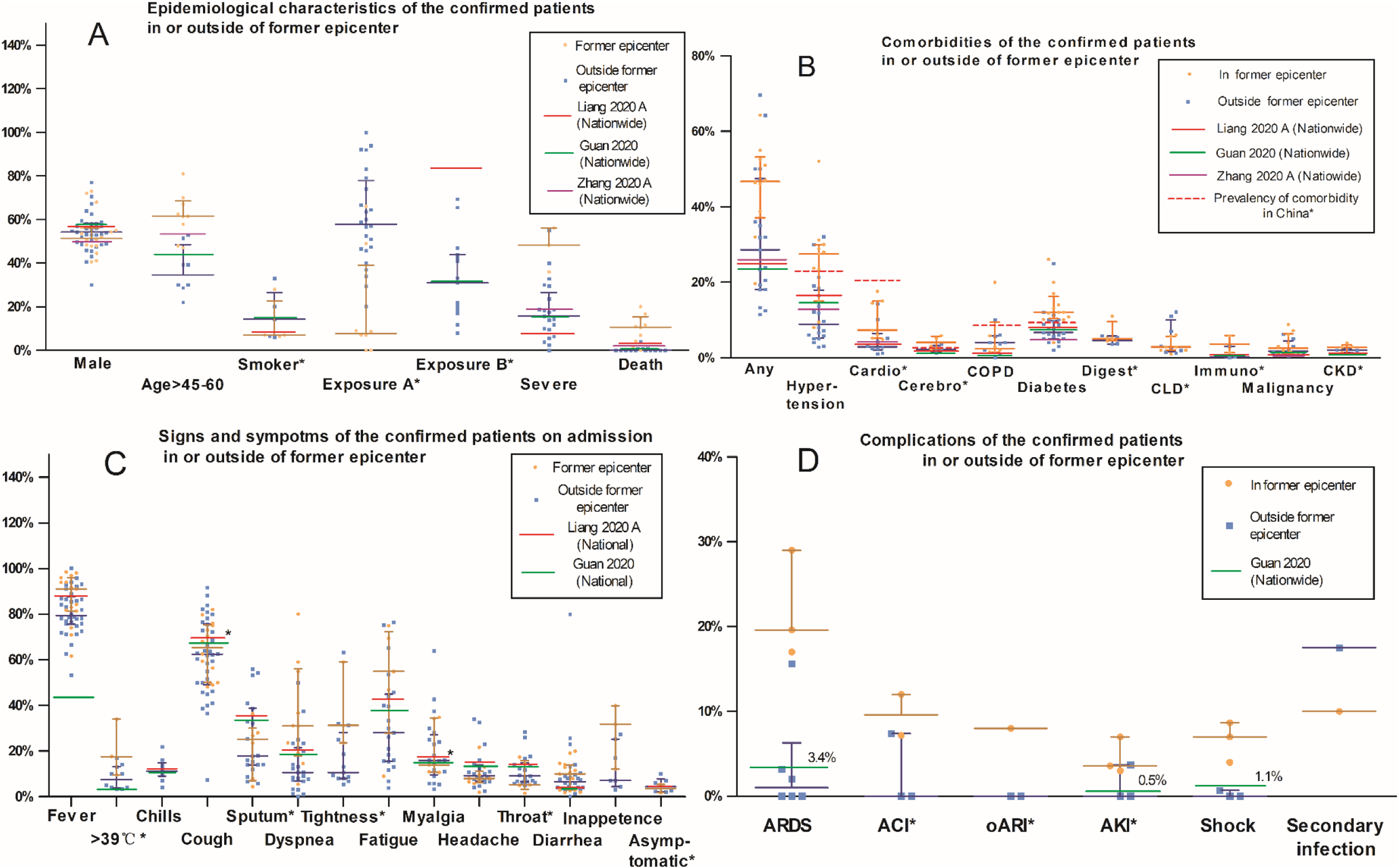
Epidemiological characteristics, clinical outcomes, common comorbidities, signs and symptoms on admission and complications. This figure demonstrates distribution of proportions among epidemiological characteristics, clinical outcomes, common comorbidities, signs and symptoms on admission and complications of confirmed COVID-19 patients in included studies. Each brown or blue point represents one study conducted in or outside of former epicenter respectively, with lines in same color shows median and interquartile range of each subject (e.g. “Male”, “Age>45-60”) and group (i.e. in or outside of former epicenter). Three studies, Liang 2020 A, Guan 2020 and Zhang 2020 A reported epidemiological and clinical characteristics of the patients nation-widely, and they are listed by lines in each subject for reference accordingly. (A) Smoker*: Includes current or(and) former smokers; Exposure A*: Exposure to the Huanan Seafood Market for studies conducted in former epicenter, and exposure to former epicenter for studies outside of former epicenter; Exposure B: Exposure to confirmed patients. (B) Cardio*: Cardiovascular disease; Cerebro*: Cerebrovascular disease; COPD: Chronic obstructive pulmonary disease; Digest*: Digestive disease; CLD*: Chronic liver disease; Immuno*: immunodeficiency or HIV; CKD*: Chronic kidney disease. (C): >39°C*: The highest temperature >39°C during admission; Sputum*: Sputum production; Tightness*: Chest tightness; Throat*: Sore throat; Asymptomatic*: Asymptomatic infection cases. (D): ARDS: Acute Respiratory Distress Syndrome; ACI*: Acute cardiac injury; oARI*: Other acute respiratory injury; AKI*: Acute kidney injury.

**Figure 2:**
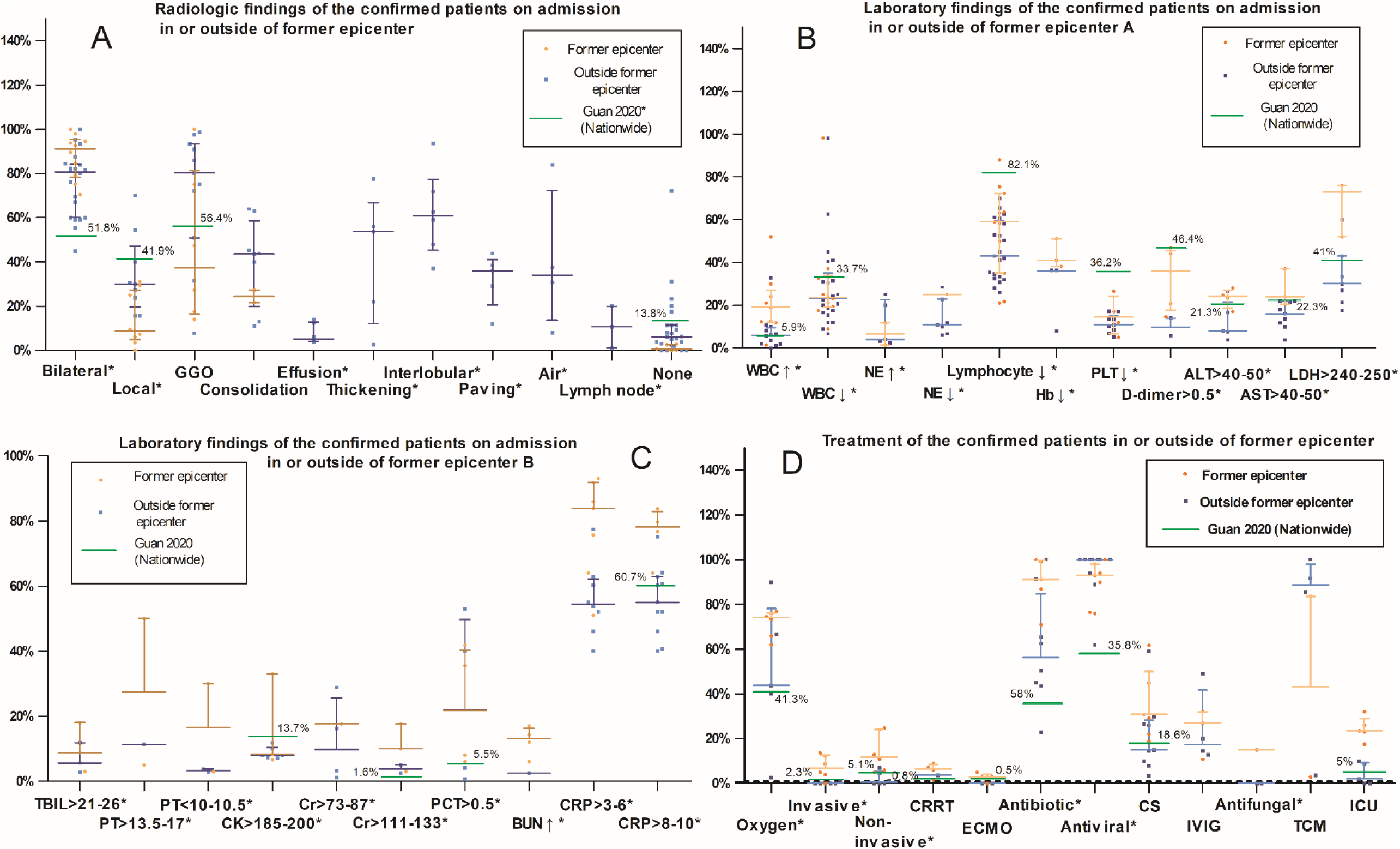
Major radiologic, laboratory findings on admission and treatments. This figure demonstrates distribution of proportions among major radiologic, laboratory findings on admission and treatments of confirmed COVID-19 patients in included studies. Each brown or blue point represents one study conducted in or outside of former epicenter respectively, with lines in same color shows median and interquartile range of each subject (e.g. “Bilateral*”, “Local*”) and group (i.e. in or outside of former epicenter). Three studies, Liang 2020 A, Guan 2020 and Zhang 2020 A reported epidemiological and clinical characteristics of the patients nation-widely, and they are listed by lines in each subject for reference accordingly. (A): Bilateral*: Bilateral involvement; Local*: Local patchy opacity; GGO: Ground glass opacity; Effusion*: Pleural effusion; Thickening*: Pleural thickening; Interlobular*: Interlobular septal thickening; Paving*: Crazy-paving pattern; Air*: Air bronchogram sign; Lymph node*: Lymph node enlargement. (B): WBC ↑*: White blood cell count > 9.5-10 × 10^9^/L; WBC ↓*: White blood cell count < 3.5-4 × 10^9^/L; NE↑*: Neutrophil count >5.8-7 × 10^9^/L; NE↓*: Neutrophil count < 1.8-2 × 10^9^/L; Lymphocyte↓*: Lymphocyte count <0.8-1.5 × 10^9^/L; Hb↓*: Hemoglobin < 110-130g/L; PLT↓*: Platelet count <100-150 × 10^9^/L; D-dimer > 0.5*: D-dimer > 0.5mg/L; ALT > 40-50*: Alanine aminotransferase > 40-50U/L; AST > 40-50*: Aspartate aminotransferase > 40-50U/L; LDH > 240-250*: Lactate dehydrogenase > 240-250U/L. (C): TBIL >21-26*: Total bilirubin >21-26 mmol/L; PT >13.5-17: Prothrombin time >13.5-17s; PT < 10-10.5*: Prothrombin time <10-10.5s; CK >185-200*: Creatine kinase > 185-200U/L; Cr > 73-87*: Serum creatinine > 73-87μmol/L; Cr > 111-133*: Serum creatinine > 111-133μmol/L; PCT > 0.5*: Procalcitonin > 0.5ng/mL; BUN↑*: Blood urea nitrogen > 7.6-9.5mmol/L; CRP > 3-6*: C-reactive protein > 3-6mg/L; CRP > 8-10*: C-reactive protein > 8-10mg/L. (D): Oxygen*: Oxygen therapy; Invasive*: Invasive mechanical ventilation; Non-invasive*: Non-invasive mechanical ventilation; CRRT: Continuous renal replacement therapy; ECMO: Extracorporeal membrane oxygenation; Antibiotic*: Antibiotic treatment; Antiviral*: Antiviral treatment; CS: Corticosteroid therapy; IVIG: Intravenous immunoglobulin therapy; Antifungal*: Antifungal treatment; TCM: Traditional Chinese medicine; ICU: Intensive care unit care.

Results shows that [median=48%(31.08%-58.08%,IQR)] of the patients had at least one comorbidity, and the top 5 comorbidities were hypertension [median=13.7%(6.7%-27.5%,IQR)], diabetes [median=9.6%(5.86%-12%,IQR)], cardiovascular disease [median=5.2%(2.68%-12.25%,IQR)], digestive disease [median=5.1%(3.91%-5.77%,IQR)] and cerebrovascular disease [median=2.8%(2.03%-4.73%,IQR)] (table S1). As for signs and symptoms, the most common one was fever [median=83.15%(76.13%-92.15%,IQR)], followed by cough [median=62.6%(50%-75.38%,IQR)], fatigue [median=31.7%(15.68%-53.93%,IQR)], sputum production [median=18.1%(13%-34.69%,IQR)], chest tightness, myalgia, inappetence, dyspnea, chills and others (table S1). Most complications occurred in about less than 10% of the patients (acute cardiac injury, other acute respiratory injury, acute kidney injury and shock) in addition to ARDS [median=3.2%(0%-18.3%,IQR)] and secondary infection [13.75%(10%-17.5%,IQR)].

Bilateral involvement [median=81.85%(68.81%-92.85%,IQR)], ground glass opacity [median=75%(29.37%-92.15%,IQR)], interlobular septal thickening [median=60.85%(45.25%-77.33%,IQR)], pleural thickening [median=53.8%(12.3%-66.8%,IQR)] and consolidation [median=41.8%(19.45%-49.73%,IQR)] were most commonly radiological findings tested for the patients on admission. As for laboratory findings, they were C-reactive protein > 3-6 mg/L [median=62.86%(52%-83.9%,IQR)], C-reactive protein > 8-10 mg/L [median=60.7%(52%-75.2%,IQR)], lymphocyte count < 0.8-1.5×10^9^/L [median=50.2%(33.6%-62.45%,IQR)], lactate dehydrogenase > 240-250 U/L [median=38.15%(25.56%-63.25%,IQR)], hemoglobin < 110-130 g/L, white blood cell count < 4× 10^9^/L, D-dimer > 0.5 mg/L and procalcitonin > 0.5 ng/mL. Details of other radiological, laboratory findings and treatments are in table S1.

### Epidemiological and clinical characteristics of the patients in vs. outside the former epicenter

In our study, 15 studies reported characteristics of COVID-19 patients in former epicenter [5,6,10,12–16,22,24,28,44,45,47,53], 30 in one city or province outside former epicenter [7–9,17–19,26,27,29–31,33,34,36–43,46,49–52 54–58] and five in more than one city, province [20,21,25,35,48] or nationwide (in 30/31 provinces) [11,23,32]. Compared with the patients of studies outside former epicenter, patients in former epicenter were older (age>45-60, y) [median=61.5%(52.95%-68.5%,IQR)] vs [median=34.65%(29.03%-48.53%,IQR)] and had higher proportions of severe cases [median=48.3%(36%-56.2%)] vs. [median=15.7%(7.95%-26.55%,IQR)], CFRs [median=10.7%(3.68%-15.43%,IQR)] vs. [median=0%(0%-0.73%,IQR)] and comorbidities [median=46.75%(37.1%-53.2%,IQR)] vs. [median=28.6%(18.1%-47.5%,IQR)] significantly (P<0.05), including hypertension, cardiovascular disease and diabetes (table S1). Higher proportions of the patients in former epicenter had apparent signs and symptoms, including fever, highest temperature>39°C, dyspnea (p<0.01), chest tightness, fatigue and inappetence, but fewer patients had headache and in asymptomatic infection in center (table S1). Higher proportions of complications, including ARDS (p<0.05), acute cardiac injury, other acute respiratory injury, acute kidney injury and shock, were reported in the patients in former epicenter. On admission, most radiological and laboratory abnormalities were found in more patients in former epicenter, including significantly (p<0.05) higher proportions of bilateral involvement (chest radiological examination), white blood cell count > 10 × 10^9^/L, lactate dehydrogenase > 240-250 U/L, C-reactive protein > 3-6 mg/L and C-reactive protein > 8-10 mg/L (Table S1). Invasive/non-invasive mechanical ventilation and ICU care were applied to higher proportions of the patients in former epicenter significantly (p<0.05).

### Characteristics and univariable risk factors among patients in severe condition/ ICU admission

Epidemiological and clinical characteristics of the COVID-19 patients were furtherly divided into ICU/ non-ICU groups in two studies [5, 12], and into severe/ non-severe groups in 20 studies [10,11,15,17,21,24–26,29,33,35,37–39,43,44,49,51,53,55] (table S37-S41).

Proportions of male, age>45-60 and comorbidities including hypertension, diabetes, cardiovascular disease and COPD were significantly higher in patients with severe condition (p<0.05, table S37), together with some signs, symptoms, radiological and laboratory findings on admission including dyspnea, lactate dehydrogenase > 225-250 U/L, bilateral involvement (radiological) and pleural effusion (radiological).

ICU cares were required in [median=13.75%(1.5%-24.15%,IQR)] of the patients, and risk factors (p<0.05) of them included hospitalized patients, with some comorbidities including hypertension, cerebrovascular disease and diabetes, with some clinical characteristics including cough, dyspnea, sore throat, dizziness, respiratory rate >24 breaths/ min, white blood cell count > 10 × 10^9^/L, hypersensitive troponin I >28 pg/mL (99th percentile) and procalcitonin ≥ 0.5 ng/mL (table S39).

Details of univariable risk factors of the patients with severe condition/ICU admission are in table S39, and characteristics among patients with severe condition/ICU admission are in table S37.

## DISCUSSION

Results of our study showed that over half of the confirmed COVID-19 patients were middle-aged and older male, with more cases of older people in former epicenter. Proportions of the male patients in our study were higher than the male population in China (t=2.36, p=0.022) as of 2019 (51.08%) and in Wuhan (t=1.03, p=0.32) as of 2017 (51%) [59, 60]. 14 studies reported 0%-48% of medical staff infection with relative milder prognosis (p<0.05). As of Feb 11^th^, 2020, a total of 1716 (3.8%) medical staff were infected with SARS-CoV-2, with death rate at 0.4%, six deaths in China [23]. One study was about two death cases of old male with a long smoking history but no previous chronic underlying disease [6]. [median=7%(6.55%-24%,IQR)] of confirmed patients had smoking history, lower than the proportion of current smokers (>15 y, 27.7%) in China by 2015 markedly (t=−4.35, p<0.002) [26]. However, higher percentages of former smokers were reported in patients with severe condition than non-severe [median=6.9%(5.2%-25%,IQR)] vs. [median=3.7%(1.3%-17%,IQR)], and it was also connected with severe condition significantly (p>0.05). This may be because of significantly higher ACE2 gene expression in lungs of former smokers, which is receptor for SARS-CoV-2 [62]. CFRs were [median=10.7%(3.68%-15.43%,IQR)] in former epicenter and [median=0%(0%-0.73%,IQR)] outside according to included studies, and significantly higher among patients with severe condition [median=6.5%(1.3%-17%,IQR)] vs. [median=0%(0%-0.9%,IQR)] and ICU admission (16.7%-38% vs 0%-4%). Epidemiological characteristics including male, age ≥ 45-60 and former smoker and hospitalized patients had significant connections with severe condition or ICU admission (p<0.05).

Nearly all comorbidities were reported in higher proportions among the patients in former epicenter, severe condition or with ICU admission than not, and some of them were also significant risk factors of severe condition or ICU admission among COVID-19 patients (P>0.05) as a result of relatively poor immunity [6], including hypertension, cardiovascular disease, cerebrovascular disease, cardio-cerebrovascular disease, diabetes, COPD, malignancy, immunosuppression. However, no evidence shows that proportions of the patients with hypertension (t=−3.12, p<0.01), cardiovascular disease (t=−10.68, p<0.01), cerebrovascular disease (t=1.75, p=0.124), diabetes (t=0.72, p=0.477) and COPD (t=−3.07, p<0.01) in most studies were significantly higher than the national prevalency of China [63–67].

However, we noticed that higher proportions of comorbidities, such as immunodeficiency HIV and malignancy, were reported in some non-ICU and non-severe groups, indicating poor immunity. Result of minimally invasive pathological examination by Xu et al also showed that severe immune injury in COVID-19 patients could be caused by overactivation of T cells, partially, manifested by increase of Th17 and high cytotoxicity of CD8 T cells [68]. Pulmonary and systemic inflammatory responses associated with coronavirus are triggered by the innate immune system mostly when viruses are recognized, and this type of immune response can also inhibit virus replication, promote virus clearance, induce tissue repair, and trigger a prolonged adaptive immune response against the viruses [69]. As a result, further research is needed on how to neutralize harmful immune response and even make it a therapy for COVID-19 patients especially with severe condition or ICU admission.

With increasing human-to-human transmission of the virus, more confirmed COVID-19 patients were reported with no typical signs, symptoms, and radiologic, laboratory findings on admission and with milder severity especially for studies outside Hubei. This is most likely because of more patients were diagnosed earlier than those in Hubei at the early stages of the SARS-CoV-2 outbreak. However, we cannot rule out that some of the characteristics and severity of the disease may change with human-to-human transmission. Research of Bai et al and Hu et al indicated that patients under asymptomatic infection may have infectivity of the virus and even become a “super infector” [70; 71]. In addition, there were reports of several cases of positive results or false-negative results of RT-PCR test in diagnosis after discharge from hospital upon treatment for even 5 to 13 days in China and Canada [72; 73; 74].

Laboratory findings showed that alanine aminotransferase, aspartate aminotransferase, total bilirubin, creatinine, creatine kinase, lactate dehydrogenase and hypersensitive troponin I of some the COVID-19 patients were beyond normal range, especially of significant relevance for patients with severe conditions or ICU admission, indicating liver, heart and kidney injury caused by SARS-CoV-2 [75–77].

Lymphocytes count of more than half of the patients on admission were below the normal range in our study [median=50.2%(33.6%-62.45%,IQR)], especially in severe groups [median=82.1%(71.8%-88.9%,IQR)], which probably had significant connections with disease severity and mortality [78]. Under the requirement of avoiding unnecessary outings and home isolation, some self-implementable traditional Chinese medicine therapies helpful for improving immunity are recommended, including moxibustion, acupressure and some traditional exercises such as the *Tai Chi Chuan* exercise [79–82]. In addition, they can relieve the burden of some comorbidities such as cancer pain, diabetic peripheral neuropathy and asthma [80,83,84] by decreasing duration, frequency or severity. As a result, they can also reduce medication intake, especially those that may have considerable addiction and adverse events (e.g. opioid, lidocaine medicated plaster and oral corticosteroids) [85,86].

Our study had several limitations. Firstly, there might be some repetition among data of patients from the 53 researches included in our study because of the overlap in range of time and location, and a table of some detailed information of the included studies are in table S2. Secondly is the relatively small number of researches included in our study for risk factor analysis among patients with/without ICU admission, so more researches are needed on epidemiological and clinical characteristics of COVID-19 patients with different ICU admission groups. Finally, results of our study are based on published researches, while many issues of SARS-Cov-2 are still uncertain, so there may exist bias in our study which justifies a modest interpretation of our results.

## CONCLUSION

This study summarized, analyzed and compared epidemiological, clinical characteristics and estimated univariable risk factors among confirmed COVID-19 patients in/outside former epicenter, with/without severe condition, with/without ICU admission of early stage in China. Higher proportions of patients were found to have older age and more comorbidities, typical characteristics on admission and complications either in former epicenter, with severe condition or ICU admission. No evidence showed that patients who were male or had smoking history had higher susceptibility, but they were significant risk factors for severe condition. Some self-implementable TCM therapies should be recommended for immunity improvement, control of comorbidities and reduction of some medication intake. Limited evidence revealed that some characteristics of the disease might be changing with human-to-human transmission, and more research, especially international collaboration, is needed.

## Supporting information

not applicable

## Data Availability

Not applicable.

## ACKNOWLEDGEMENTS

We thank all the patients and authors involved in the researches included in our study.

## CONTRIBUTORS

QF and QZ put forwarded the study design. YaL, HY, LZ, and XL contributed to the study searching and data collection. QF, CH, QZ, HH, PZ, XL and XZ contributed to data summarization, analyzation and comparison. QF, WW and YaL contributed to statistical analysis. WW, QF, and YiL contributed to the design and drawing of tables and figures. QF, QZ, HH, WW, YiL and CH contributed to the writing of the manuscript. All reviewed and approved the final version of the manuscript.

## REFERENCES

1 Lu H, Stratton CW, Tang YW. Outbreak of pneumonia of unknown etiology in Wuhan China: the mystery and the miracle. J Med Virol 2020,; published online Jan 16. doi:10.1002/jmv.25678.

2 Ji W, Wang W, Zhao X, Zai J, Li X. Homologous recombination within the spike glycoprotein of the newly identified coronavirus may boost cross-species transmission from snake to human. J Med Virol 2020; published online Jan 22. doi:10.1002/jmv.25682.

3 Yu WB, Tang GD, Zhang, L, Corlett RT. Decoding evolution and transmissions of novel pneumonia coronavirus using the whole genomic data. ChinaXiv 2020; published online Feb 21. doi: 10.12074/202002.00033.

4 World Health Organization. Coronavirus disease 2019 (COVID-19). Situation report-97. Apr 26, 2020. https://www.who.int/docs/default-source/coronaviruse/situation-reports/20200426-sitrep-97-Covid-19.pdf?sfvrsn=d1c3e800_6.

5 Huang C, Wang Y, Li X, et al. Clinical features of patients infected with 2019 novel coronavirus in Wuhan, China. Lancet 2020; 395(10223):497–506.

6 Chen N, Zhou M, Dong X, et al. Epidemiological and clinical characteristics of 99 cases of 2019 novel coronavirus pneumonia in Wuhan, China: a descriptive study. Lancet 2020; 395(10223):507– 513.

7 Li Q, Guan X, Wu P, et al. Early Transmission Dynamics in Wuhan, China, of Novel Coronavirus-Infected Pneumonia. N Engl J Med 2020; published online Jan 29. doi:10.1056/NEJMoa2001316.

8 Xu XW, Wu XX, Jiang XG, et al. Clinical findings in a group of patients infected with the 2019 novel coronavirus (SARS-Cov-2) outside of Wuhan, China: retrospective case series. BMJ 2020; published online Feb 19. doi:10.1136/bmj.m606.

9 Chang D, Lin M, Wei L, et al. Epidemiologic and Clinical Characteristics of Novel Coronavirus Infections Involving 13 Patients Outside Wuhan, China. JAMA 2020; published online Feb 7. doi:10.1001/jama.2020.1623.

10 Mao L, Jin H, Wang M, et al. Neurologic Manifestations of Hospitalized Patients with Coronavirus Disease 2019 in Wuhan, China. JAMA Neurol 2020; published online Apr 10. doi:10.1001/jamaneurol.2020.1127.

11 Guan WJ, Ni ZY, Hu Y, et al. Clinical Characteristics of Coronavirus Disease 2019 in China. N Engl J Med 2020; published online Feb 28. doi:10.1056/NEJMoa2002032.

12 Wang D, Hu B, Hu C, et al. Clinical Characteristics of 138 Hospitalized Patients With 2019 Novel Coronavirus-Infected Pneumonia in Wuhan, China. JAMA 2020; published online Feb 7. doi:10.1001/jama.2020.1585.

13 Chen L, Liu HG, Liu W, et al. Analysis of clinical features of 29 patients with 2019 novel coronavirus pneumonia. Zhonghua Jie He He Hu Xi Za Zhi 2020; published online Feb 06. doi:10.3760/cma.j.issn.1001-0939.2020.0005. (Chinese).

14 Huang Y, Tu M, Wang S, et al. Clinical characteristics of laboratory confirmed positive cases of SARS-CoV-2 infection in Wuhan, China: A retrospective single center analysis. Travel Med Infect Dis 2020; published online 2020 Feb 27. doi:10.1016/j.tmaid.2020.101606.

15 Zhang JJ, Dong X, Cao YY, et al. Clinical characteristics of 140 patients infected with SARS-CoV-2 in Wuhan, China. Allergy 2020; published online Feb 19. doi:10.1111/all.14238.

16 Liu K, Fang YY, Deng Y, et al. Clinical characteristics of novel coronavirus cases in tertiary hospitals in Hubei Province. Chin Med J (Engl) 2020; published online Feb 7. doi:10.1097/CM9.0000000000000744.

17 Tian S, Hu N, Lou J, et al. Characteristics of COVID-19 infection in Beijing. J Infect. 2020; published online Feb 27. doi:10.1016/j.jinf.2020.02.018.

18 Wu J, Liu J, Zhao X, et al. Clinical Characteristics of Imported Cases of COVID-19 in Jiangsu Province: A Multicenter Descriptive Study. Clin Infect Dis 2020; published online Feb 29. doi:10.1093/cid/ciaa199.

19 Yang W, Cao Q, Qin L, et al. Clinical characteristics and imaging manifestations of the 2019 novel coronavirus disease (COVID-19): A multi-center study in Wenzhou city, Zhejiang, China. J Infect 2020; published online Feb 26. doi:10.1016/j.jinf.2020.02.016.

20 Liu C, Jiang ZC, Shao CX, et al. Zhonghua Gan Zang Bing Za Zhi 2020; 28(2):148–152. doi:10.3760/cma.j.issn.1007-3418.2020.02.003 (Chinese).

21 Xu YH, Dong JH, An WM, et al. Clinical and computed tomographic imaging features of novel coronavirus pneumonia caused by SARS-CoV-2. J Infect 2020; published online Feb 25. doi:10.1016/j.jinf.2020.02.017.

22 Cao J, Tu WJ, Cheng W, et al. Clinical Features and Short-term Outcomes of 102 Patients with Corona Virus Disease 2019 in Wuhan, China. Clin Infect Dis 2020; published online Apr 2. doi: 2020;ciaa243. doi:10.1093/cid/ciaa243.

23 The Novel Coronavirus Pneumonia Emergency Response Epidemiology Team. The Epidemiological Characteristics of an Outbreak of 2019 Novel Coronavirus Diseases (COVID-19) — China, 2020. China CDC Weekly 2020; 2(8): 113–122.

24 Li YK, Peng S, Li LQ, et al. Clinical and Transmission Characteristics of Covid-19 - A Retrospective Study of 25 Cases from a Single Thoracic Surgery Department. Curr Med Sci 2020; published online Mar 30. doi: 10.1007/s11596-020-2176-2. doi:10.1007/s11596-020-2176-2

25 Li K, Wu J, Wu F, et al. The Clinical and Chest CT Features Associated with Severe and Critical COVID-19 Pneumonia. Invest Radiol 2020; published online Feb 29. doi: 10.1097/RLI.0000000000000672.

26 Zhao W, Zhong Z, Xie X, et al. Relation Between Chest CT Findings and Clinical Conditions of Coronavirus Disease (COVID-19) Pneumonia: A Multicenter Study. AJR Am J Roentgenol 2020; published online Mar 3. doi:10.2214/AJR.20.22976.

27 Xu T, Chen C, Zhu Z, et al. Clinical features and dynamics of viral load in imported and non-imported patients with COVID-19. Int J Infect Dis 2020; published online Mar 13. doi:10.1016/j.ijid.2020.03.022.

28 Wang K, Kang S, Tian R, et al. Imaging manifestations and diagnostic value of chest CT of coronavirus disease 2019 (COVID-19) in the Xiaogan area. Clin Radiol 2020; 75(5):341–347.

29 Tang GX, Li CH, et al. Clinical and CT findings of coronavirus disease 2019. Chinese Journal of Respiratory and Critical Care Medicine 2020; 19(02):161–165. (Chinese).

30 Huang XQ, Nie LH, Li FM, et al. Analysis of Chinese Medical Characteristics of 35 Patients with Novel Coronavirus Pneumonia. Journal of Emergency in Traditional Chinese Medicine 2020; 29(03):381–383+398. (Chinese).

31 Liu SD, Jiang XG, Ning HY, et al. Analysis of clinical features of 42 cases of new coronavirus pneumonia. Shanghai Journal of Preventive Medicine 2020; published online Apr 19. doi: 10.19428/j.cnki.sjpm.2020.20092. (Chinese).

32 Liang WH, Guan WJ, Li CC, et al. Clinical characteristics and outcomes of hospitalised patients with COVID-19 treated in Hubei (epicenter) and outside Hubei (non-epicenter): A Nationwide Analysis of China. Eur Respir J. 2020; published online Apr 8. doi:10.1183/13993003.00562-2020.

33 Zheng F, Tang W, Li H, Huang YX, Xie YL, Zhou ZG. Clinical characteristics of 161 cases of corona virus disease 2019 (COVID-19) in Changsha. Eur Rev Med Pharmacol Sci 2020; 14 24(6):3404–3410.

34 Dai H, Zhang X, Xia J, et al. High-resolution Chest CT Features and Clinical Characteristics of Patients Infected with COVID-19 in Jiangsu, China. Int J Infect Dis 2020; published online Apr 6. doi:10.1016/j.ijid.2020.04.003.

35 Feng Y, Ling Y, Bai T, et al. COVID-19 with Different Severity: A Multi-center Study of Clinical Features. Am J Respir Crit Care Med 2020; published online Apr 10. doi:10.1164/rccm.202002-0445OC.

36 Peng L, Liu KY, Xue F, et al. Improved Early Recognition of Coronavirus Disease-2019 (COVID-19): Single-Center Data from a Shanghai Screening Hospital. Arch Iran Med 2020; 23(4):272–276. doi:10.34172/aim.2020.10.

37 Lo IL, Lio CF, Cheong HH, et al. Evaluation of SARS-CoV-2 RNA shedding in clinical specimens and clinical characteristics of 10 patients with COVID-19 in Macau. Int J Biol Sci 2020; 16(10):1698–1707. doi:10.7150/ijbs.45357.

38 Wan S, Xiang Y, Fang W, et al. Clinical features and treatment of COVID-19 patients in northeast Chongqing. J Med Virol 2020; published online Mar 21. doi:10.1002/jmv.25783. doi:10.1002/jmv.25783.

39 Qian GQ, Yang NB, Ding F, et al. Epidemiologic and Clinical Characteristics of 91 Hospitalized Patients with COVID-19 in Zhejiang, China: A retrospective, multi-centre case series. QJM 2020; published online Mar 17. doi:10.1093/qjmed/hcaa089

40 Chen J, Qi T, Liu L, et al. Clinical progression of patients with COVID-19 in Shanghai, China. J Infect 2020; 80(5):e1–e6.

41 Xu X, Yu C, Qu J, et al. Imaging and clinical features of patients with 2019 novel coronavirus SARS-CoV-2. Eur J Nucl Med Mol Imaging 2020; 47(5):1275–1280.

42 Wu J, Wu X, Zeng W, et al. Chest CT Findings in Patients With Coronavirus Disease 2019 and Its Relationship With Clinical Features. Invest Radiol 2020; 55(5):257–261.

43 Liu KC, Xu P, Lv WF, et al. CT manifestations of coronavirus disease-2019: A retrospective analysis of 73 cases by disease severity. Eur J Radiol 2020; 126:108941.

44 Bin YF, Ji P, Liang XD, et al. Clinical characteristics of 55 hospitalized patients with COVID-19 in Wuhan, China. Journal of Guangxi Medical University 2020; 37(02):338–342. (Chinese).

45 Chen ZY, Chen ZY, Zhang XH, et al. Clinical manifestations and CT characteristics of corona virus disease 2019 (COVID-19). Radiologic Practice 2020; 35(03):286–290. (Chinese).

46 Diao KY, Han PL, Pang T, et al. The features and longitudinal changes on high-resolution computed tomography for patients with coronavirus disease 2019. Chinese Journal of Respiratory and Critical Care Medicine 2020; 19(02):166–171. (Chinese).

47 Din Y, Huang ZF, Zhao SC, et al. Clinical and imaging characteristics of corona virus disease 2019 (COVID-19. Radiologic Practice 2020; 35(03):281–285. (Chinese).

48 Gao T, Xu YL, He XP, et al. Epidemiological and clinical characteristics of 40 patients with coronavirus disease 2019 outside Hube. Chinese Journal of Respiratory and Critical Care Medicine 2020; 02:148–153. (Chinese).

49 Li YL, Shan NB, Sun W, et al. Comparative Study for Clinical Features between COVID-19 Patients with Conventional Type and Heavy/Critical Type. Practical Journal of Cardiac Cerebral Pneumal and Vascular Disease 2020; 28(03):14–19. (Chinese).

50 Liang YL, Yue F, Song L. Epidemiological and Clinical Characteristics of 28 Confirmed Cases of Novel Coronavirus Pneumonia. Clinical research 2020; (04):1–3. (Chinese).

51 Lv R, Feng YY, Zhang YN, et al. CT features and analysis of 17 cases of COVID-19. International Journal of Medical Radiology 2020; 02:131–134. (Chinese).

52 Mao Y, Cui X, Jiang ZC, et al. COVID-19 in Ankang District, Shaanxi: analysis of 24 cases of TCM syndromes and clinical characteristics. Shaanxi Journal of Traditional Chinese Medicine 2020; 04:427–428+517. (Chinese).

53 Wang R, Xie LL, D. P, et al. Clinical characteristics of 96 hospitalized patients with coronavirus disease 2019. Chinese Journal of Respiratory and Critical Care Medicine 2020; 02:144–147. (Chinese).

54 Wang SB, Peng W, Yu L, et al. Clinical analysis of 17 cases of coronavirus disease 2019. Zhejiang Medical Journal 2020; 04:365–367. (Chinese).

55 Xiang TX, Liu JM, Xu F, et al. Analysis of clinical characteristics of 49 patients with coronavirus disease 2019 in Jiangxi. Chinese Journal of Respiratory and Critical Care Medicine 2020; 02:154–160. (Chinese).

56 Xu H, Li SH, Zhao CC, et al. Clinical and imaging features of 32 cases of corona virus disease 2019. Journal of Bengbu Medical College 2020; 02:150–153+155. (Chinese).

57 Yu SM, Wang ZX, Qin EQ, et al. Clinical characteristics of 25 patients with COVID-19. Chinese Journal of Integrated Traditional and Western Medicine 2020; 03:369–370. (Chinese).

58 Yu JX, Zhang XM, Huang JS. Epidemiological and clinical characteristics of 87 cases of coronavirus disease 2019 in Hangzhou. Zhejiang Medical Journal 2020; 04:318–320+324. (Chinese).

59 National Bureau of Statistics of China. Gender ratio of Chinese population as of 2018. 2018. http://data.stats.gov.cn/easyquery.htm?cn=C01&zb=A0301&sj=2019 (Chinese).

60 Hu Z, Jin Y, Liu J, et al. POPULATION, EMPLOYMENT AND WAGE. In: Wuhan Municipal Bureau of Statistics, ed. Wuhan statistical yearbook, 2018. China statistics press. Beijing: Aug 2018: 38. PDF: http://117.136.191.144/cache/tjj.wuhan.gov.cn/Attachment/201901/201901041649116279.pdf?ich_args2=121-12104410022478_bfef43f0a1290cad7d2961da8cafcfab_10001002_9c896c2fdecbf0d19032518939a83798_8e1152749827eec2f887427ab963d394 (Chinese).

61 Hu SS, Gao RL, Wang Z, et, al OUTLINE. In: National Center for Cardiovascular Diseases, China, ed. REPORT ON CARDIOVASCULAR DISEASES IN CHINA 2018. Encyclopedia of China Publishing House. Beijing: Jan 2019: 3. PDF: http://117.128.6.21/cache/nccd.org.cn/Sites/Uploaded/File/2019/8/14-%E4%B8%AD%E5%9B%BD%E5%BF%83%E8%A1%80%E7%AE%A1%E7%97%85%E6%8A%A5%E5%91%8A2018%EF%BC%88%E4%B8%AD%E6%96%87%E7%89%88%EF%BC%89.pdf?ich_args2=462-11095509026575_6043db1fec4c58b2fce944dd03e44b67_10001002_9c896c2fdfcaf0d6973b518939a83798_2e40ada2aadd6db8d534753403e6cc64 (Chinese).

62 Cai GS. Bulk and single-cell transcriptomics identify tobacco-use disparity in lung gene expression of ACE2, the receptor of 2019-nCov. MedRxiv 2020; published online Feb 27. doi: https://doi.org/10.1101/2020.02.05.20020107.

63 Liu LS, Wu ZS, Wang JG, et al. Prevalence of hypertension in Chinese population. In: China hypertension prevention and control guidelines revision committee, ed. Guidelines for the prevention and treatment of hypertension in China, revised edition, 2018. People’s Health Press. 14 Beijing: Oct 2018: 12–13 (Chinese).

64 Hu SS, Gao RL, Wang Z, et, al. The prevalence of cardiovascular disease. In: National Center for Cardiovascular Diseases, China, ed. REPORT ON CARDIOVASCULAR DISEASES IN CHINA 2018. Encyclopedia of China Publishing House. Beijing: Jan 2019: 12. PDF: http://117.128.6.21/cache/nccd.org.cn/Sites/Uploaded/File/2019/8/14-%E4%B8%AD%E5%9B%BD%E5%BF%83%E8%A1%80%E7%AE%A1%E7%97%85%E6%8A%A5%E5%91%8A2018%EF%BC%88%E4%B8%AD%E6%96%87%E7%89%88%EF%BC%89.pdf?ich_args2=462-11095509026575_6043db1fec4c58b2fce944dd03e44b67_10001002_9c896c2fdfcaf0d6973b518939a83798_2e40ada2aadd6db8d534753403e6cc64 (Chinese).

65 Wang LD, Liu JM, Yang Y, Peng B, Wang YL. The Prevention and Treatment of Stroke Still Face Huge Challenges——Brief Report on Stroke Prevention and Treatment in China. Chinese Circulation Journal 2018; 34(02):105–119 (Chinese).

66 IDF DIABETES ATLAS, 9th edition 2019. Diabetes estimates in China (20-79 y). 2019. https://diabetesatlas.org/data/en/country/42/cn.html.

67 Wang C, Xu J, Yang L, et al. Prevalence and risk factors of chronic obstructive pulmonary disease in China (the China Pulmonary Health [CPH] study): a national cross-sectional study. Lancet 2018; 391(10131):1706–1717.

68 Ma CS, Deenick EK. Human T follicular helper (Tfh) cells and disease. Immunol Cell Biol 2014; 92(1):64–71.

69 Xu Z, Shi L, Wang YJ, et al. Pathological findings of COVID-19 associated with acute respiratory distress syndrome. The Lancet Respiratory Medicine 2020; published online Feb 18. doi: https://doi.org/10.1016/S2213-2600(20)30076-X.

70 Li, G, Fan, Y, Lai, Y, et al. Coronavirus infections and immune responses. J Med Virol 2020; 92: 424–432.

71 Bai Y, Yao L, Wei T, et al. Presumed Asymptomatic Carrier Transmission of COVID-19. JAMA 2020; published online Feb 21. doi:10.1001/jama.2020.2565.

72 Hu ZL, Song C, Xu CJ, et al. Clinical Characteristics of 24 Asymptomatic Infections with COVID-19 Screened among Close Contacts in Nanjing, China. medRxiv 2020; published online Feb 25. doi: https://doi.org/10.1101/2020.02.20.20025619.

73 Lan L, Xu D, Ye G, et al. Positive RT-PCR Test Results in Patients Recovered From COVID-19. JAMA 2020; published online Feb 27. doi:10.1001/jama.2020.2783.

74 Toronto Star. Ontario couple have recovered but are still testing positive for coronavirus. Feb 13, 2020. https://www.thestar.com/politics/provincial/2020/02/13/ontario-couple-have-recovered-but-are-still-testing-positive-for-coronavirus.html.

75 Fan CB, Li K, Ding YH, Lu WL, Wang JQ. ACE2 Expression in Kidney and Testis May Cause Kidney and Testis Damage After 2019-nCoV Infection. medRxiv 2020; published online Feb 13. doi: https://doi.org/10.1101/2020.02.12.20022418.

76 Fan ZY, Chen LP, Li J, et al. Clinical Features of COVID-19 Related Liver Damage. medRxiv 2020; published online Feb 28. doi: https://doi.org/10.1101/2020.02.26.20026971.

77 Zou X, Chen K, Zou JW, Han PY, Hao J, Han ZG. The single-cell RNA-seq data analysis on the receptor ACE2 expression reveals the potential risk of different human organs vulnerable to Wuhan 2019-nCoV infection. Front. Med. 2020; published online Mar 13. doi: 10.1007/s11684-020-0754-0.

78 Diao B, Wang CH, Tan YJ, et al. Reduction and Functional Exhaustion of T Cells in Patients with Coronavirus Disease 2019 (COVID-19). medRxiv 2020; published online Feb 20. doi: https://doi.org/10.1101/2020.02.18.20024364.

79 Liao PC, Lin HH, Chiang BL, et al. Tai Chi Chuan Exercise Improves Lung Function and Asthma Control through Immune Regulation in Childhood Asthma. Evid Based Complement Alternat Med 2019; published online Oct 23. doi:10.1155/2019/9146827.

80 Zheng G, Xiong Z, Zheng X, et al. Subjective perceived impact of Tai Chi training on physical and mental health among community older adults at risk for ischemic stroke: a qualitative study. BMC Complement Altern Med 2017; 17(1):221.

81 Yu SG, Jing XH, Tang Y, et al. Acupuncture and Moxibustion and Immunity: The Actuality and Future. Zhen Ci Yan Jiu 2018; 43(12):747–753. (Chinese).

82 He Y, Guo X, May BH, et al. Clinical Evidence for Association of Acupuncture and Acupressure With Improved Cancer Pain: A Systematic Review and Meta-Analysis. JAMA Oncol 2019; 6(2):271–278.

83 Fu Q, Yang H, Zhang L, et al. Traditional Chinese medicine foot bath combined with acupoint massage for the treatment of diabetic peripheral neuropathy: A systematic review and meta-analysis of 31 RCTs. Diabetes Metab Res Rev 2020; 36(2):e3218.

84 Vowles KE, McEntee ML, Julnes PS, et al. Rates of opioid misuse, abuse, and addiction in chronic pain: a systematic review and data synthesis. Pain 2015; 156(4):569–576.

85 Buksnys T, Armstrong N, Worthy G, et al. Systematic review and network meta-analysis of the efficacy and safety of lidocaine 700?mg medicated plaster vs. pregabalin. Curr Med Res Opin 2020; 36(1):101–115.

86 Bloechliger M, Reinau D, Spoendlin J, et al. Adverse events profile of oral corticosteroids among asthma patients in the UK: cohort study with a nested case-control analysis. Respir Res 2018; 19(1):75.

