## Supplementary material for "Epidemiological, Clinical Characteristics and Risk Factors for Severe Condition and ICU Admission of COVID-19 Patients of Early Stage in China: A review": not applicable

| Table S1 Percentages of epidemiological and clinical characteristics among the confirmed patients: In/ outside of former epicenter (median, IQR) | | | | |
|---|---|---|---|---|
| **Subject** | **Total, %** | **In former epicenter, %** | **Outside former epicenter, %** | **P** |
| **Epidemiological characteristics and comorbidities** | | | | |
| Male | 53.45(48.48-58.03) | 51.45(47.93-56.05) | 54.15(48.94-58.15) | 0.394 |
| Age>45-60, y | 51.60(34.65-62.00) | 61.50(52.95-68.50) | 34.65(29.03-48.53) | 0.001 |
| Smoking history | 7.00(6.55-24.00) | 7.00(6.55-22.75) | 14.29(6.40-26.50) | 1 |
| Exposure to confirmed patients | 31.00(18.20-43.96) | / | 31.00(18.20-43.96) | / |
| Exposure B* | 51.80(30.57-66.52) | 7.90(1.75-39.00) | 57.97(46.03-77.90) | 0.001 |
| Severe condition | 18.00(9.60-32.20) | 48.30(36.00-56.20) | 15.70(7.95-26.55) | 0.007 |
| Death | 0.75(0-9.53) | 10.70(3.68-15.43) | 0(0-0.73) | 0.001 |
| Any comorbidities | 48.00(31.08-58.08) | 46.75(37.10-53.20) | 28.60(18.10-47.50) | 0.031 |
| Hypertension | 13.70(6.70-27.50) | 27.50(15.00-30.00) | 8.80(5.23-17.92) | 0.003 |
| Cardiovascular disease | 5.20(2.68-12.25) | 7.30(5.20-15.00) | 2.93(2.10-6.45) | 0.01 |
| Cerebrovascular disease | 2.80(2.03-4.73) | 4.10(2.35-5.70) | 2.25(1.40-3.33) | 0.2 |
| COPD | 3.70(1.50-5.90) | 2.45(1.48-9.43) | 4.00(1.50-5.96) | 0.864 |
| Diabetes | 9.60(5.86-12.00) | 12.05(10.35-16.25) | 6.70(5.00-9.80) | <0.001 |
| Digestive disease | 5.10(3.91-5.77) | 5.10(4.55-9.55) | 4.56(3.64-5.77) | 0.686 |
| Chronic liver disease | 2.90(1.96-6.00) | 2.90(2.00-5.70) | 2.82(1.62-10.03) | 0.694 |
| Immunodeficiency/HIV | 1.40(0.10-4.45) | 3.65(1.40-5.90) | 0.20(0-3.00) | 0.4 |
| Malignancy | 2.00(1.00-5.28) | 2.50(1.00-6.38) | 1.67(0.99-4.50) | 0.46 |
| Chronic kidney disease | 2.00(1.33-2.85) | 2.80(1.20-3.40) | 1.98(1.43-2.30) | 0.556 |
| **Signs, symptoms and complications** | | | | |
| Fever | 83.15(76.13-92.15) | 91.10(81.40-96.00) | 79.38(75.60-87.55) | 0.073 |
| Highest temperature > 39℃ | 10.00(4.10-15.72) | 17.70(9.60-34.00) | 7.55(3.85-13.33) | 0.112 |
| Chills | 11.30(9.10-14.79) | / | 11.30(9.10-14.79) | / |
| Cough | 62.60(50.00-75.38) | 65.25(50.00-75.25) | 62.55(49.05-75.75) | 0.708 |
| Sputum production | 18.10(13.00-34.69) | 25.15(7.03-30.10) | 17.90(14.00-38.80) | 0.733 |

| | | | | |
|---|---|---|---|---|
| Dyspnea | 14.30(7.70-28.00) | 31.10(17.93-56.00) | 10.70(6.90-21.40) | 0.003 |
| Chest tightness | 15.99(8.35-31.39) | 31.25(23.70-59.00) | 10.74(8.10-28.22) | 0.146 |
| Fatigue | 31.70(15.68-53.93) | 54.90(27.95-72.30) | 28.00(15.65-44.90) | 0.157 |
| Myalgia | 16.00(10.85-29.00) | 14.06(10.85-34.55) | 16.00(9.55-27.25) | 0.974 |
| Headache | 8.57(6.60-12.80) | 8.00(6.20-11.30) | 9.36(6.90-14.00) | 0.295 |
| Sore throat | 9.00(6.30-14.50) | 5.30(3.28-15.95) | 9.25(6.63-14.79) | 0.29 |
| Diarrhea | 7.70(3.60-12.20) | 10.10(3.00-14.00) | 6.69(3.95-10.00) | 0.779 |
| Inappetence | 14.67(6.43-28.50) | 31.80(12.20-39.90) | 7.20(4.40-25.27) | 0.117 |
| Asymptomatic infection | 4.10(2.08-6.78) | 3.60(1.80-5.40) | 4.45(2.23-7.75) | 0.429 |
| ARDS | 3.20(0-18.30) | 19.60(17.00-29.00) | 1.00(0-6.30) | 0.024 |
| Acute cardiac injury | 7.20(0-9.70) | 9.60(7.20-12.00) | 0(0-7.40) | 0.4 |
| Other acute respiratory injury | 0(0-8.00) | 8.00(8.00-8.00) | 0 | 0.667 |
| Acute kidney injury | 3.30(0-4.53) | 3.60(3.00-7.00) | 0(0-3.70) | 0.4 |
| Shock | 2.35(0-7.43) | 7.00(4.00-8.70) | 0(0-0.70) | 0.1 |
| Secondary infection | 13.75(10.00-17.50) | 10.00(10.00-10.00) | 17.50(17.50-17.50) | 1 |
| **Radiological and laboratory findings** | | | | |
| Bilateral involvement | 81.85(68.81-92.85) | 91.15(78.30-95.45) | 80.65(60.00-84.30) | 0.028 |
| Local patchy opacity | 24.38(8.43-30.70) | 8.80(4.90-27.20) | 30.00(19.73-47.14) | 0.014 |
| Ground glass opacity | 75.00(29.37-92.15) | 37.35(16.70-81.25) | 80.30(50.90-93.30) | 0.216 |
| Consolidation | 41.80(19.45-49.73) | 24.45(21.60-27.30) | 43.70(19.75-58.58) | 0.4 |
| Pleural effusion | 5.05(4.00-12.85) | / | 5.05(4.00-12.85) | / |
| Pleural thickening | 53.80(12.30-66.80) | / | 53.80(12.30-66.80) | / |
| Interlobular septal thickening | 60.85(45.25-77.33) | / | 60.85(45.25-77.33) | / |
| Crazy-paving pattern | 36.10(20.50-41.10) | / | 36.10(20.50-41.10) | / |
| Air bronchogram | 34.10(13.68-72.38) | / | 34.10(13.68-72.38) | / |
| Lymph node enlargement | 10.70(1.00-20.00) | / | 10.70(1.00-20.00) | / |
| No abnormality | 5.00(1.35-11.42) | 0.70(0-2.75) | 6.15(2.58-11.43) | 0.019 |
| White blood cell count > 10 × $10^9$/L | 10.00(2.80-20.00) | 19.00(12.05-27.00) | 5.98(1.93-9.58) | 0.007 |
| White blood cell count < 4× $10^9$/L | 23.60(17.60-33.18) | 23.60(19.10-33.91) | 23.33(16.35-35.10) | 0.826 |
| Neutrophil count > 5.8-7×$10^9$/L | 4.03(2.40-20.00) | 6.68(1.56-11.80) | 4.03(2.85-22.50) | 0.571 |

| | | | | |
|---|---|---|---|---|
| Neutrophil count < 1.8-2×10⁹/L | 11.40(7.53-24.46) | 25.00(25.00-25.00) | 10.99(6.70-22.82) | 0.5 |
| Lymphocyte count < 0.8-1.5×10⁹/L | 50.20(33.60-62.45) | 58.90(35.00-72.30) | 43.00(32.90-59.52) | 0.168 |
| Hemoglobin < 110-130 g/L | 37.23(29.21-43.50) | 41.00(38.20-51.00) | 36.25(8.10-36.25) | 0.1 |
| Platelet count < 100-150×10⁹/L | 12.00(7.69-17.00) | 14.50(6.75-24.13) | 10.99(7.69-15.38) | 0.604 |
| D-dimer > 0.5 mg/L | 20.80(14.09-44.00) | 36.00(17.75-45.44) | 10.00(5.90-14.09) | 0.095 |
| Alanine aminotransferase > 40-50 U/L | 17.00(7.90-25.65) | 24.15(18.63-27.20) | 8.10(5.72-21.60) | 0.111 |
| Aspartate aminotransferase >40-50 U/L | 19.36(12.28-22.20) | 24.00(20.60-37.00) | 16.10(10.85-21.80) | 0.064 |
| LDH > 240-250 U/L | 38.15(25.56-63.25) | 73.00(52.00-76.00) | 30.20(21.25-43.00) | 0.033 |
| Total bilirubin > 21-26 mmol/L | 7.20(2.92-13.43) | 8.80(3.00-18.00) | 5.60(2.68-11.90) | 0.7 |
| Prothrombin time > 13.5-17 s | 11.41(5.00-50.00) | 27.50(5.00-50.00) | 11.41(11.41-11.41) | 1 |
| Prothrombin time < 10-10.5 s | 3.33(2.74-23.44) | 16.45(2.90-30.00) | 3.22(2.68-3.75) | 0.667 |
| Creatine kinase >185-200 U/L | 8.05(7.20-10.90) | 8.30(6.70-33.00) | 8.03(7.30-10.45) | 0.905 |
| Serum creatinine > 73-87 μmol/L | 16.30(2.25-23.23) | 17.60(17.60-17.60) | 9.80(1.73-25.72) | 0.8 |
| Serum creatinine >111-133 μmol/L | 5.00(2.75-13.80) | 10.00(3.00-17.60) | 3.75(2.50-5.00) | 0.4 |
| Procalcitonin > 0.5 ng/mL | 21.75(4.63-41.43) | 21.75(6.50-40.30) | 22.09(1.57-49.75) | 0.886 |
| Blood urea nitrogen > 8-10 mmol/L | 12.00(4.25-15.65) | 13.15(7.50-16.33) | 2.50(2.50-2.50) | 0.4 |
| C-reactive protein > 3-6 mg/L | 62.86(52.00-83.90) | 83.90(64.00-91.90) | 54.44(47.50-62.20) | 0.021 |
| C-reactive protein > 8-10 mg/L | 60.70(52.00-75.20) | 78.25(67.20-82.85) | 55.00(46.00-62.86) | 0.006 |
| **Treatment** | | | | |
| Oxygen therapy | 66.70(43.75-76.00) | 74.00(65.00-76.20) | 43.75(21.25-78.32) | 0.247 |
| Invasive mechanical ventilation | 0.70(0-8.80) | 6.90(3.00-12.67) | 0(0-0.35) | 0.03 |
| Non-invasive mechanical ventilation | 5.40(0.34-12.48) | 11.95(5.65-24.20) | 0.67(0-5.30) | 0.009 |
| CRRT | 5.90(2.58-8.00) | 6.45(2.56-8.50) | 3.70(3.70-3.70) | 0.8 |
| ECMO | 2.90(0-3.00) | 2.90(1.45-4.00) | 0 | 0.19 |
| Antibiotic treatment | 71.00(47.70-95.13) | 91.20(78.95-99.50) | 56.45(44.03-84.76) | 0.065 |
| Antiviral treatment | 98.00(89.45-100.00) | 93.00(76.60-98.00) | 100.00(92.72-100.00) | 0.109 |
| Corticosteroid therapy | 26.35(14.63-41.40) | 30.90(22.00-50.00) | 15.00(8.95-28.35) | 0.055 |
| Intravenous immunoglobulin therapy | 20.00(12.75-32.10) | 27.00(10.80-32.10) | 17.25(13.19-41.75) | 1 |
| Antifungal treatment | 0(0-15) | 15.00(15.00-15.00) | 0 | 0.667 |
| TCM | 84.65(3.54-93.85) | 43.25(2.90-83.60) | 88.75(24.24-97.95) | 0.267 |

| | | | | |
|---|---|---|---|---|
| ICU | 13.75(1.50-24.15) | 23.50(20.30-29.05) | 2.00(0-9.35) | 0.008 |
| Percentage of patients in severe condition or not of each subject is in median and interquartile ranges (IQR). Exposure B*: Exposure to the Huanan Seafood Market for studies conducted in former epicenter, and exposure to former epicenter for studies outside of former epicenter. COPD: Chronic obstructive pulmonary disease; ARDS: Acute respiratory distress syndrome; LDH: Lactate dehydrogenase; CRRT: Continuous renal replacement therapy; ECMO: Extracorporeal membrane oxygenation; TCM: Traditional Chinese medicine; ICU: Intensive unit care. | | | | |

**Table S2 Details of hospital(s), city/cities, province(s), admission period and number of patients in each study**

| | Study | Hospital | City | Province | Admission period | Number |
|---|---|---|---|---|---|---|
| In former epicenter | Huang 2020 A | Jinyintan Hospital | Wuhan | Hubei | Dec 16th, 2019- Jan 2th, 2020 | 41 |
| | Chen 2020 A | Jinyintan Hospital | Wuhan | Hubei | Jan 1th, 2020- Jan 20th, 2020 | 99 |
| | Wang 2020 A | Wuhan University Zhongnan Hospital | Wuhan | Hubei | Jan 1th, 2020- Jan 28th, 2020 | 138 |
| | Huang 2020 B | Wuhan University Zhongnan Hospital | Wuhan | Hubei | Dec 21th, 2020- Jan 28th, 2020 | 34 |
| | Chen | Tongji Hospital affiliated to Tongji Medical College of | Wuhan | Hubei | Jan, 2020 | 29 |

| | | | | | | |
|---|---|---|---|---|---|---|
| | 2020 B | Huazhong University of Science and Technology | | | | |
| | Zhang 2020 B | No. 7 Hospital of Wuhan | Wuhan | Hubei | Jan 16th, 2020-Feb 3th, 2020 | 140 |
| | Mao 2020 A | Main District, West Branch, and Tumor Center of the Union Hospital of Huazhong University of Science and Technology | Wuhan | Hubei | Jan 16th, 2020-Feb 19th, 2020 | 214 |
| | Cao 2020 | Wuhan University Zhongnan Hospital | Wuhan | Hubei | Jan 3th, 2020-Feb 1th, 2020 | 102 |
| | Li 2020 A | the Department of Thoracic Surgery of Tongji Hospital affiliated to Tongji Medical College of Huazhong University of Science and Technology | Wuhan | Hubei | Jan 1th, 2020-Feb 20th, 2020 | 25 |
| | Liu 2020 A | Nine tertiary hospitals in Hubei province | Wuhan, Shiyan, Jingzhou, Yichang, Xiaogan and Enshi | Hubei | Dec 30th, 2020-Jan 24th, 2020 | 137 |
| | Wang 2020 B | Xiaogan Hospital | Xiaogan | Hubei | Jan 25th, 2020-Feb 9th, 2020 | 114 |
| | Bin 2020 | Wuhan Huangpi district hospital of traditional Chinese medicine | Wuhan | Hubei | Jan 29th, 2020-Feb 16th, 2020 | 55 |
| | Chen 2020 D | Wuhan east lake district people's hospital | Wuhan | Hubei | Jan 3th, 2020-Feb 7th, 2020 | 64 |
| | Wang 2020 C | Yangtze river shipping general hospital | Wuhan | Hubei | Jan 20th, 2020-Feb 14th, 2020 | 96 |
| | Din 2020 | Wuhan central hospital affiliated to Tongji Medical College of Huazhong University of Science and Technology | Wuhan | Hubei | Jan 1th, 2020-Feb 3th, 2020 | 56 |
| One province outside of former epicenter | Chang 2020 | 1. Beijing Tsinghua Changgung Hospital;<br>2. Beijing Anzhen Hospital;<br>3.Chinese PLA General Hospital | Beijing | | Jan 16th, 2020-Jan 29th, 2020 | 13 |
| | Tian 2020 | Beijing Emergency Medical Service (EMS) | | | Jan 20th, 2020-Feb 10th, 2020 | 262 |
| | Yu 2020 | The fifth medical center of PLA general hospital | | | Jan 21th, 2020- | 25 |

| | | | | | | |
|---|---|---|---|---|---|---|
| | A | | | | Feb 2th, 2020 | |
| | Chen 2020 C | Shanghai Public Health Clinical Center (SPHCC) | Shanghai | | Jan 20th, 2020-Feb 6th, 2020 | 249 |
| | Peng 2020 | Zhoupu Hospital | | | Jan 23th, 2020-Feb 16th, 2020 | 11 |
| | Dai 2020 | 13 tertiary hospitals in Jiangsu Province | Not reported | Jiangsu Province | Jan 10th, 2020-Feb 7th, 2020 | 234 |
| | Wu 2020 A | 1. the First People's Hospital of Yancheng City;<br>2. the second People's Hospital of Yancheng City;<br>3. the Fifth People's Hospital of Wuxi | Yancheng, Wuxi | | Jan 22th, 2020-Feb 14th, 2020 | 80 |
| | Xu 2020 D | the Third Hospital of Changzhou | Changzhou | | Jan 22th, 2020-Feb 18th, 2020 | 51 |
| | Huang 2020 C | The second people's hospital of Guangdong province | Guangzhou | Guangdong | Jan 23th, 2020-Feb 14th, 2020 | 35 |
| | Xu 2020 C | Guangzhou Eighth People's Hospital | | | Jan 23th, 2020-Feb 4th, 2020 | 90 |
| | Liu 2020 C | Six tertiary hospitals in Anhui Province | Not reported | Anhui | Jan 21th, 2020-Feb 3th, 2020 | 73 |
| | Li 2020 C | Fuyang second people's hospital | Fuyang | | Feb 5th, 2020-Feb 27th, 2020 | 49 |
| | Xu 2020 E | The first affiliated hospital of Bengbu medical college | Bengbu | | Jan 21th, 2020-Feb 8th, 2020 | 32 |
| | Liu 2020 D | Wenzhou sixth people's hospital | Wenzhou | Zhejiang | Jan 17th, 2020-Feb 19th, 2020 | 42 |
| | Qian 2020 | Five tertiary hospitals in Anhui Province | Not reported | | Jan 20th, 2020-Feb 11th, 2020 | 91 |
| | Xu 2020 A | 1. First Affiliated Hospital, Zhejiang University;<br>2. Wenzhou Central Hospital;<br>3. Enze Medical Center(Group) Enze Hospital;<br>4. First People's Hospital of Wenling; | Hangzhou, Wenzhou, Taizhou, Wenling, Zhoushan and Ningbo | | Jan 10th, 2020-Jan 26th, 2020 | 62 |

| | | | | | | |
|---|---|---|---|---|---|---|
| | | 5. Affiliated Zhoushan Hospital, Wenzhou Medical University;<br>6. Affiliated Yinzhou Hospital, Ningbo University;<br>7. TaiZhou Hospital | | | | |
| | Yang 2020 | 1. Wenzhou central hospital;<br>2. Ruian people's hospital;<br>3. Yueqing people's hospital | Wenzhou | | Jan 17th, 2020-Feb 10th, 2020 | 149 |
| | Wang 2020 D | Jinhua city central hospital | Jinhua | | Jan 22th, 2020-Feb 7th, 2020 | 17 |
| | Yu 2020 B | Hangzhou xixi hospital | Hangzhou | | Jan 20th, 2020-Feb 12th, 2020 | 87 |
| | Lo 2020 | the Centro Hospitalar Conde de São Januário (CHCSJ) | Macau | Macau | Jan 21th, 2020-Feb 16th, 2020 | 10 |
| | Tang 2020 | Chongqing public health medical treatment center | Chongqing | | Jan 24th, 2020-Feb 4th, 2020 | 83 |
| | Wan 2020 | the Chongqing University Three Gorges Hospital | | | Jan 23th, 2020-Feb 8th, 2020 | 135 |
| | Wu 2020 B | 1. Chongqing Three Gorges Central Hospital;<br>2. The Second Affiliated Hospital of Chongqing Medical University | | | Jan 2020-Feb 2020 | 80 |
| | Zhao 2020 | The database of the Radiology Quality Control Center, Hunan | Changsha, YueYang, ChangDe and Xiang-Tan | Hu'nan | Not reported | 101 |
| | Zheng 2020 | the North Hospital of Changsha First Hospital (Changsha Public Health Center) | Changsha | | Jan 17th, 2020-Feb 7th, 2020 | 161 |
| | Diao 2020 | West China Hospital of Sichuan University | Chengdu | Sichuan | Jan 17th, 2020-Feb 26th, 2020 | 15 |
| | Liang 2020 B | To the first affiliated hospital of He'nan university of science and technology | Luoyang | He'nan | From Jan 22th | 28 |
| | Lv 2020 | 1. Tianjin third central hospital;<br>2. Tianjin fourth central hospital; | Tianjin | | Jan 20th, 2020-Feb 12th, 2020 | 17 |

| | | | | | | |
|---|---|---|---|---|---|---|
| | | 3. The second hospital of Tianjin medical university | | | | |
| | Mao 2020 B | Ankang hospital of infectious diseases | Ankang | Shaanxi | As of Feb 18th, 2020 | 24 |
| | Xiang 2020 | The first affiliated hospital of Nanchang university | Nanchang | Jiangxi | Jan 21th, 2020-Jan 27th, 2020 | 49 |
| >1 and <30 provinces | Feng 2020 | Three hospitals in Wuhan, Shanghai and Anhui | Wuhan, Shanghai and Tongling | Hubei, Shanghai and Anhui | Jan 1th, 2020-Feb 15th, 2020 | 476 |
| | Li 2020 B | 1. The second affiliated hospital of Chongqing medical university;<br>2. Chongqing Three Gorges Central Hospital;<br>3. Yanzhuang Central Hospital of Gangcheng District | Chongqing, Ji'nan | Chongqing, Shandong | Jan 2020-Feb 2020 | 83 |
| | Liu 2020 B | 1. The first hospital of lanzhou university;<br>2. Shenyang sixth people's hospital;<br>3. Ankang city central hospital;<br>4. Lishui city central hospital;<br>5. Zhenjiang third people's hospital;<br>6. Baoding people's hospital;<br>7. Linxia state people's hospital | Lanzhou, Shenyang, Ankang, Lishui, Zhenjiang, Baoding and Linxia | Gansu, Liaoning, Shaanxi, Zhejiang, Jiangsu, Hebei | Jan 23th, 2020-Feb 8th, 2020 | 32 |
| | Xu 2020 B | 1. The Fifth Medical Center of Chinese PLA General Hospital;<br>2. Affiliated Hospital of Hebei University;<br>3. Baoding people's hospital | Beijing, Baoding | Beijing, Hebei | Jan 2020-Feb 2020 | 41 |
| | Gao 2020 | 1. Hospital of infectious diseases, Xianyang city central hospital;<br>2. Liaocheng infectious disease hospital | Xianyang, Liaocheng | Shaanxi, Shandong | 1. Jan 21th, 2020-Feb 16th, 2020;<br>2. Jan 30th, 2020-Feb 16th, 2020 | 40 |
| 30-31 provinces (nationwide) | Guan 2020 | 30 provinces in China | Nationwide | | Dec 11th, 2019-Jan 29th, 2020 | 1099 |
| | Zhang 2020 A | 31 provinces in China | | | As of Feb 11th, 2020 | 44672 |

| | | | | | |
|---|---|---|---|---|---|
| | Liang 2020 A | 31 provinces in China | | Nov 21$^{th}$, 2019-Jan 31$^{th}$, 2020 | 1590 |

### Table S3 Epidemiological characteristics of the confirmed patients in former epicenter a

| | | Huang 2020 A | Chen 2020 A | Liu 2020 A | Wang 2020 A | Huang 2020 B | Chen 2020 B | Zhang 2020 B | Mao 2020 A |
|---|---|---|---|---|---|---|---|---|---|
| **Total** | | 41 | 99 | 137 | 138 | 34 | 29 | 140 | 214 |
| **Male** | | 30 (73%) | 67 (68%) | 61 (44.5%) | 75 (54.3%) | 14 (41.2%) | 21 (72%) | 71 (50.7%) | 87 (40.7%) |
| **Female** | | 11 (27%) | 32 (32%) | 76 (55.5%) | 63 (45.7%) | 20 (58.8%) | 8 (28%) | 69 (49.3%) | 127 (59.3%) |
| **Age, y** | **Median (IQR)** | 49 (41-58) | NR | 57 | 56 (42-68) | NR | 56 | 57 | NR |
| | **Range** | NR | 21-82 | 20-83 | 22-92 | 26-88 | 26-79 | 25-87 | NR |
| | **≥50** | A* | 67 (67%) | NR | NR | ⩾58: 16 (47%) | NR | 98 (70%) | 124 (57.9%) |
| | **< 50** | A* | 32 (32%) | NR | NR | < 58: 18 (53%) | NR | 42 (30%) | 90 (42.1%) |
| | **Mean (SD)** | NR | 55.5 (13.1) | 55 (16) | NR | 56.24 (17.14) | NR | NR | 52.7 (15.5) |
| **Huanan seafood market exposure** | | 27 (66%) | 49 (49%) | 0 | 12 (8.7%) | NR | 2 (7%) | 0 | NR |
| **Smoking history** | | 3 (7%) | B* | NR | NR | NR | 2 (7%) | 9 (6.4%) | NR |
| **Time from onset of symptoms to first hospital admission** | | 7.0(4.0-8.0) | NR | NR | 7.0 (4.0-8.0) | NR | NR | 8 (6-11) | NR |
| **Incubation period** | | NR | NR | NR | NR | NR | NR | NR | NR |
| **Medical staff infection** | | NR | 0 | NR | 40 (29%) | NR | NR | 3 (2.1%) | NR |
| **Severe condition** | | NR | NR | NR | NR | NR | 14 (48.3%) | NR | NR |
| **Discharge** | | 28 (68%) | 31 (31%) | 44 (32.1%) | 47 (34.1%) | D* | E* | NR | NR |
| **Hospitalization** | | 7 (17%) | 57 (58%) | 77 (56.2%) | 85 (61.6%) | 33 (97.1%) | NR | NR | NR |
| **Death** | | 6 (15%) | 11 (11%) | 16 (11.7%) | 6 (4.3%) | NR | 2 (6.9%) | NR | NR |

Continuous data are median (IQR) or mean (SD); Categorical data are n (N%). NR: not reported. A*: 20 (49%) of the patients were 25–49, and 14 (34%) were 50–64; B*: Two cases of old male death with a long history of smoking but no previous chronic underlying disease; C*: Mean (95th percentile): 5.2 (12.5) for all 425 cases; D*: Home isolation was reported in 1 (2.9％), and unknown in 23 (67.6％); E*: Nine cases reported as alleviated.

#### Table S4 Epidemiological characteristics of the confirmed patients in former epicenter b

| | | Cao 2020 | Li 2020 A | Wang 2020 B | Bin 2020 | Chen 2020 D | Din 2020 | Wang 2020 C |
|---|---|---|---|---|---|---|---|---|
| **Total** | | 102 | 25 | 114 | 55 | 64 | 56 | 96 |
| **Male** | | 53 (52%) | 12 (48%) | 58 (50.9%) | 31 (56.4%) | 31 (48.4%) | 30 (53.6%) | 46 (47.9%) |
| **Female** | | 49 (48%) | 13 (52%) | 56 (49.1%) | 24 (43.6%) | 33 (51.6%) | 26 (46.4%) | 50 (52.1%) |
| | **Median (IQR)** | 54 (37-67) | NR | 53 | NR | NR | NR | NR |
| | **Range** | NR | NR | 23-78 | NR | 28-84 | 24-86 | NR |
| **Age, y** | **≥50** | NR | > 51: 12 (48%) | 70 (61.4%) | NR | 40 (62.5%) | NR | > 60: 59 (61.5%) |
| | **< 50** | NR | ≤51: 13 (52%) | 46 (40.4%) | NR | 24 (37.5%) | NR | ≤60: 37 (38.5%) |
| | **Mean (SD)** | NR | A* | NR | 53.9 (17.1) | 54.55±14.04 | 54.6±15.5 | NR |
| **Exposure to the Huanan Seafood Market** | | 47 (46.1%) | 19 (76%) | 90 (78.9%) | NR | NR | 4 (7.1%) | NR |
| **Exposure to confirmed patients** | | | | | NR | NR | NR | NR |
| **Smoking history** | | NR | 7 (28%) | NR | NR | NR | NR | NR |
| **Time from onset of symptoms to first hospital admission** | | 6 (3-7) | NR | NR | NR | NR | NR | NR |
| **Incubation period** | | 3 (2-6) | NR | NR | NR | NR | NR | NR |
| **Medical staff infection** | | 24 (23.5%) | 12 (48%) | NR | NR | NR | NR | NR |
| **Hospitalized patients or outpatients** | | 10 (9.8%) | 13 (52%) | NR | NR | NR | NR | NR |
| **Severe condition** | | NR | 9 (36%) | NR | NR | NR | NR | 54 (56.2%) |
| **Discharge** | | 85 (83.3%) | 20 (80%) | NR | 10 (18.2%) | NR | 11 (19.6%) | 86 (89.6%) |
| **Hospitalization** | | 0 | | NR | NR | NR | 45 (80.4%) | 0 |
| **Death** | | 17 (16.7%) | 5 (20%) | NR | 1 (1.8%) | NR | 0 | 10 (10.4%) |
| **Others** | | BMI, kg/$m^2$: 24.4 (21.8-26.0) | | | | | | |

Continuous data are median (IQR) or mean (SD); Categorical data are n (N%). NR: not reported. A*: 60.2 (5.6%) for hospitalized patients and 35.8 (9.2%) for health care staff.

#### Table S5 Epidemiological characteristics of the confirmed patients outside former epicenter/ nationwide a

| | | Xu 2020 A | Chang 2020 | Guan 2020 | Liu 2020 B | Yang 2020 | Tian 2020 | Wu 2020 | Xu 2020 B | Zhang 2020 A |
|---|---|---|---|---|---|---|---|---|---|---|
| **Total** | | 62 | 13 | 1099 | 32 | 149 | 262 | 80 | 50 | 44672 |
| **Male** | | 35(56%) | 10(77%) | 640(58.2%) | 20(62.50%) | 81(54%) | 127(48.5%) | 39(48.75%) | 29(58%) | 22981(51.4%) |
| **Female** | | 27(44%) | 3(23%) | 459/1096(41.9%) | 12(37.50%) | 68(46%) | 135(51.5%) | 41(51.25%) | 21(42%) | 21691(48.6%) |
| **Age, y** | **Median(IQR)** | 41(32-52) | 34(34-48) | 47.0(35.0–58.0) | 38.50(26.25-45.75) | NR | 47.5(1-94) | 46.1(30.7-61.5) | NR | NR |
| | **Range** | NR | NR | NR | NR | NR | 1-94 | NR | 3-85 | NR |
| | **≥50** | >40: 27(43%) | NR | 445/1011(44%) | NR | NR | ⩾45: 139(53.1%) | 28(35%) | >50: 15(30%) | 23917(53.6%) |
| | **<50** | ⩽40: 35(56%) | NR | 566/1011(56%) | NR | NR | <45: 123(46.9%) | 52(65%) | ⩽50: 35(70%) | 20755(46.4%) |
| | **Mean(SD)** | NR | NR | NR | NR | 45.11(13.35) | NR | 46.10(15.42) | 43.9(16.8) | NR |
| **Exposure to confirmed patients** | | NR | NR | NR | NR | | NR | NR | NR | NR |
| **Exposure to the former epicenter** | | 23(37%) | 12(92.3%) A* | 193/616(31.3%) | NR | 80(53.69%) B* | 53(20.2%) | 100% C* | 30(60%) | 7753/ 11302(68.6%) D* |
| **Smoking history** | | NR | NR | 158/1085(14.6%) | NR | NR | NR | NR | NR | NR |
| **Time from onset of symptoms to first hospital admission** | | 2.0(1.0-4.3) | NR | NR | NR | 6.8(5.0) | 4.5(3.7) | NR | NR | NR |
| **Incubation period** | | 4(3-5)(n=56) | NR | 4.0(2.0–7.0) | NR | NR | 6.7(5.2) | NR | NR | NR |
| **Medical staff infection** | | NR | NR | 23(2.09%) | NR | 0 | NR | 0 | NR | 1716(3.8%) |
| **Severe condition** | | NR | NR | 173(15.74%) | NR | 0 | 46(17.6%) | 3(3.75%) | NR | 8255/ 44415(18.5%) |
| **Discharge** | | 1(2%) | 13(100%) E* | 55(5%) | NR | 73(48.99%) | 45(17.2%) | 61(76.25%) | NR | NR |
| **Hospitalization** | | 61(98%) | 0 | 1029(93.7%) | NR | 76(51.01%) | 214(81.7%) | 21(23.75%) | NR | NR |
| **Death** | | 0 | 0 | 15(1.37%) | NR | 0 | 3/342(0.9%) | 0 | NR | 1023(2.3%) |

Continuous data are median(IQR) or mean(SD); Categorical data are n(N%). NR: not reported; A*: Either visited or had family members visited Wuhan; B*: With stayed in Hubei Province except Wuhan for 5(3.36%), contacted with people from Hubei Province for 49(32.89%) and no relation with Hubei Province for 15(10.06%); C*: All with a history of epidemic in Wuhan. D*: Patients who had contacted with people from Wuhan were included; E*: All the 13 patients were recovered, but 12 were still being quarantined in hospital.

#### Table S6 Epidemiological characteristics of the confirmed patients outside former epicenter b

| | | Huang 2020 C | Xu2020 C | Liu2020 D | Qian 2020 | Chen 2020 C | Peng 2020 | Liu 2020 C | Dai 2020 | Xu 2020 D | Wan 2020 |
|---|---|---|---|---|---|---|---|---|---|---|---|
| **Total** | | 35 | 90 | 42 | 91 | 249 | 11 | 73 | 234 | 51 | 135 |
| **Male** | | 19(54.3%) | 39(43%) | 23(54.8%) | 37(40.66%) | 126(50.6%) | 5(45.5%) | 41(56.2%) | 136(58.1%) | 25(49%) | 72(53.3%) |
| **Female** | | 16(45.7%) | 51(57%) | 19(45.2%) | 54(59.34%) | 123(49.4%) | 6(54.5%) | 32(43.8%) | 98(41.9%) | 26(51%) | 63(46.7%) |
| **Age, y** | **Median(IQR)** | 41(32-52) | 50 | NR | 50(36.5–57) | 51(36-64) | NR | NR | NR | A* | 47(36-55) |
| | **Range** | 12-74 | 18-86 | NR | NR | NR | NR | NR | 7-82 | NR | 3-85 |
| | **≥50** | NR | NR | NR | 47(51.6%) | NR | NR | NR | 92(39.3%) | NR | NR |
| | **<50** | NR | NR | NR | 44(48.4%) | NR | NR | NR | 142(60.7%) | NR | NR |
| | **Mean(SD)** | 44(15.17) | NR | 42.5 | NR | NR | 40.73(11.32) | B* | NR | NR | NR |
| **Exposure to confirmed patients** | | 15(42.86%) | 86(96%) | 13(31%) | 40(43.96%) | NR | 2(18.2%) | 16(21.9%) | 78(33.3%) | NR | 23(17.0%) C* |
| **Exposure to the former epicenter** | | 20(57.14%) | | 20(47.6%) | 31(34.07%) D* | NR | 7(63.6%) | 54(74%) | 133(56.8%) | 15(29.4%) | 71(52.6%) |
| **Smoking history** | | 5(14.29%) | NR | NR | NR | NR | NR | NR | NR | NR | 9(6.7%) E* |
| **Time from onset of symptoms to first hospital admission** | | NR | NR | 1-14(mean=7.02) | NR | 4(2-7) | NR | NR | NR | NR | NR |
| **Incubation period** | | NR | NR | NR | 6(3-8) | NR | NR | NR | NR | NR | NR |
| **Medical staff infection** | | NR | NR | NR | 0 | NR | NR | NR | 3(1.3%) | NR | NR |
| **Severe condition** | | NR | NR | 4(9.5%) | 9(9.9%) | NR | NR | 24(32.9%) | 15(6.4%) | 0 | 40(29.6%) |
| **Discharge** | | NR | NR | 42(100%) | 31(34.07%) | 215(86.3%) | NR | Recovered: 12(16.4%) | NR | NR | 120(88.9%) |
| **Hospitalization** | | NR | NR | NR | 60(65.93%) | 32(12.9%) | NR | NR | NR | NR | NR |
| **Death** | | NR | NR | 0 | 0 | 2(0.8%) | NR | NR | NR | 0 | 1(0.7%) |

Continuous data are median(IQR) or mean(SD); Categorical data are n(N%). NR: not reported; A*: 35.0(29.0-51.0) for imported cases(n=15), 37.0(24.0-47.5) for secondary cases(n=17) and 53.0(35.0-65.0) for tertiary cases(n=19). B*: 29.2±10.9 in mild type(n=6), 33.4±12.2 in common type(n=43), 44.2±12.0 in severe type(n=21), 63.0±21.2 in critical type(n=3). C*: Contact with suspected patients in epidemic area 19(14.1%). D*: And vehicle exposure(e.g. airplane, coach, ship): 11(12.09%), contact with Wuhan personnel: 8(8.79%). E*: Current smoking 9(6.7%).

#### Table S7 Epidemiological characteristics of the confirmed patients outside former epicenter/ nationwide c

| | | Li 2020 B | Wu 2020 B | Tang 2020 | Zheng 2020 | Zhao 2020 | Lo 2020 | Feng 2020 | Liang 2020 A | Diao 2020 | Gao 2020 |
|---|---|---|---|---|---|---|---|---|---|---|---|
| **Total** | | 83 | 80 | 83 | 161 | 101 | 10 | 476 | 1590 | 15 | 40 |
| **Male** | | 44(53%) | 42(52%) | 44(53%) | 80(49.7%) | 56(55.4%) | 3(30%) | 271(56.9%) | 904/1578(57.3%) | 8(53.3%) | 19(47.5%) |
| **Female** | | 39(47%) | 38(48%) | 39(47%) | 81(50.3%) | 45(44.6%) | 7(70%) | 205(43.1%) | 674/1578(42.7%) | 7(46.7%) | 21(52.5%) |
| **Age, y** | **Median(IQR)** | NR | 50 | NR | 45(33.5-57) | NR | 54(27-64) | 53(40-64) | NR | NR | NR |
| | **Range** | NR | 15-79 | 10-77 | NR | 17-75 | NR | NR | NR | NR | NR |
| | **≥50** | NR | NR | NR | NR | > 50: 29(28.7%) | > 60: 3(30%) | ⩾65: 118(24.8%) | NR | NR | NR |
| | **<50** | NR | NR | NR | NR | ⩽50: 72(71.3%) | ⩽60: 7(70%) | < 65: 358(75.2%) | NR | NR | NR |
| | **Mean(SD)** | 45.5(12.3) | 44(11) | 44.9(15.8) | NR | 44.44 | 40.73(11.32) | NR | 48.9(16.3) | NR | 41±16.4 |
| **Exposure to confirmed patients** | | NR | NR | 39(47%) | NR | 12(11.9%) | A* | NR | NR | B* | NR |
| **Exposure to the former epicenter** | | NR | NR | 38(45.8%) | 104(64.6%) | 84(83.2%) | | 425(89.3%) | 1334(83.9%) | 7(46.7%) | 16(40%) |
| **Smoking history** | | NR | 26(33%) | NR | NR | NR | C* | 44/454(9.7%) | 111(7.0%) | NR | NR |
| **Time from onset of symptoms to first hospital admission** | | 7(4.75-9) | 7(4) | NR | 6(3-8) | NR | 5(1-11) | 6(4-10) | NR | NR | (1-13, range),(4.9 ±3.5) |
| **Medical staff infection** | | NR | NR | NR | NR | NR | 0 | NR | NR | NR | NR |
| **Severe condition** | | 25(30.1%) | NR | 13(15.7%) | 30(18.6%) | 14(13.9%) | 4(40%) | 124(26.1%) | 131(8.24%) | NR | 1(2.5%) |
| **Discharge** | | NR | NR | NR | NR | NR | 5(50%) | 287/300(95.7%) | NR | NR | 14(35%) |
| **Hospitalization** | | NR | NR | NR | NR | NR | 5(50%) | 11/300(3.7%) | NR | NR | 26(65%) |
| **Death** | | NR | NR | NR | NR | NR | 0 | 2/300(0.7%) | 50(3.14%) | NR | 0 |

Continuous data are median(IQR) or mean(SD); Categorical data are n(N%). NR: not reported; A*: Visited local hospital in Wuhan, China 2(20%), visited local hospital in Zhuhai, China 1(10%). B*: Close contact with people from Wuhan or confirmed patients 5(33.3%). C*: Ex-smoker 2(20%), current smoker 0.

#### Table S8 Epidemiological characteristics of the confirmed patients outside former epicenter d

| | | Liang 2020 B | Lv 2020 | Mao 2020 B | Wang 2020 D | Xiang 2020 | Xu 2020 E | Yu 2020 A | Yu 2020 B | Li 2020 C |
|---|---|---|---|---|---|---|---|---|---|---|
| **Total** | | 28 | 17 | 24 | 17 | 49 | 32 | 25 | 87 | 49 |
| **Male** | | 16(57.1%) | 12(70.6%) | 14(58.3%) | 10(58.8%) | 33(67.3%) | 19(59.4%) | 16(64%) | 40(46%) | 28(57.1%) |
| **Female** | | 12(42.9%) | 5(29.4%) | 10(41.7%) | 7(41.2%) | 16(32.7%) | 13(40.6%) | 9(36%) | 47(54%) | 21(42.9%) |
| **Age, y** | **Median(IQR)** | NR | NR | NR | NR | NR | NR | NR | NR | 45(32-60) |
| | **Range** | 24-87 | 28-75 | 4-83 | 19-86 | 18-78 | NR | 3-79 | 4-88 | 14-82 |
| | **≥50** | > 60: 11(39.3%) | NR | NR | NR | NR | NR | > 45: 8(22%) | NR | NR |
| | **<50** | ≤60: 17(60.7%) | NR | NR | NR | NR | NR | ≤45: 17(68%) | NR | NR |
| | **Mean(SD)** | 51.75(19.74) | 52.6(11.6) | 43.1 | 42.1(16.6) | 42.9 | 52(31) | 37.9(17.1) | 42.89±17.02 | NR |
| **Exposure to confirmed patients** | | 3(10.7%) | NR | 5(20.8%) | 7(41.2%) | NR | 21(65.6%) | 2(8%) | 17(19.5%) | 34(69.4%) |
| **Exposure to the former epicenter** | | 10(35.7%) | NR | 19(79.2%) | 10(58.8%) | 46(93.9%) | NR | 23(92%) | 58(66.7%) | 25(51%) |
| **Smoking history** | | NR | NR | NR | NR | 3(6.1%) | NR | NR | NR | NR |
| **Incubation period** | | NR | NR | NR | 1-11(5.5±3.1) | 4.7(1-13, range) | NR | 5.5±3(3-13,range) | 6.30±3.99,(range, 1~26) | 6(2-11, range) |
| **Severe condition** | | 4(14.3%) | 4(23.5%) | NR | 2(11.8%) | 9(18.4%) | 6(18.8%) | NR | 5(5.7%) | 27(55.1%) |
| **Discharge** | | NR | NR | NR | 11(64.7%) | NR | NR | 1(4%) | NR | 49(100%) |
| **Hospitalization** | | NR | NR | NR | NR | NR | NR | 23(92%) | NR | 0 |
| **Death** | | NR | NR | NR | NR | NR | NR | 1(4%) | NR | 0 |

Continuous data are median(IQR) or mean(SD); Categorical data are n(N%). NR: not reported; A*: Close contact with family members from Wuhan to Luoyang 3(10.7%), close contact with family members in fever 1(3.6%).

#### Table S9 Comorbidities of the confirmed patients in former epicenter a

| | Huang 2020 A | Chen 2020 A | Liu 2020 A Hubei | Wang 2020 A | Huang 2020 B | Chen 2020 B | Zhang 2020 C | Mao 2020 A |
|---|---|---|---|---|---|---|---|---|
| **Any comorbidity** | 13(32%) | 50(51%) | 27(19.7%) | 64(46.4%) | 16(47.1%) | 16(55%) | 90(64.3%) | 83(38.8%) |
| **Hypertension** | 6(15%) | NR | 13(9.5%) | 43(31.2%) | 8(23.5%) | 8(28%) | 42(30.0%) | 51(23.8%) |
| **Diabetes** | 8(20%) | 12(12%) | 14(10.2%) | 14(10.1%) | 4(11.8%) | 5(17%) | 17(12.1%) | 30(14.0%) |
| **Cardiovascular disease** | 6(15%) | 40(40%) A* | 10(7.3%) | 20(14.5%) | 6(17.6%) | NR | B* | 15(7.0%) A* |
| **Cerebrovascular disease** | NR | | NR | 7(5.1%) | NR | NR | 3(2.1%) | |
| **COPD** | 1(2%) | NR | 2(1.5%) | 4(2.9%) | 2(5.9%) | NR | 2(1.4%) | NR |
| **Malignancy** | 1(2%) | 1(1%) | 2(1.5%) | 10(7.2%) | 3(8.8%) | 1(3%) | NR | 13(6.1%) |
| **Chronic liver disease** | 1(2%) | NR | NR | 4(2.9%) | 1(2.9%) | 2(6%) | 8(5.7%) | NR |
| **Chronic kidney disease** | NR | NR | NR | 4(2.9%) | NR | NR | 2(1.4%) | 6(2.8%) |
| **Immunodeficiency/HIV** | NR | NR | NR | 2(1.4%) | 2(5.9%) | NR | NR | NR |
| **Others** | NR | Digestive system disease 11(11%) | Other chronic diseases: 24(17.5%) | NR | Hyperuricemia: 1(2.9%); Hypothyroidism: 2(5.9%) | After lung cancer surgery: 1(3%) | Hyperlipidemia: 7(5.0%); Cholelithiasis: 6(4.3%); Arrhythmia: 5(3.6%); Thyroid diseases: 5(3.6%); Electrolyte imbalance: 4(2.9%); Urolithiasis: 3(2.1%); Secondary pulmonary tuberculosis: 2(1.4%); Chronic gastritis and gastric ulcer: 7(5.0%) | NR |

Categorical data are n(N%). NR: not reported. COPD: chronic obstructive pulmonary disease; HIV: human immunodeficiency virus; A*: Cardiovascular and cerebrovascular diseases; B*: Reported as aorta sclerosis 2(1.4%) and coronary heart disease 7(5.0%).

#### Table S10 Comorbidities of the confirmed patients in former epicenter b

| | Cai 2020 | Li 2020 A | Wang 2020 B | Wang 2020 C |
|---|---|---|---|---|
| **Any comorbidity** | 47(46.1%) | NR | 60(52.6%) | NR |
| **Hypertension** | 28(27.5%) | 2(8%) | 33(28.9%) | 50(52.1%) |
| **Diabetes** | 11(10.8%) | 1(4%) | 15(13.2%) A* | 25(26.1%) |
| **Cardiovascular disease** | 5(4.9%) | B* | 7(6.1%) | 5(5.2%) |
| **Cerebrovascular disease** | 6(5.9%) | NR | NR | 3(3.1%) |
| **COPD** | NR | 5(20%) | NR | Chronic bronchitis or COPD: 15(15.6%) |
| **Other respiratory diseases** | 10(9.8%) | | 5(4.4%) | |
| **Digestive diseases** | | | 5(4.4%) | Gastritis: 5(5.2%) |
| **Malignancy** | 4(3.9%) | NR | 1(0.9%) | 1(1%) |
| **Chronic liver disease** | 2(2%) | NR | NR | 1(1%) |
| **Chronic kidney disease** | 4(3.9%) | NR | NR | 1(1%) |
| **Others** | | NR | Neural system diseases:1(0.9%). | Hyperthyroidism: 2(2.1%); Phlebothrombosis: 2(2.1%). |

Categorical data are n(N%). NR: not reported. COPD: chronic obstructive pulmonary disease; A*:Endocrine system diseases. B*: Coronary heart disease: 4(16%).

#### Table S11 Comorbidities of the confirmed patients outside former epicenter a

| | Xu 2020 A | Guan 2020 | Liu 2020 B | Yang 2020 | Wu 2020 A | Zhang 2020 A |
|---|---|---|---|---|---|---|
| **Any comorbidity** | 20(32%) | 261(23.7%) | NR | 52(34.9%) | 38(47.50%) | 5446/20982(26%) |
| **Hypertension** | 5(8%) | 165(15.0%) | 1(3.13%) | NR | NR | 2683/20982(12.8%) |
| **Diabetes** | 1(2%) | 81(7.4%) | NR | 9(6.04%) A* | 5(6.25%) A* | 1102/20982(5.3%) |
| **Cardiovascular disease** | NR | NR | 1(3.13%) | 28(18.79%) B* | 25(31.25%) B* | 873/20982(4.2%) |
| **Cerebrovascular disease** | 1(2%) | 15(1.4%) | NR | | | NR |
| **COPD** | 1(2%) | 12(1.1%) | NR | NR | 0 | NR |
| **Malignancy** | NR | 10(0.9%) | 2(6.25%) | 2(1.34%) | 1(1.25%) | 107/20982(0.5%) |
| **Respiratory system diseases** | NR | NR | 1(3.13%) | 1(0.67%) | 1(1.25%) | 511/20982(2.4%) |
| **Chronic liver disease** | 7(11%) C* | NR | 1(3.13%) | NR | 1(1.25%) | NR |
| **Chronic kidney disease** | 1(2%) E* | 8(0.7%) | NR | NR | 1(1.25%) | NR |
| **Immunodeficiency/HIV** | NR | 2(0.2%) | NR | NR | 0 | NR |
| **Others** | NR | Hepatitis B infection: 23(2.1%) | NR | Digestive system diseases: 8(5.37%); Neural system diseases: 0(0%); Others: 4(2.68%) | Digestive system disease: 3(3.75%); Nervous system diseases: 1(1.25%) | NR |

Categorical data are n(N%). NR: not reported. COPD: chronic obstructive pulmonary disease; HIV: human immunodeficiency virus; A*: Endocrine diseases. B*: Cardiovascular and cerebrovascular diseases; C*: Reported as liver disease; D*: Alcoholic hepatitis; E*: Reported as renal disease.

#### Table S12 Comorbidities of the confirmed patients outside former epicenter b

| | Huang 2020 C | Xu 2020 C | Liu 2020 D | Qian 2020 | Chen 2020 C | Xu 2020 D | Wan 2020 | Li 2020 B |
|---|---|---|---|---|---|---|---|---|
| **Any comorbidity** | 4(11.43%) | 45(50%) | NR | NR | 90(36.1%) | 12(23.5%) | 43(31.9%) | 15(18.1%) |
| **Hypertension** | 1(2.86%) | 17(19%) | 9(21.4%) | 15(16.48%) | 1(5.9%) | NR | 13(9.6%) | 5(6%) |
| **Diabetes** | 2(5.71%) | 5(6%) | 2(5%) | 8(8.79%) | 25(10.0%) A* | 4(7.8%) | 12(8.9%) | 7(7.8%) |
| **Cardiovascular disease** | 1(2.86%) B* | 3(3%) | NR | 3(3.30%) C* | 55(21.7%) C* | NR | 7(5.2%) | 1(1.2%) |
| **Cerebrovascular disease** | NR | NR | NR | | | | NR | NR |
| **COPD** | 2(5.71%) | 1(1%) | NR | NR | NR | NR | 4(10%) | 5(6%) |
| **Malignancy** | NR | 10(0.9%) | NR | NR | 1(0.4%) | NR | 4(3.0%) | NR |
| **Respiratory system diseases** | NR | Tuberculosis: 2(2%) | NR | NR | 5(2.0%) | 1(1.96%) | 1(0.7%) | NR |
| **Chronic liver disease** | NR | NR | 3(7.1%) | NR | NR | 1(1.96%) | 2(1.5%) | NR |
| **Chronic kidney disease** | NR | NR | 1(2.4%) | NR | NR | 1(1.96%) | NR | NR |
| **Others** | | Others: 15(17%) | | | Digestive system diseases: 9(3.6%); Chronic hepatitis B virus infection: 2(0.8%) | | | |

Categorical data are n(N%). NR: not reported. COPD: chronic obstructive pulmonary disease; A*: Endocrine diseases. B*: Coronary heart disease. C*: Cardiovascular and cerebrovascular diseases.

#### Table S13 Comorbidities of the confirmed patients outside former epicenter c

| | Wu 2020 B | Tang 2020 | Zheng 2020 | Zhao 2020 | Lo 2020 | Feng 2020 | Liang 2020 A | Diao 2020 | Gao 2020 |
|---|---|---|---|---|---|---|---|---|---|
| **Any comorbidity** | 15(18%) | 16(19.3%) | 33(20.5%) | 40(69.7%) | 5(50%) | 205(43.1%) | 399(25.1%) | 2(13.3%) | 14(35%) |
| **Hypertension** | 4(5%) | 6(7.2%) | 22(13.7%) | NR | 3(30%) | 113(23.7%) | 269(16.9%) | 1(6.7%) | 2(5%) |
| **Diabetes** | 4(5%) | 8(9.6%) | 7(4.3%) | 3(3.0%) A* | NR | 49(10.3%) | 130(8.2%) | 1(6.7%) | 4(10%) |
| **Cardiovascular disease** | 1(1%) | 2(2.4%) | 4(2.5%) | 16(15.8%) B* | NR | 38(8%) | 59(3.7%) | NR | NR |
| **Cerebrovascular disease** | NR | 1(1.2%) | 4(2.5%) | | NR | 17(3.6%) | 30(1.9%) | NR | NR |
| **COPD** | 3(4%) | NR | 6(3.7%) | NR | NR | 22(4.6%) | 24(1.5%) | NR | NR |
| **Malignancy** | NR | NR | NR | NR | NR | 12(2.5%) | 18(1.1%) | NR | 2(5%) |
| **Respiratory system diseases** | 0 | 3(3.6%) | NR | 5(4.9%) | NR | NR | NR | NR | NR |
| **Chronic liver disease** | NR | NR | 4(2.5%) | NR | NR | NR | NR | NR | NR |
| **Hepatitis B infection** | NR | 2(2.4%) | NR | NR | Past: 1(10%) | NR | 28(1.8%) | NR | 2(5%) |
| **Chronic kidney disease** | NR | NR | NR | NR | NR | 4(0.8%) | 21(1.3%) | NR | NR |
| **Immunodeficiency/HIV** | Immunosuppression: 3(3%) | NR | NR | NR | NR | Immunosuppression: 7(1.5%) | 3(0.2%) | NR | NR |
| **Others** | | Hyperlipidemia: 1(1.2%); Gout: 2(2.4%); | | Surgical history: 7(6.9%); Digestive system disease: 6(5.9%); | Dyslipidemia: 2(20%). | Others: 103(21.6%) | | | Surgical history: 2(5%). |

Categorical data are n(N%). NR: not reported. COPD: chronic obstructive pulmonary disease; HIV: human immunodeficiency virus; A*: Endocrine diseases. B*: Cardiovascular and cerebrovascular diseases.

#### Table S14 Comorbidities of the confirmed patients outside former epicenter d

| | Li 2020 C | Liang 2020 B | Mao 2020 B | Wang 2020 D | Xiang 2020 | Yu 2020 A |
|---|---|---|---|---|---|---|
| **Any comorbidity** | NR | 18(64.3%) | 3(12.5%) | NR | 14(28.6%) | NR |
| **Hypertension** | 9(18.4%) | 9(32.1%) | NR | 2(11.8%) | 6(12.2%) | 1(4%) |
| **Diabetes** | 11(11.4%) | 7(25%) | NR | NR | 2(4.1%) | 3(12%) |
| **Cardiovascular disease** | 5(10.2%) | 4(14.3%) | 1(4.2%) | NR | NR | NR |
| **Cerebrovascular disease** | NR | 1(3.6%) | NR | NR | NR | NR |
| **COPD** | 2(4%) | NR | NR | NR | NR | NR |
| **Malignancy** | 1(2%) | NR | NR | NR | NR | NR |
| **Respiratory system diseases** | 0 | 1(3.6%) | NR | NR | NR | NR |
| **Chronic liver disease** | NR | NR | NR | NR | 6(12.2%) | NR |
| **Hepatitis B infection** | 3(6.1%) | NR | NR | NR | NR | NR |
| **Chronic kidney disease** | NR | NR | NR | NR | NR | NR |
| **Immunodeficiency/HIV** | NR | NR | NR | NR | NR | NR |
| **Others** | | Autoimmune diseases: 1(3.6%); Surgical history: 1(3.6%). | Tonsillitis: 1(4.2%); A/H1N1 influenza: 1(4.2%). | Hyperlipidemia: 1(5.9%) | | |

Categorical data are n(N%). NR: not reported. COPD: chronic obstructive pulmonary disease; HIV: human immunodeficiency virus.

#### Table S15 Signs and symptoms on admission of the confirmed patients in former epicenter a

| | Huang 2020 A | Chen 2020 A | Liu 2020 A | Wang 2020 A | Huang 2020 B | Chen 2020 B | Zhang 2020 B | Mao 2020 A |
|---|---|---|---|---|---|---|---|---|
| **Fever** | 40(98%) | 82(83%) | 112(81.8%) | 136(98.6%) | 32(94.1%) | 28(97%) | 110/120(91.7%) | 132(61.7%) |
| **Highest temperature＞39℃** | 14(34%) | NR | 20(17.7%) | NR | NR | NR | NR | NR |
| **Highest temperature＜37.5℃** | < 37.3℃: 1(2%) | NR | < 37.3℃: 28(25.0%) | NR | NR | NR | NR | NR |
| **Cough** | 31(76%) A* | 81(82%) | 66(48.2%) | 82(59.4%) A* | 17(50.0%) | Cough and sputum production: 21(72%); | 90/120(75.0%) | 107(50.0%) A* |
| **Sputum production** | 11/39(28%) | NR | 6(4.4%) | 37(26.8%) | 8(23.5%) | | NR | NR |
| **Dyspnea** | 22/40(55%) | 31(31%) | 26(19.0%) | 43(31.2%) | 5(14.7%) | 17(59%) B* | 44/120(36.7%) B* | NR |
| **Fatigue** | Myalgia or fatigue: 18(44%) | 9(9%) | Myalgia or fatigue: 44(32.1%) | 96(69.6%) | Myalgia or fatigue: 22(64.7%) | Myalgia or fatigue: 12(41%) | 90/120(75.0%) | NR |
| **Myalgia** | | 11(11%) | | 48(34.8%) | | | NR | 23(10.7%) |
| **Headache** | 3/38(8%) | 8(8%) | 13(9.5%) | 9(6.5%) | 2(5.9%) | 2(7%) | NR | 28(13.1%) |
| **Hemoptysis** | 2/39(5%) | NR | 7(5.1%) | NR | NR | NR | NR | NR |
| **Diarrhea** | 1/38(3%) | 2(2%) | 11(8.0%) | 14(10.1%) | 5(14.7%) | 4(14%) | 18/139(12.9%) | 41(19.2%) |
| **Time from onset of symptoms to shortness of breath, d** | 8.0(5.0-13.0) | NR | 7(1–20) | 5.0(1.0-10.0) | NR | NR | NR | NR |
| **Time from onset of symptoms to ARDS, d** | 9.0(8.0-14.0) | NR | NR | 8.0(6.0-12.0) | NR | NR | NR | NR |
| **Time from onset of symptoms to ICU admission, d** | 10.5(8.0-17.0) | NR | NR | 10(6-12) | NR | NR | NR | NR |
| **Respiratory rate** | 12(29%) > 24/min | NR | NR | 20(19-21) | NR | 6(21%) > 24/min | NR | NR |
| **Sore throat** | NR | 5(5%) | NR | 24(17.4%) | NR | NR | NR | 31(14.5%) |
| **Nausea or vomiting** | NR | 1(1%) | NR | Nausea: 14(10.1%); Vomit: 5(3.6%) | NR | NR | Nausea: 24/139(17.3%); Vomiting: 7/139(5.0%) | NR |
| **Dizziness** | NR | NR | NR | 13(9.4%) | NR | NR | NR | 36(16.8%) |

| | | | | | | | | |
|---|---|---|---|---|---|---|---|---|
| **Anorexia** | NR | NR | NR | 55(39.9%) | NR | NR | 17/139(12.2%) | 68(31.8%) |
| **Abdominal pain** | NR | NR | NR | 3(2.2%) | NR | NR | 8/139(5.8%) | 10(4.7%) |
| **Others** | Time from onset of symptoms to mechanical ventilation, d: 10.5(7.0-14.0) | Chest pain: 2(2%); Rhinorrhea: 4(4%); | Heart palpitations: 10(7.3%); | Time from hospital admission to ICU admission, d: 1(0-3); | NR | NR | Belching: 7/139(5.0%); | C* |

Continuous data are median(IQR) or mean(SD); Categorical data are n(N%). NR: not reported. ARDS: acute respiratory distress syndrome ICU, intensive care unit. A*: Most patients with cough were dry cough in Huang 2020 A and Mao 2020, and were all dry cough in Wang 2020; B*: Chest tightness and dyspnea. C*: Impaired consciousness: 16(7.5%); Acute cerebrovascular disease: 6(2.8%); Ataxia: 1(0.5%); Seizure: 1(0.5%); Taste impairment: 12(5.6%); Smell impairment: 11(5.1%); Vision impairment: 3(1.4%); Nerve pain: 5(2.3%).

#### Table S16 Signs and symptoms on admission of the confirmed patients in former epicenter b

| | Cao 2020 | Li 2020 A | Wang 2020 B | Bin 2020 | Chen 2020 D | Din 2020 | Wang 2020 C |
|---|---|---|---|---|---|---|---|
| **Fever** | 83(81.4%) | 24(96%) | 107(93.9%) | 39(70.9%) | 54(84.37%) | 51(91.1%) | 71(74%) |
| **Highest temperature＞39℃** | NR | NR | 11(9.6%) | NR | NR | NR | NR |
| **Highest temperature＜37.5℃** | NR | < 37.3℃: 1(4%) | < 37.3℃: 5(4.4%) | NR | NR | NR | NR |
| **Cough** | Dry cough: 50(49%) | 17(68%) | 91(79.8%) | 31(56.4%) | 45(70.31%) | 35(62.5%) | 70(72.9%) |
| **Sputum production** | NR | NR | 9(7.9%) | 20(36.4%) | NR | NR | NR |
| **Dyspnea** | NR | 20(80%) | 27(23.7%) | 3(5.5%) | NR | NR | NR |
| **Chest tightness** | NR | NR | 27(23.7%) | NR | 20(31.25%) | NR | NR |
| **Fatigue** | 56(54.9%) | Fatigue or muscular soreness: | Myalgia or fatigue: | NR | NR | Myalgia or fatigue: | 45(46.9%) |
| **Myalgia** | 35(34.3%) | 17(68%) | 44(32.1%) | NR | 9(14.06%) | 29(51.8%) | NR |
| **Headache** | NR | NR | NR | NR | 14(21.87%) | NR | 2(2.1%) |
| **Diarrhea** | 11(10.8%) | 5(20%) | 3(2.7%) | 1(1.8%) | 2(3.12%) | 2(3.6%) | 11(11.5%) |
| **Sore throat** | NR | NR | 6(5.3%) | NR | 1(1.56%) | NR | NR |
| **Abdominal pain** | NR | NR | NR | NR | NR | 1(1.8%) | NR |
| **Asymptomatic infection** | NR | NR | NR | 1(1.8%) | NR | 3(5.4%) | NR |
| **Others** | Multiple symptom: 92(90.2%) | Surgical history: 13(52%) | | Shortness of breath: 15(27.3%). | | | |

Continuous data are median(IQR) or mean(SD); Categorical data are n(N%). NR: not reported.

#### Table S17 Signs and symptoms of the confirmed patients outside of former epicenter/ nationwide a

| | Xu 2020 A | Chang 2020 | Guan 2020 | Liu 2020 B | Yang 2020 | Tian 2020 | Li 2020 B | Wu 2020 A | Xu 2020 B |
|---|---|---|---|---|---|---|---|---|---|
| **Fever** | 48(77%) | 12(92.3%) | 473/1081(43.8%) A* | 25(78.13%) | 114(76.51%) | 215(82.1%) | 12(70.6%) | 63(78.75%) | 34(86%) |
| **Highest temperature ＞39℃** | 8(13%) | NR | 38/1081(3.5%) B* | NR | NR | 9(3.4%) | NR | NR | 5(10%) |
| **Highest temperature ＜37.5℃** | ＜ 37.3℃: 14(23%) | NR | 608/1081(56.2%) C* | NR | NR | ＜ 37.3℃: 47(17.9%) | NR | NR | 7(14%) |
| **Cough** | 50(81%) | 6(46.2%) | 745(67.8%) | 22(68.72%) | 87(58.39%) | 120(45.8%) | 13(76.5%) D* | 51(63.75%) | 20(40%) |
| **Sputum production** | 35(56%) | 2(15.4%) | 370(33.7)) | NR | 48(32.21%) | NR | 3(17.6%) | NR | 7(14%) |
| **Dyspnea** | NR | NR | 205(18.7%) | NR | 2(1.34%) | 18(6.9%) | NR | 30(37.50%) | 4(8%) E* |
| **Fatigue** | Myalgia or fatigue: 32(52%) | NR | 419(38.1%) | NR | NR | 69(26.3%) | 8(47.1%) | NR | 8(16%) |
| **Myalgia** | | 3(23.1%) | Myalgia or arthralgia: 164(14.9%) | NR | 5(3.36%) | NR | 4(23.5%) | 18(22.50%) | 8(16%) |
| **Headache** | 21(34%) | 3(23.1%) | 150(13.6%) | NR | 13(8.72%) | 17(6.5%) | NR | 13(16.25%) F* | 5(10%) |
| **Hemoptysis** | 2(3%) | NR | 10(0.9%) | NR | 21(14.09%) | NR | NR | NR | NR |
| **Chills** | NR | NR | 126(11.5%) | NR | NR | NR | NR | NR | NR |
| **Chest pain** | NR | NR | NR | NR | 5(3.36%) | NR | NR | 3(3.75%) | NR |
| **Diarrhea** | 3(8%) | 1(7.7%) | 42(3.8%) | NR | 11(7.38%) | NR | 2(11.8%) | 1(1.25%) | NR |
| **Respiratory rate** | 2(3%) > 24/min | NR | NR | NR | NR | 124.5(14.8%) | NR | NR | NR |
| **Sore throat** | NR | NR | 153(13.9%) | NR | 21(14.09%) | NR | NR | 11(13.75%) | 4(8%) |
| **Rhinorrhea** | NR | 1(7.7%) | NR | NR | 5(3.36%) | NR | 2(11.8%) | 5(6.10%) | NR |
| **Nausea or vomiting** | NR | NR | 55(5.0%) | NR | 2(1.34%) | NR | NR | 1(1.25%) | NR |
| **Throat congestion** | NR | NR | 19(1.7%) | NR | NR | NR | NR | NR | NR |

| | | | | | | | | | | |
|---|---|---|---|---|---|---|---|---|---|---|
| **Others** | Oxygenation index: 379(292-448)(n=51); Arterial oxygen pressure(mm Hg): 90(80-105)(n=51) | Upper airway congestion: | 8(61.5%) | Conjunctival congestion: 9(0.8%); Nasal congestion: 53(4.8); Rash: 2(0.2%); Enlargement of lymph nodes: 2(0.2%); Tonsil swelling: 23(2.1%); | NR | Chest tightness: 16(10.74%); | NR | Dizziness: 2(11.8%) | More than one sign or symptom: 66(82.50%); Time from onset of symptoms to shortness of breath, d: 8.0(5.0-13.0) | Gastrointestinal Reaction: 1(2%); |

Continuous data are median(IQR) or mean(SD); Categorical data are n(N%). NR: not reported. A*: Fever during hospitalization: 975/1099(88.7%). B*: Highest temperature > 39℃ during hospitalization:114/926(12.3%). C*: Highest temperature < 37.5℃ during hospitalization:92/926(9.9%). D*: Including dry cough 7(41.2%); E*: Dyspnea and chest tightness. F*: Headache and mental disorder symptoms.

#### Table S18 Signs and symptoms of the confirmed patients on admission outside of former epicenter/ nationwide b

| | Huang 2020 C | Xu 2020 C | Liu 2020 D | Qian 2020 | Chen 2020 C | Peng 2020 | Liu 2020 C | Dai 2020 | Yu 2020 B | Liang 2020 A |
|---|---|---|---|---|---|---|---|---|---|---|
| **Fever** | 30(85.71%) | 70(78%) | 35(83.3%) | 65(71.43%) | 217(87.1%) | 10(91%) | 68(93.2%) | 170(72.6%) | 62(71.3%) | 1351/1536(88%) |
| **Highest temperature＞39℃** | 6(17.14%) | NR | NR | ≥ 39.1℃: 3(3.3%) | NR | NR | NR | 5(10%) | NR | NR |
| **Highest temperature＜37.5℃** | NR | NR | NR | < 37.3℃: 26(28.57%) | NR | NR | NR | 7(14%) | NR | NR |
| **Cough** | 28(80%) | 57(63%) | 26(61.9%) | 55(60.44%) | 91(36.5%) | 6(55%) | 60(82.2%) | 150(64.1%) | 45(51.7%) | Dry cough: 1052/1498 (70.2%) |
| **Sputum production** | A* | 11(12%) | NR | 30(32.97%) | NR | NR | 39(53.4%) | NR | NR | 513/1424 (36.0%) |
| **Dyspnea** | 7(20%) | NR | 9(21.4%) | 3(3.3%) | 19(7.6%) | 0 | NR | 7(3%) | NR | 331 (20.8%) |
| **Shortness of breath** | 8(22.86%) | NR | NR | 10(10.99%) | NR | NR | NR | 5(2.1%) | NR | NR |
| **Fatigue** | 14(40%) | 19(21%) | 14(33.3%) | 40(43.96%) | 39(15.7%) | NR | 55(75.3%) | 31(13.2%) | 6(6.9%) | 584/1365 (42.8%) |
| **Myalgia** | 15(42.86%) | 25(28%) | 5(11.9%) | 5(5.49%) | NR | Systemic soreness: 7(64%) | NR | 21(9%) | NR | Myalgia or arthralgia: 234/1338 (17.5%) |
| **Headache** | 3(8.57%) | 4(4%) | 2(4.7%) | 7(7.69%) | 28(11.2%) B* | NR | NR | NR | NR | 205/1328 (15.4%) |
| **Hemoptysis** | NR | NR | NR | NR | NR | NR | NR | NR | NR | 16/1315(1.2%) |
| **Chills** | 5(14.29%) | 6(7%) | NR | NR | NR | NR | NR | 23(9.8%) | NR | 163/1333 (12.2%) |
| **Chest pain** | NR | NR | NR | NR | NR | NR | NR | 8(3.4%) | NR | NR |
| **Chest distress** | 11(31.43%) | NR | | 17(18.68) | NR | NR | NR | 13(5.6%) | 6(6.9%) | NR |
| **Diarrhea** | 9(25.71%) | 5(6%) | 6(14.2%) | 21(23.08%) | 8(3.2%) | NR | NR | 9(3.8%) | 4(4.6%) | 57/1359 (4.2%) |
| **Inappetence** | 6(17.14%) | NR | NR | Anorexia: 23(25.27%) | 8(3.2%) | NR | 20(27.4%) | NR | NR | NR |

| | | | | | | | | | | |
|---|---|---|---|---|---|---|---|---|---|---|
| **Abdominal pain** | NR | NR | 1(2.3%) | NR | NR | NR | NR | NR | Abdominal distention/ pain: 3(3.5%) | NR |
| **Sore throat** | 10(28.57%) | 23(26%) | 4(9.5%) | NR | 16(6.4%) | 1(9%) | NR | NR | 6(6.9%) | 194/1317 (14.7%) |
| **Rhinorrhea** | NR | NR | NR | NR | 17(6.8%) | 0 | NR | 13(5.6%) | 3(3.5%) | NR |
| **Nausea or vomiting** | NR | Nausea: 5(6%); Vomiting: 2(2%) | Nausea: 4(9.5%) | Nausea: 11(12.09%); Vomiting: 6(6.59%) | NR | NR | NR | Nausea and vomiting: 5(2.1%) | NR | 80/1371 (5.8%) |
| **Asymptomatic infection** | NR | 6(7%) | NR | NR | 7(2.8%) | NR | NR | NR | 2(2.3%) | 73(4.6%) |
| **Others** | Constipation: 2(5.71%); Bitter taste: 4(11.43%); Thirst: 2(5.71%). | | | Back discomfort: 3(3.3) | | | | pharyngeal discomfort: 35(15%). | Dry throat: 6(6.9%). | Unconsciousness: 20/1421 (1.4%); Conjunctival congestion: 10/1345 (0.7%); Tonsil swelling: 31/1376 (2.3%); Lymphadenectasis: 2/1375 (0.1%); Rash: 3/1378 (0.2%); Throat congestion: 21/1286 (1.6%). |

Continuous data are median(IQR) or mean(SD); Categorical data are n(N%). NR: not reported. A*: White phlegm 8(22.86%), yellow sputum 9(25.71%). B*: Dizziness and headache.

#### Table S19 Signs and symptoms of the confirmed patients outside of former epicenter c

| | Feng 2020 | Xu 2020 D | Wan 2020 | Li 2020 B | Wu 2020 B | Tang 2020 | Zheng 2020 | Zhao 2020 | Lo 2020 | Yu 2020 A |
|---|---|---|---|---|---|---|---|---|---|---|
| **Fever** | 390/454(85.9%) | 34(66.7%) | 120(88.9%) | 72(86.7%) | 61(76%) | 72(86.7%) | 122(75.8%) | 79(78.2%) | 8(80%) | 18(72%) |
| **Highest temperature ＞39℃** | NR | NR | 7(5.1%) | NR | 3(4%) | NR | NR | NR | NR | NR |
| **Highest temperature ＜37.5℃** | NR | < 38℃: 18(35.3%) | < 37.3℃: 16(11.9%) | NR | < 37.3℃: 19(24%) | NR | NR | NR | NR | NR |
| **Cough** | Dry cough: 269/453(59.4%) | 23(45.1%) | 102(76.5%) | 65(78.3%) | 58(73%) | 60(72.3) | 101(62.7%) | 63(62.4%) | 5(50%) | Dry cough: 21(84%) |
| **Sputum production** | 161/453(35.5%) | 13(25.5%) | 12(8.8%) | 15(18.1%) | 11(14%) | 34(41.0) | NR | NR | NR | 4(16%) |
| **Dyspnea** | 109/447(24.4%) | 4(7.8%) | 18(13.3%) | 9(10.8%) | 7(9%) | 7(8.4%) | 23(14.3%) | 1(1.0%) | 5(50%) | NR |
| **Shortness of breath** | NR | NR | Chest tightness and shortness of breath: 12(8.8%) | NR | NR | 19(22.9) | NR | NR | NR | NR |
| **Fatigue** | NR | 2(3.9%) | Fatigue or myalgia: 44(32.5%) | NR | NR | 25(30.1) | 64(39.8%) | Fatigue or myalgia: 17(16.8%) | NR | 7(28%) |
| **Myalgia** | 55/438(12.6%) | 8(15.7%) | | 15(18.1%) | 13(16%) | 12(14.5) | 18(11.2%) | | 3(30%) | 6(24%) |
| **Headache** | NR | NR | 34(32.5%) | 9(10.8%) | Dizziness and headache: 8(10%) | 12(14.5) | 12(7.5%) | NR | NR | NR |
| **Dizziness** | NR | NR | NR | NR | NR | 5(6.0%) | NR | NR | 2(20%) | NR |
| **Hemoptysis** | 5/435(1.1%) | NR | 4(3%) | NR | NR | 1(1.2) | NR | NR | NR | NR |
| **Chills** | 24/374(6.4%) | NR | 14(10.3%) | NR | NR | 9(10.8) | NR | NR | NR | NR |
| **Chest pain** | 21/440(4.8%) | NR | NR | 5(6%) | 5(6%) | NR | NR | NR | NR | NR |
| **Diarrhea** | NR | 5(9.8%) | 18(13.3%) | Abdominal pain and | Abdominal pain and | 5(6.0) | 17(10.6%) | 3(3.0%) | 8(80%) | 1(4%) |

| | | | | | | | | | | |
|---|---|---|---|---|---|---|---|---|---|---|
| | | | | diarrhea: 7(8.4%) | diarrhea: 7(9%) | | | | | |
| **Inappetence** | NR | NR | 6(4.4%) | NR | NR | 6(7.2%) | NR | NR | NR | NR |
| **Abdominal pain** | NR | NR | NR | NR | NR | 1(1.2%) | NR | NR | 2(20%) | NR |
| **Respiratory rate** | NR | NR | > 24/min: 12(8.9%) | 20(20-22) | 21.05(3.74) | NR | NR | NR | NR | NR |
| **Sore throat** | 35/433(8.1%) | 3(5.9%) | 24(17.7%) | NR | NR | 14(16.9) | NR | 12(11.9%) | 5(50%) | 3(12%) |
| **Pharyngeal discomfort** | NR | NR | NR | 6(7.2%) | 9(11%) | NR | NR | NR | NR | NR |
| **Rhinorrhea** | NR | NR | NR | NR | NR | NR | 0 | NR | 2(20%) | NR |
| **Nausea or vomiting** | NR | NR | Retching: 4(3.0%) | NR | NR | 4(4.8) | Nausea: 6(3.7%) | 2(2.0%) | Nausea: 5(50%) | NR |
| **Others** | Digestive symptoms: 49/446(11%); Neurological symptoms: 47/440(10.7%). | | Palpitation 5(3.7%). | | Blood in sputum 3(4%); | Catarrh symptoms: 7(8.4); Thirst: 2(2.4); Chest distress: 8(9.6%) | NR | More than one symptom: 67(66.3%); Asymptomatic infection: 2(2.0%). | Nasal congestion: 2(20%). | |

Continuous data are median(IQR) or mean(SD); Categorical data are n(N%). NR: not reported.

#### Table S20 Signs and symptoms on admission of the confirmed patients outside of former epicenter d

| | Diao 2020 | Gao 2020 | Li 2020 C | Liang 2020 B | Lv 2020 | Mao 2020 B | Wang 2020 D | Xiang 2020 | Xu 2020 E |
|---|---|---|---|---|---|---|---|---|---|
| **Fever** | 8(53.3%) | 30(75%) | 47(95.9%) | 26(92.9%) | 17(100%) | 15(62.5%) | 16(94.1%) | 38(78.7%) | 27(84.3%) |
| **Highest temperature＞39℃** | NR | NR | NR | 4(14.3%) | NR | 1(4.2%) | NR | NR | NR |
| **Cough** | 10(66.7%) | Dry cough: 3(7.5%) | 37(75.5%) | 14(50%) | 15(88.2%) | 22(91.66%); Dry cough: 9(37.5%) | Dry cough: 7(41.2%) | 19(38.8%) | 19(59.4%) |
| **Sputum production** | 1(6.7%) | NR | 19(38.8%) | 5(17.9%) | NR | 13(54.2%) A* | 1(5.9%) | 7(14.3%) | NR |
| **Dyspnea** | NR | NR | NR | 3(10.7%) | 4(23.5%) | 6(25%) | 4(23.5%) | NR | 4(12.5%) |
| **Fatigue** | NR | NR | 32(65.3%) | 15(53.6%) | 13(76.4%) | 11(45.83%) | 2(11.8%) | 9(18.4%) | 5(15.6%) |
| **Myalgia** | NR | NR | NR | 7(25%) | 1(5.9%) | NR | 1(5.9%) | NR | 12(37.5%) |
| **Headache** | 1(6.7%) | 1(2.5%) | NR | NR | NR | 2(8.33%) | NR | 6(12.2%) | 4(12.5%) |
| **Dizziness** | NR | NR | NR | NR | NR | 1(4.16%) | 1(5.9%) | | NR |
| **Chills** | 2(13.3%) | NR | NR | NR | NR | 1(4.16%) | 2(11.8%) | 8(16.3%) | 7(21.9%) |
| **Chest pain** | NR | NR | NR | NR | NR | NR | NR | NR | NR |
| **Chest distress** | 2(13.3%) | NR | Chess distress and shortness of breath 31(63.2%) | 9(32.1%) | NR | 6(25%) | NR | 4(8.2%) | NR |
| **Diarrhea** | NR | NR | 4(8.2%) | 1(3.6%) | NR | 1(4.16%) | 1(5.9%) | 2(4.1%) | 0 |
| **Respiratory rate** | 19.0(18.0 ~ 20.0) | NR | NR | NR | NR | NR | NR | 20.6(14 ~ 32) | NR |
| **Sore throat** | 1(6.7%) | 2(5%) | NR | 2(7.1%) | NR | 3(12.5%) | 2(11.8%) | Sore throat/Pharyngeal discomfort: 7(14.9%) | 2(6.3%) |
| **Rhinorrhea** | NR | NR | NR | 1(3.6%) | NR | 4(16.7%) | NR | NR | 1(3.1%) |
| **Nausea or vomiting** | NR | NR | NR | NR | NR | 1(4.16%) | Nausea: 2(11.8%) | NR | NR |
| **Asymptomatic infection** | NR | 4(10%) | NR | NR | NR | NR | NR | 3(6.1%) | NR |
| **Others** | Other digestive symptoms: 1(6.7%); | | | Inappetence: 2(7.1%); Black stool: 2(7.1%) | Thirst: 1(5.9%) | | | | Pneumonia: 31(96.9%); |

Continuous data are median(IQR) or mean(SD); Categorical data are n(N%). NR: not reported. A*: White sputum 10(41.67%), yellow sputum 3(12.5%).

### Table S21 Complications of the confirmed patients in/ outside of former epicenter/ nationwide

| | Huang 2020 A | Chen 2020 A | Liu 2020 A | Wang 2020 | Xu 2020 A | Guan 2020 | Yang 2020 | Chen 2020 C | Wan 2020 | Xu 2020 D | Lo 2020 | Feng 2020 |
|---|---|---|---|---|---|---|---|---|---|---|---|---|
| ARDS | 12(29%) | 17(17%) | | 27(19.6%) | 1(2%) | 37(3.4%) | 0 | 8(3.2%) | 21(15.6%) | 0 | 0 | NR |
| Acute cardiac injury | 5(12%) | NR | | 10(7.2%) | NR | NR | 0 | NR | 10(7.4%) | NR | 0 | NR |
| Acute respiratory injury | NR | 8(8%) | Comorbid organ dysfunction: 26(18.9%), and patients with underlying cardiovascular diseases often demonstrated comorbid heart failure. | NR | NR | NR | 0 | NR | NR | NR | 0 | NR |
| Acute kidney injury | 3(7%) | 3(3%) A* | | 5(3.6%) | NR | 6(0.5%) | 0 | NR | 5(3.7%) | NR | 0 | NR |
| Shock | 3(7%) | 4(4%) | | 12(8.7%) | NR | 12(1.1%) | 0 | NR | 1(0.7%) | NR | 0 | NR |
| Secondary infection | 4(10%) | NR | | NR | NR | NR | NR | NR | 7(17.5%) | NR | NR | 35/410(8.5%) |
| Others | RNAaemia: 6(15%) | Ventilator-associated pneumonia: 1(1%) | | Arrhythmia: 23(16.7%) | | Rhabdomyolysis: 2(0.2%); Disseminated intravascular coagulation: 1(0.1%) | | | | | | |

Categorical data are n(N%). ARDS, acute respiratory distress syndrome. A*: With renal function damage 7(7%). B*: Cycle threshold of RNAaemia was 35.1(34.7–35.1) median(IQR).

### Table S22 Radiologic findings of the confirmed patients in former epicenter a

| | Huang 2020 | Chen 2020 | Liu 2020 A | Wang 2020 | Huang 2020 B | Chen 2020 B | Zhang 2020 B |
|---|---|---|---|---|---|---|---|
| **Bilateral involvement of chest radiological examination on admission** | CT: 40 (98%) | CT and x-ray: 74 (75%) | CT: 116 (84·7%) | CT: 138 (100%) | 27 (79·4%) | C* | CT: 121/135 (89·6%) |
| **Local patchy shadowing of chest radiological examination on admission** | NR | CT and x-ray: 25 (25%) | CT:36 (31·0%) | CT: 0 | 3 (8·8%) | | 13/135 (9·6%) |
| **Ground glass opacity of chest radiological examination on admission** | A* | CT and x-ray: 14 (14%) | CT:55 (47·4%) | CT: 138 (100%) B* | NR | | NR |
| **Consolidation shadow of chest radiological examination on admission** | D* | NR | CT:25 (21·6%) | NR | NR | | NR |
| **No abnormality of chest radiological examination on admission** | CT: 0 | CT and x-ray: 0 | NR | NR | 1 (2·9%) | NR | CT: 1/135 (0·7%) |

Categorical data are n (N%). NR: not reported. A*: Huang2020 reported that most patients presented with bilateral ground-glass opacities on chest CT scans, but no certain frequency reported. B*: Ground glass opacity or patchy shadows with bilateral involvement. C*:Typical signs reported including: (1) local ground glass shadow; (2) multiple peripheral ground glass shadow; (3) patchy GGO with segmental consolidation; (4) paving stone sign; (5) diffuse ground glass shadow with bronchial sign; (6) large area consolidation shadow with interlobular thickening; D*: The typical findings of chest CT images of ICU patients on admission.

#### Table S23 Radiologic findings of the confirmed patients in former epicenter b

| | Cao 2020 | Li 2020 A | Wang 2020 B | Chen 2020 D | Din 2020 | Wang 2020 C |
|---|---|---|---|---|---|---|
| **Bilateral involvement of chest radiological examination on admission** | CT and x-ray: 72(70.6%) | CT A* | CT B* | CT: 60(93.8%) | HRCT: 53(94.6%) | CT: 89(92.7%) |
| **Local patchy shadowing of chest radiological examination on admission** | CT and x-ray: 30(29.4%) | | CT C* | CT: 4(6.2%) | HRCT: 2(3.6%) | CT: 7(7.3%) |
| **Ground glass opacity of chest radiological examination on admission** | CT and x-ray: 18(17.6%) | | CT: 30(27.3%) | CT: 48(75%) | NR | NR |
| **Consolidation shadow of chest radiological examination on admission** | NR | | CT: 30(27.3%) | D* | NR | NR |
| **No abnormality of chest radiological examination on admission** | NR | NR | CT: 3(2.6%) | NR | NR | NR |
| **Others** | | | | | Pericardial effusion: 8(14.3%); Mediastinal emphysema 2(3.6%); Pulmonary embolism 1(1.8%). | Bilateral pleural effusion (CT): 5(5.2%) |

Categorical data are n(N%). NR: not reported. A*: Mild cases: single or multiple ground glass opacity(GGO) mainly in the dorsal or periphery part of the lung, local or bilateral patchy shadowing for most common cases/ Severe cases: rapidly increased size and density of lesions, and diffused damage covered both lungs, with air bronchogram and interstitial abnormalities, and “White lung” predicted very poor prognosis. B*: Multiple lobes of both lungs 80(72.7%), bilateral lower lungs 4(3.6%), bilateral middle and lower lungs 10(9.1%). C*: Single lobe of one lung 10(9.1%), multiple lobes of one lung 6(5.5%), patchy 56(50.1%), spherical 28(25.5%). D*: Patchy consolidation 7(10.9%), Large scale consolidation 8(12.5%).

#### Table S24 Radiologic findings of the confirmed patients outside of former epicenter/ nationwide a

| | Xu 2020 A | Guan 2020 | Liu2020 B | Yang 2020 | Wu 2020 A | Xu 2020 B |
|---|---|---|---|---|---|---|
| **Bilateral involvement of chest radiological examination on admission** | CT and x-ray: 52 (84%) | CT: 505/975 (51·8%); X-ray: 100/274 (36·5%) | A* | B* | CT: 36 (45·00%) | D* |
| **Local patchy shadowing of chest radiological examination on admission** | NR | CT: 409/975 (41·9%); X-ray: 77/274 (28·1%) | | | CT: 19 (23·75%) | E* |
| **Ground glass opacity of chest radiological examination on admission** | NR | CT: 550/975 (56·4%); X-ray: 55/274 (20·1%) | | | NR | 21 (75·0%) |
| **Interstitial abnormalities of chest radiological examination on admission** | NR | CT: 143/975 (14·7%); X-ray: 12/274 (4·4%) | | | NR | NR |
| **No abnormality of chest radiological examination on admission** | CT and x-ray: 1 (2%) | CT: 135/ 975 (13·8%)； X-ray: 112/274 (40·9%) | NR | 17 (11·4%) | CT: 25 (31·25%) | NR |

Categorical data are n (N%). NR: not reported. A*: Typical findings as bilateral multiple patchy ground glass and consolidation shadows. B*: Finding were reported in the total number of pulmonary segments. C*: 8 (47·1%) were patchy shadows or ground glass opacity; D*:15(53·6%) with bilateral upper lobes involved, and 12(42·9%) with bilateral lower lobes involved. E*: 30 (73·2%) with right upper lobe involved; 22 (53·7%) with right middle lobe involved; 39 (95·1%) with right lower lobe involved; 33 (80·5%) with left upper lobe involved; 36 (87·8%) with left lower lobe involved.

### Table S25 Radiologic findings of the confirmed patients outside of former epicenter b

| | Huang 2020 C | Xu 2020 C | Liu 2020 D | Qian 2020 | Chen 2020 C | Peng 2020 | Liu 2020 C | Dai 2020 | Xu2020 D | Wan 2020 |
|---|---|---|---|---|---|---|---|---|---|---|
| **Bilateral involvement of chest radiological examination on admission** | CT: 21(60%) | CT: 53(59%) | | CT: 61(67.03%) | CT or X-Ray: 203(81.5%) | | | CT: 192/219(87.7%) | CT: 39(76%) | CT: 135(100%) |
| **Local patchy opacity of chest radiological examination on admission** | CT: 19(54.28%) | CT: U* | B* | CT: 25(27.47%) | CT or X-Ray: 39(15.7%) | D* | E* | F* | CT: 8(15.7%), Unilateral pneumonia | NR |
| **Ground glass opacity of chest radiological examination on admission** | CT: 11(31.43%) A* | CT: 65(72%) | | C* | NR | | | NR | CT: 4(7.8%) G* | NR |
| **Consolidation** | NR | CT: 12(13%) | NR | NR | NR | NR | CT: 8(11%) | NR | NR | NR |
| **Crazy-paving pattern** | NR | CT: 11(12%) | NR | NR | NR | NR | CT: 28(38.4%) | NR | NR | NR |
| **Bronchial wall thickening** | NR | NR | NR | NR | NR | NR | CT: 19(26%) | HRCT: 76/219(34.7%) | NR | NR |
| **Interlobular septal thickening** | NR | CT: 33(37%) | NR | NR | NR | NR | NR | HRCT: 205/219(93.6%) | NR | NR |
| **Pleural effusion** | NR | 4(4%) | NR | NR | NR | NR | 3(4.1%) | NR | NR | NR |
| **Pleural thickening** | NR | Adjacent pleura thickening, CT: 50(56%) | NR | NR | NR | NR | NR | HRCT: 170/219(77.6%) | NR | NR |
| **Air bronchogram sign** | NR | CT: 7(8%) | NR | NR | NR | NR | NR | HRCT: 184/219(84%) | NR | NR |
| **No abnormality of chest radiological examination on admission** | CT: 4(11.43%) | CT: 21(23.3%) | NR | CT: 5(5.49%) | CT and X-Ray: 6(2.41%) | NR | CT: 3(4.1%) | HRCT: 15(6.4%) | NR | 0 |
| **Others** | | CT: H* | | | | | | HRCT: I* | | |

Categorical data are n(N%). NR: not reported. A*: And cloud opacity 3(8.57%). U*: Periphery distribution 46(51%), multifocal involvement 62(69%), unifocal involvement 7(8%). B*: Bilateral involvement for most patients, and the ground glass opacity appeared in the outer zone of both lungs in the early stage. The density opacity of the two upper lobes is scattered in the flake ground glass, and the edge is fuzzy for mild patients on admission. Bilateral extrapulmonary bands are scattered in patchy, flocculent high-density opacities, with fuzzy edges and local pulmonary tissue consolidation for patients with severe condition on admission. For critical patients, bilateral lungs presented cord like fibrosis, fuzzy margin, distorted bronchi and local stenosis during treatment. C*: Patchy ground-glass opacities in lung as typical findings. D*: Were mainly patchy ground-glass opacities(GGO) and consolidation opacities distributed along the bronchovascular bundles and subpleural space, with a thickened leaflet interval and thickened lobular septum, showing "crazy-paving pattern" and vascular bundle sign. E*: Unilateral lung involvement 15(10.5%), bilateral lungs involvement 55(75.3%), unique ground-glass opacities 15(10.5%), multiple ground-glass opacities 50(68.5%), thickening of lung texture 65(89%). F*: Single lobe involved 16/219(7.31%), distributed in the lower lungs and/or the periphery of the lungs mainly 208/219(94.98%), irregular shape of the lesions mainly 193/219(88.13%), followed by small patches 189/219(86.3%), strip-like 152/219(69.41%), round-like 108/219(49.32%) and "anti-butterfly" 105/219(47.95%,). G*: Multiple mottling and ground-glass opacity. H*: Linear opacities combined: 55(61%), cavitation: 0; pericardial effusion: 1(1%); lymphadenopathy: 1(1%); pulmonary emphysema: 0. I*: With VES 207/219(94.5%), with intralesional and/or perilesional bronchiectasis 173/219(79%), with solid nodules 138/219(63%), with reticular/mosaic sign 135/219(61.6%), with interlobar fissure displacement 124/219(56.6%), with minor pleural effusion and pericardial effusion 29/219(13.2%), with mediastinal lymphadenopathy 21/219(9.6%).

#### Table S26 Radiologic findings of the confirmed patients outside of former epicenter c

| | Li 2020 B | Wu 2020 B | Tang 2020 | Zheng 2020 | Zhao 2020 | Lo 2020 | Feng 2020 | Diao 2020 | Gao 2020 | Li 2020 C |
|---|---|---|---|---|---|---|---|---|---|---|
| **Bilateral involvement of chest radiological examination on admission** | CT: 79(95.2%) | CT: 53(59%) | CT: 70/75(93.3%) | CT: 89(55.3%) | CT: 83(82.2%) | CT: 8(80%) | CT: 373/442(84.4%) | HRCT: 9(60%) | CT: U* | CT: 34(69.4%) |
| **Local patchy opacity of chest radiological examination on admission** | NR | NR | NR | CT: 113(70.2%) | NR | A* | NR | HRCT: 6(40%) | CT: 12(30%) | CT: 15(30.6%) |
| **Ground glass opacity of chest radiological examination on admission** | CT: 81(97.6%) | CT: 73(91%) | CT: 74/75(98.7%) | CT: 82(50.9%) | CT: 87(86.1%) | | CT: 425/442(96.2%) | HRCT: 14(93.3%) | CT: U* | NR |
| **Interstitial abnormalities of chest radiological examination on admission** | NR | NR | CT: 8/75(10.7%) | CT: 2(1.2%) | NR | NR | NR | NR | NR | NR |
| **Consolidation** | CT: 53(63.9%) | CT: 50(63%) | CT: 34/75(45.3%) | NR | CT: 44(43.6%) | NR | CT: 87/442(19.7%) | HRCT: 6(40%) | NR | NR |
| **Bronchial wall thickening** | CT: 19(22.9%) | CT: 9(11%) | NR | NR | CT: 29(28.7) | NR | NR | NR | NR | NR |
| **Interlobular septal thickening** | CT: 52(62.7%) | CT: 47(59%) | CT: 36/75(48%) | NR | NR | NR | NR | NR | NR | NR |
| **Pleural effusion** | NR | CT: 5(6%) | CT: 3/75(4%) | NR | CT: 14(13.9%) | NR | CT: 25/442(5.7%) | NR | NR | NR |
| **Lymph node enlargement** | NR | CT: 3(4%) | Mediastinal lymph node, CT: 8/75(10.7%) | NR | Intrathoracic lymph node, CT: 1(1.0) | NR | NR | Mediastinal lymph node, HRCT:3(20%) | NR | NR |
| **Crazy-paving pattern** | CT: 30(36.1%) | CT: 23(29%) | NR | NR | NR | NR | NR | NR | NR | NR |
| **Pleural thickening** | NR | NR | Interlobular pleural, CT: 2/75(2.7%) | NR | NR | NR | CT: 238/442(53.8%) | NR | NR | NR |
| **No abnormality of chest** | CT: | CT: 4(5%) | 0 | NR | CT: 8(7.9%) | CT: | NR | NR | CT: | 0 |

| | | | | | | | |
|---|---|---|---|---|---|---|---|
| **radiological examination on admission** | 4(11.43%) | | | 2(20%) | | | 3(7.5%) |
| **Others** | CT: B* | CT: C* | CT: D* | CT: E* | CT: linear opacity: 129/442(29.2%). | HRCT: fibrotic lesion 3(20%). | |

Categorical data are n(N%). NR: not reported. A*: Bilateral distribution of patchy opacities or ground glass opacity 6(60%). B*: 54(65.1%) for linear opacities, 21(25.3%) for spider web sign, 17(20.5%) for subpleural curvilinear. C*: Spider web sign 20(25%), subpleural line 16(20%), pericardial effusion 4(5%), subpleural distribution 42(53%), diffuse distribution 7(9%), peribronchial distribution 3(4%), mixed distribution 24(30%). D*: patchy opacity 72/75(96%), flake opacity 46/75(61.3%), nodular opacity 11/75(14.7%), stripe opacity 51/75(68%), subpleural line 27/75(36%), gridding opacity11/75(14.7%), mixing density 34/75(45.3%), air bronchogram 23/75(30.7%), thickening of blood vessels in ground glass opacity 5/75(6.7%), bronchiectasia 11/75(14.7%),. E*: Mixed ground-glass opacities and consolidation 65(64.4), centrilobular nodules 23(22.8), architectural distortion 22(21.8), reticulation 49(48.5), subpleural bands 28(27.7) traction bronchiectasis 53(52.5), vascular enlargement 72(71.3). U*: subpleural strip opacity and ground glass opacities 3(7.5%), bilateral gridding and ground glass opacities 5(12.5%), bilateral multiple ground glass opacities 17(42.5%).

### Table S27 Radiologic findings of the confirmed patients outside of former epicenter d

| | Liang 2020 B | Lv 2020 | Mao 2020 B | Wang 2020 D | Xiang 2020 | Xu 2020 E | Yu 2020 A | Yu 2020 B |
|---|---|---|---|---|---|---|---|---|
| **Bilateral involvement of chest radiological examination on admission** | | | | CT: 14(82.4%) | | CT: 26(81.3%) | CT: F* | NR |
| **Local patchy opacity of chest radiological examination on admission** | CT(Typical radiologic findings of COVID-19): 28(100%) | CT: A* | CT: B* | CT: C* | CT: D* | CT: E* | | NR |
| **Ground glass opacity of chest radiological examination on admission** | | | | | | CT: 26(80.3%) | NR | NR |
| **Consolidation** | NR | NR | NR | NR | NR | CT: 14(43.8%) | NR | NR |
| **Pleural effusion** | NR | NR | NR | NR | NR | CT: 4(12.5%) | NR | NR |
| **Pleural thickening** | NR | NR | NR | NR | NR | CT: 7(21.9%) | NR | NR |
| **No abnormality of chest radiological examination on admission** | CT: 0 | CT: 1(5.9%) | NR | CT: 3(17.6%) | CT: 2(4.1%) | CT: 1(3.1%) | CT: 18(72%) | CT: 9(9.3%) |

Categorical data are n(N%). NR: not reported. A*: Ground glass opacity, mixed ground glass opacity and crazy-paving pattern mainly. B*: Bilateral ground glass opacity 22(91.67%), bilateral ground glass opacity with unliteral pleural effusion 2(8.33%), bilateral ground glass opacity with slight pericardial effusion 1(4.17%), lung texture increased and thickened 1(4.17%). C*: Reported as bilateral patchy, ground glass opacity, pulmonary consolidation and interstitial lesions mainly; D*: Single or scattered patchy ground glass density opacitys 47(95.9%), and the lesions involved a wide range with an average of 2.8 lobes. E*: Patchy opacity 27(84.6%), nodular opacity 8(23.1%), sector opacity 12(36.5%), irregular opacity 15(48.1%), ground glass opacity with consolidation 20(62.5%), tubercle opacity 7(21.9%), interlobular septal thickening 23(71.9%), bronchovesicular bundle thickening 22(68.8%), crazy-paving pattern 14(43.8%), air bronchogram 12(37.5), cavity 9(28.1%), halo sign 8(25.0%), reversed halo sign 6(18.8%), tree-in-bud 4(12.5%), fibrosis 6(18.8%), “White lung”1(3.1%), mediastinal lymphadenopathy 1(3.1%). F*: Patchy exudative opacities and interstitial abnormalities 7(28%), bilateral pneumonia 2(8%), unilateral pneumonia 5(20%).

#### Table S28 Laboratory findings of the confirmed patients in former epicenter a

| | Huang 2020 A | Chen 2020 A | Liu 2020 A | Wang 2020 A | Huang 2020 B | Chen 2020 B | Zhang 2020 B | Mao 2020 A |
|---|---|---|---|---|---|---|---|---|
| **White blood cell count, × $10^9$/L** | 6·2 (4·1–10·5) | 7·5 (3·6) | NR | 4·5 (3·3-6·2) | NR | NR | 4·7 (3·7-6·7) | 4·9 (0·1-20·4) |
| **White blood cell count ＞ 10 × $10^9$/L** | 12/40 (30%) | 24 (24%) | 26 (19·0%) | NR | Increased: 4 (11·8%) | 6 (21%) | ＞ 9·5 × $10^9$/L: 17/138 (12·3) | |
| **White blood cell count ＜ 4× $10^9$/L** | 10/40 (25%) | 9 (9%) | 51 (37·2%) | NR | Decreased: 6 (17·6%) | 6 (21%) | ＜ 3·5× $10^9$/L: 27/138 (19·6%) | |
| **Neutrophil count, × $10^9$/L** | 5·0 (3·3–8·9) | 5·0 (3·3–8·1) | NR | 3·0 (2·0-4·9) | Increased: 4 (11·8%) | NR | NR | 3·0 (0·0-18·7) |
| **Monocyte count, $10^9$/L** | NR | > 6·3 × $10^9$/L: 38 (38%) | NR | 0·4 (0·3-0·5) | NR | NR | NR | NR |
| **Lymphocyte count, × $10^9$/L** | 0·8 (0·6–1·1) | NR | NR | 0·8 (0·6-1·1) | NR | NR | 0·8 (0·6-1·1) | 1·1 (0·1-2·6) |
| | < 1× $10^9$/L: 26 (63%) | < 1·1× $10^9$/L: 35 (35%) | < 1× $10^9$/L: 99 (72·3%) | | Decreased: 17 (50·0%) | < 1× $10^9$/L: 6 (21%) | < 1× $10^9$/L: 104/138 (75·4%) | |
| **Haemoglobin, g/L** | 126·0 (118·0–140·0) | 129·8 (14·8) | NR | NR | NR | NR | NR | NR |
| | | < 130g/L: 50 (51%) | | | Decreased: 13 (38·2%) | < 130g/L: 12(41%) | | |
| **Platelet count, × $10^9$/L** | 164·5 (131·5–263·0) | 213·5 (79·1) | NR | 163 (123-191) | NR | NR | NR | 209·0 (18·0-583·0) |
| | < 100× $10^9$/L: 2/40 (5%) | < 125× $10^9$/L: 12 (12%) | | | Decreased: 9 (26·5%) | < 125× $10^9$/L: 5(17%) | | |
| | | > 350× | | | | | | |

| | | | | | | | | |
|---|---|---|---|---|---|---|---|---|
| | | $10^9$/L: 4 (4%) | | | | | | |
| **Prothrombin time, s** | 11·1 (10·1–12·4) | 11·3 (1·9) | NR | 13·0 (12·3-13·7) | NR | NR | NR | NR |
| | | < 10·5 s: 30 (30%) | | | Decreased: 1 (2·9%) | | | |
| | | > 13·5 s: 5 (5%) | | | Increased: 17 (50·0%) | | | |
| **Activated partial thromboplastin time, s** | 27·0 (24·2–34·1) | 27·3 (10·2) | NR | 31·4 (29·4-33·5) | NR | NR | NR | NR |
| | | < 21 s: 16 (16%) | | | | | | |
| | | > 37 s: 6 (6%) | | | | | | |
| **D-dimer, mg/L** | 0·5 (0·3–1·3) | 0·9 (0·5–2·8) | NR | 203 (121-403) μg/L | NR | NR | 0·2 (0·1-0·5) | 0·5 (0·1-20·0) |
| | | > 1·5mg/L: 36 (36%) | | | Increased: 5 (14·7%) | | > 0·243mg/L: 35/81 (43·2%) | |
| **Albumin, g/L** | 31·4 (28·9–36·0) | 31·6 (4·0) | NR | NR | NR | NR | NR | NR |
| | | < 40g/L: 97 (98%) | | | Decreased: 25 (73·5%) | < 35g/L: 15(52%) | | |
| **Alanine aminotransferase, U/L** | 32·0 (21·0–50·0) | 39·0 (22·0–53·0) | NR | 24 (16-40) | NR | NR | NR | 26·0 (5·0-1933·0) |
| | | > 50 U/L: 28 (28%) | | | Increased: 8 (23·5%) | > 41 U/L: 5(17%) | | |
| **Aspartate aminotransferase, U/L** | 34·0 (26·0–48·0) | 34·0 (26·0–48·0) | NR | 31 (24-51) | NR | NR | NR | 26·0 (8·0-8191·0) |
| | > 40 U/L: 15 (37%) | > 40 U/L: 35 (35%) | | | Increased: 7 (20·6%) | > 40 U/L: 7(24%) | | |
| **Total bilirubin, mmol/L** | 11·7 (9·5–13·9) | 15·1 (7·3) | NR | 9·8 (8·4-14·1) | NR | NR | NR | NR |

| | | | | | | | | |
|---|---|---|---|---|---|---|---|---|
| | | > 21 mmol/L: 18 (18%) | | | Increased: 3 (8·8%) | > 26 mmol/L: 1(3%) | | |
| **Potassium, mmol/L** | 4·2 (3·8–4·8) | NR | NR | NR | Decreased: 4 (11·8%); Increased: 2 (5·9%) | NR | NR | NR |
| **Sodium, mmol/L** | 139·0 (137·0–140·0) | NR | NR | NR | NR | NR | NR | NR |
| **Serum creatinine, µmol/L** | 74·2 (57·5–85·7) | 75·6 (25·0) | NR | 72 (60-87) | NR | NR | NR | 68·2 (35·9-9435·0) |
| | > 133 µmol/L: 4 (10%) | > 111 µmol/L: 3 (3%) | | | Increased: 6 (17·6%) | > 104 µmol/L: 2(7%) | | |
| | | < 57 µmol/L: 51 (52%) | | | Decreased: 7 (20·6%) | | | |
| **Creatine kinase–MB, U/L** | NR | NR | NR | 14 (10-18) | NR | NR | NR | NR |
| **Creatine kinase, U/L** | 132·5 (62·0–219·0) | 85·0 (51·0–184·0) | NR | 92 (56-130) | NR | NR | 72·5 (52·2-115) | 64·0 (8·8-12216·0) |
| | > 185 U/L: 13/40 (33%) | > 310 U/L: 13 (13%) | | | Increased: 1/12 (8·3%) | | > 200 U/L: 4/60 (6·7) | |
| | ≤185 U/L: 27/40 (68%) | < 50 U/L: 23 (23%) | | | | | | |
| **Lactate dehydrogenase, U/L** | 286·0 (242·0–408·0) | 336·0 (260·0–447·0) | NR | 261 (182-403) | NR | NR | NR | 241·5 (2·2-908·0) |
| | > 245 U/L: 29/40 (73%) | > 250 U/L: 75 (76%) | | | | > 225 U/L: 20(69%) | | |
| **Hypersensitive troponin I, pg/mL** | 3·4 (1·1–9·1) | NR | NR | 6·4 (2·8-18·5) | NR | NR | NR | NR |
| | > 28 pg/mL | | | | Increased: 1/15 (6·7%) | | | |

| | | | | | | | | |
|---|---|---|---|---|---|---|---|---|
| | (99th percentile): 5 (12%) | | | | | | | |
| **Procalcitonin, ng/mL** | 0·1 (0·1–0·1) | 0·5 (1·1) | NR | NR | NR | NR | 0·07 (0·04-0·1) | NR |
| | ⩾0·5 ng/mL: 3/39 (8%)<br>< 0·1 ng/mL: 27/39 (69%) | > 5·0 ng/mL: 6 (6%) | | ⩾0·5 ng/mL: 49 (35·5%) | Increased: 13/31 (41·9%) | | > 0·1 ng/mL: 41/118 (34·7%) | |
| **Blood urea nitrogen, mmol/L** | NR | 5·9 (2·6) | NR | 4·4 (3·4-5·8) | NR | NR | NR | 4·1 (1·5-48·1) |
| | | > 9·5 mmol/L: 6 (6%)<br>< 3·6 mmol/L: 17 (17%) | | | | > 8 mmol/L: 5(17%) | | |
| **C-reactive protein, mg/L** | NR | 51·4 (41·8) | NR | NR | NR | NR | 34·2 (12·5-67·4) | 12·2 (0·1-212·0) |
| | | > 5 mg/L: 63/73 (86%) | Elevated: 115 (83·9%) | | | ⩾5·0 mg/L: 27(93%) | > 3 mg/L: 125/136 (91·9%) | |
| **Others** | NR | NR | NR | NR | Myoglobin: Increased 3/12 (25·0%); Erythrocyte sedimentation rate (ESR): Increased 13/22 (59·1%); Interleukin Increased 9/9 (100·0%); Brain natriuretic peptide (BNP): Increased 1/4 (25·0%) | NR | Lymphocyte percentage, %: 16·9 (9·2-26·0); Eosinophils, $\times 10^9$/L: 0·01 (0·0-0·05), decreased: 73/138 (52·9); Serum amyloid A (SAA), mg/L: 92·53 (44·6-161·3), increased: 46/51 (90·2%) | NR |

Continuous data are median (IQR) or mean (SD); Categorical data are n (N%). NR: not reported.

#### Table S29 Laboratory findings of the confirmed patients in former epicenter b

| | Huang 2020 A | Chen 2020 A | Liu 2020 A | Wang 2020 A | Huang 2020 B | Chen 2020 B | Zhang 2020 B | Mao 2020 A |
|---|---|---|---|---|---|---|---|---|
| **White blood cell count, × $10^9$/L** | 6·2 (4·1–10·5) | 7·5 (3·6) | NR | 4·5 (3·3-6·2) | NR | NR | 4·7 (3·7-6·7) | 4·9 (0·1-20·4) |
| **White blood cell count ＞ 10 × $10^9$/L** | 12/40 (30%) | 24 (24%) | 26 (19·0%) | NR | Increased: 4 (11·8%) | 6 (21%) | ＞ 9·5 × $10^9$/L: 17/138 (12·3) | |
| **White blood cell count ＜ 4× $10^9$/L** | 10/40 (25%) | 9 (9%) | 51 (37·2%) | NR | Decreased: 6 (17·6%) | 6 (21%) | ＜ 3·5× $10^9$/L: 27/138 (19·6%) | |
| **Neutrophil count, × $10^9$/L** | 5·0 (3·3–8·9) | 5·0 (3·3–8·1) | NR | 3·0 (2·0-4·9) | Increased: 4 (11·8%) | NR | NR | 3·0 (0·0-18·7) |
| **Monocyte count, $10^9$/L** | NR | > 6·3 × $10^9$/L: 38 (38%) | NR | 0·4 (0·3-0·5) | NR | NR | NR | NR |
| **Lymphocyte count, × $10^9$/L** | 0·8 (0·6–1·1) | NR | NR | 0·8 (0·6-1·1) | NR | NR | 0·8 (0·6-1·1) | 1·1 (0·1-2·6) |
| | < 1× $10^9$/L: 26 (63%) | < 1·1× $10^9$/L: 35 (35%) | < 1× $10^9$/L: 99 (72·3%) | | Decreased: 17 (50·0%) | < 1× $10^9$/L: 6 (21%) | < 1× $10^9$/L: 104/138 (75·4%) | |
| **Haemoglobin, g/L** | 126·0 (118·0–140·0) | 129·8 (14·8) | NR | NR | NR | NR | NR | NR |
| | | < 130g/L: 50 (51%) | | | Decreased: 13 (38·2%) | < 130g/L: 12(41%) | | |
| **Platelet count, × $10^9$/L** | 164·5 (131·5–263·0) | 213·5 (79·1) | NR | 163 (123-191) | NR | NR | NR | 209·0 (18·0-583·0) |
| | < 100× $10^9$/L: 2/40 (5%) | < 125× $10^9$/L: 12 (12%) | | | Decreased: 9 (26·5%) | < 125× $10^9$/L: 5(17%) | | |
| | | > 350× | | | | | | |

| | | | | | | | | |
|---|---|---|---|---|---|---|---|---|
| | | $10^9$/L: 4 (4%) | | | | | | |
| **Prothrombin time, s** | 11·1 (10·1–12·4) | 11·3 (1·9) | NR | 13·0 (12·3-13·7) | NR | NR | NR | NR |
| | | < 10·5 s: 30 (30%)<br>> 13·5 s: 5 (5%) | | | Decreased: 1 (2·9%)<br>Increased: 17 (50·0%) | | | |
| **Activated partial thromboplastin time, s** | 27·0 (24·2–34·1) | 27·3 (10·2) | NR | 31·4 (29·4-33·5) | NR | NR | NR | NR |
| | | < 21 s: 16 (16%)<br>> 37 s: 6 (6%) | | | | | | |
| **D-dimer, mg/L** | 0·5 (0·3–1·3) | 0·9 (0·5–2·8) | NR | 203 (121-403) μg/L | NR | NR | 0·2 (0·1-0·5) | 0·5 (0·1-20·0) |
| | | > 1·5mg/L: 36 (36%) | | | Increased: 5 (14·7%) | | > 0·243mg/L: 35/81 (43·2%) | |
| **Albumin, g/L** | 31·4 (28·9–36·0) | 31·6 (4·0) | NR | NR | NR | NR | NR | NR |
| | | < 40g/L: 97 (98%) | | | Decreased: 25 (73·5%) | < 35g/L: 15(52%) | | |
| **Alanine aminotransferase, U/L** | 32·0 (21·0–50·0) | 39·0 (22·0–53·0) | NR | 24 (16-40) | NR | NR | NR | 26·0 (5·0-1933·0) |
| | | > 50 U/L: 28 (28%) | | | Increased: 8 (23·5%) | > 41 U/L: 5(17%) | | |
| **Aspartate aminotransferase, U/L** | 34·0 (26·0–48·0) | 34·0 (26·0–48·0) | NR | 31 (24-51) | NR | NR | NR | 26·0 (8·0-8191·0) |
| | > 40 U/L: 15 (37%) | > 40 U/L: 35 (35%) | | | Increased: 7 (20·6%) | > 40 U/L: 7(24%) | | |
| **Total bilirubin, mmol/L** | 11·7 (9·5–13·9) | 15·1 (7·3) | NR | 9·8 (8·4-14·1) | NR | NR | NR | NR |

| | | | | | | | | |
|---|---|---|---|---|---|---|---|---|
| | | > 21 mmol/L: 18 (18%) | | | Increased: 3 (8·8%) | > 26 mmol/L: 1(3%) | | |
| **Potassium, mmol/L** | 4·2 (3·8–4·8) | NR | NR | NR | Decreased: 4 (11·8%); Increased: 2 (5·9%) | NR | NR | NR |
| **Sodium, mmol/L** | 139·0 (137·0–140·0) | NR | NR | NR | NR | NR | NR | NR |
| **Serum creatinine, µmol/L** | 74·2 (57·5–85·7) | 75·6 (25·0) | NR | 72 (60-87) | NR | NR | NR | 68·2 (35·9-9435·0) |
| | > 133 µmol/L: 4 (10%) | > 111 µmol/L: 3 (3%) | | | Increased: 6 (17·6%) | > 104 µmol/L: 2(7%) | | |
| | | < 57 µmol/L: 51 (52%) | | | Decreased: 7 (20·6%) | | | |
| **Creatine kinase–MB, U/L** | NR | NR | NR | 14 (10-18) | NR | NR | NR | NR |
| **Creatine kinase, U/L** | 132·5 (62·0–219·0) | 85·0 (51·0–184·0) | NR | 92 (56-130) | NR | NR | 72·5 (52·2-115) | 64·0 (8·8-12216·0) |
| | > 185 U/L: 13/40 (33%) | > 310 U/L: 13 (13%) | | | Increased: 1/12 (8·3%) | | > 200 U/L: 4/60 (6·7) | |
| | ≤185 U/L: 27/40 (68%) | < 50 U/L: 23 (23%) | | | | | | |
| **Lactate dehydrogenase, U/L** | 286·0 (242·0–408·0) | 336·0 (260·0–447·0) | NR | 261 (182-403) | NR | NR | NR | 241·5 (2·2-908·0) |
| | > 245 U/L: 29/40 (73%) | > 250 U/L: 75 (76%) | | | | > 225 U/L: 20(69%) | | |
| **Hypersensitive troponin I, pg/mL** | 3·4 (1·1–9·1) | NR | NR | 6·4 (2·8-18·5) | NR | NR | NR | NR |
| | > 28 pg/mL | | | | Increased: 1/15 (6·7%) | | | |

| | | | | | | | | |
|---|---|---|---|---|---|---|---|---|
| | (99th percentile): 5 (12%) | | | | | | | |
| **Procalcitonin, ng/mL** | 0·1 (0·1–0·1)<br>⩾0·5 ng/mL: 3/39 (8%)<br>< 0·1 ng/mL: 27/39 (69%) | 0·5 (1·1)<br>> 5·0 ng/mL: 6 (6%) | NR | NR<br>⩾0·5 ng/mL: 49 (35·5%) | NR<br>Increased: 13/31 (41·9%) | NR | 0·07 (0·04-0·1)<br>> 0·1 ng/mL: 41/118 (34·7%) | NR |
| **Blood urea nitrogen, mmol/L** | NR | 5·9 (2·6)<br>> 9·5 mmol/L: 6 (6%)<br>< 3·6 mmol/L: 17 (17%) | NR | 4·4 (3·4-5·8) | NR | NR<br>> 8 mmol/L: 5(17%) | NR | 4·1 (1·5-48·1) |
| **C-reactive protein, mg/L** | NR | 51·4 (41·8)<br>> 5 mg/L: 63/73 (86%) | NR<br>Elevated: 115 (83·9%) | NR | NR | NR<br>⩾5·0 mg/L: 27(93%) | 34·2 (12·5-67·4)<br>> 3 mg/L: 125/136 (91·9%) | 12·2 (0·1-212·0) |
| **Others** | NR | NR | NR | NR | Myoglobin: Increased 3/12 (25·0%); Erythrocyte sedimentation rate (ESR): Increased 13/22 (59·1%); Interleukin Increased 9/9 (100·0%); Brain natriuretic peptide (BNP): Increased 1/4 (25·0%) | NR | Lymphocyte percentage, %: 16·9 (9·2-26·0); Eosinophils, ×$10^9$/L: 0·01 (0·0-0·05), decreased: 73/138 (52·9); Serum amyloid A (SAA), mg/L: 92·53 (44·6-161·3), increased: 46/51 (90·2%) | NR |

Continuous data are median (IQR) or mean (SD); Categorical data are n (N%). NR: not reported.

### Table S30 Laboratory findings of the confirmed patients outside of former epicenter/ nationwide a

| | Xu 2020 A | Chang 2020 | Guan 2020 | Liu 2020 B | Yang 2020 | Wu 2020 A | Xu 2020 B |
|---|---|---|---|---|---|---|---|
| **White blood cell count, × $10^9$/L** | 4.7(3.5-5.8) | 5.83(2.32) | 4.7(3.5– 6.0) | 4.71(3.72-5.77) | 4.56(2.48) | 4.1(3.2-5.7) | NR |
| **White blood cell count ＞ 10 × $10^9$/L** | 1(2%) | | 58/978(5.9%) | | ＞ 9.5 × $10^9$/L: 2(1.34%) | ＞ 9.5 × $10^9$/L: 5(6.25%) | |
| **White blood cell count ＜ 4× $10^9$/L** | 19(31%) | | 330/978(33.7%) | | ＜ 3.5× $10^9$/L: 33(24.16%) | ＜ 3.5× $10^9$/L: 36(45.00%) | Normal or decreased: 49(98%) |
| **Neutrophil count, × $10^9$/L** | 2.9(2.0-3.7) | 3.67(1.71) | NR | 3.29(2.17-4.06) | 2.60(2.03) | 4.3(2.3-5.9) | NR |
| | | | | | ＞ 6.3× $10^9$/L: 6(4.03%)<br>< 1.8× $10^9$/L: 34(22.82%) | ＞ 6.3× $10^9$/L: 20(25.00%) | |
| **Monocyte count, $10^9$/L** | NR | 0.526(0.208) | NR | NR | NR | 0.5(0.3-0.7) | NR |
| | | | | | | > 0.6× $10^9$/L: 15(18.75%) | |
| **Lymphocyte count, × $10^9$/L** | 1.0(0.8-1.5) | 1.58(0.653) | 1.0(0.7– 1.3) | 1.17(0.87-1.35) | 1.21(0.68) | 0.6(0.4-1.0) | NR |
| | < 1× $10^9$/L: 26(42%) | | < 1.5× $10^9$/L: 731/890(82.1%) | | < 1.1× $10^9$/L: 53(35.57%) | < 1.1× $10^9$/L: 26(32.50%) | Decreased: 14(28%) |
| **Hemoglobin, g/L** | 137.0(128.8-152.3) | 147(12.1) | 134.0(119.0–148.0) | NR | NR | 125.3(13.4) | NR |
| | | | | | | < 130g/L: 29(36.25%) | |
| **Platelet count, × $10^9$/L** | 176.0(135.8-215.5) | 199(72.5) | 168.0(132.0–207.0) | NR | 174.5(78.25) | 155(116-188) | NR |
| | < 100× $10^9$/L: 3(5%) | | < 150× $10^9$/L: 315/869(36.2%) | | < 125× $10^9$/L:20(13.42%),<br>> 350× $10^9$/L: 8(5.37%) | < 125× $10^9$/L: 11(13.75%) | |
| **Prothrombin time, s** | NR | NR | NR | NR | 12.20(1.53)<br>> 13.5 s: 17(11.41%)<br>< 10 s: 4(2.68%) | 10.8(9.3-13.2)<br>< 10.5 s: 3(3.75%) | NR |

| | | | | | | | |
|---|---|---|---|---|---|---|---|
| **Activated partial thromboplastin time, s** | NR | NR | NR | NR | 33.29(4.98) | 18.6(17.2-28.6) | NR |
| | | | | | > 36 s: 40(26.85%) | < 21 s: 2(2.50%) | |
| **D-dimer, mg/L** | 0.2(0.2-0.5) | NR | NR | NR | 0.22(0.28) | 0.9(0.4-2.4) | NR |
| | | | > 0.5mg/L 260/560(46.4%) | | > 0.55mg/L: 21(14.09%) | > 1.5 mg/L: 3(3.75%) | |
| **Albumin, g/L** | NR | NR | NR | 39.00(36.20-44.20) | 41.65(5.48) | 38.3(37.0-46.2) | NR |
| | | | | | > 57 g/L: 3(2.01%) | | |
| | | | | | < 35 g/L: 9(6.04%) | < 40 g/L: 2(2.50%) | |
| **Alanine aminotransferase, U/L** | 22(14-34) | NR | NR | 26.98(16.88-46.09) | 20(20.5) | 24(12-38) | NR |
| | | | > 40 U/L: 158/741(21.3%) | | > 64 U/L: 18(12.08%) | > 50 U/L: 3(3.75%) | |
| **Aspartate aminotransferase, U/L** | 26(20-32) | NR | NR | 24.75(18.71-31.79) | 23(15.75) | 30(19-39) | NR |
| | ⩾40 U/L: 10(16.1%) | | > 40 U/L: 168/757(22.2%) | | > 40 U/L: 27(18.12%) | > 40 U/L: 3(3.75%) | |
| **Total bilirubin, mmol/L** | NR | NR | NR | 16.40(11.34-21.15) | 9.9(5.95) | 6.6(5.4-12.0) | NR |
| | | | > 17.1 mmol/L: 76/722(10.5%) | | > 24 mmol/L: 4(2.68%) | > 12 mmol/L: 1(1.25%) | |
| **Potassium, mmol/L** | 3.7(3.5-3.9) | NR | 3.8(3.5– 4.2) | NRN | NR | NR | NR |
| **Sodium, mmol/L** | 139(127-141) | NR | 138.2(136.1–140.3) | NR | NR | NR | NR |
| **Serum creatinine, µmol/L** | 72.0(61.0-84.0) | NR | NR | NR | 66.07(14.79) | 78(60-90) | NR |
| | > 133 µmol/L: 3(5%) | | > 133 µmol/L: 12/752(1.6%) | | > 73 µmol/L: 43(28.86%) | > 111 µmol/L: 2(2.50%) | |
| | | | | | < 41 µmol/L: 1(0.67%) | | |
| **Creatine kinase–MB, U/L** | NR | NR | NR | NR | NR | 22(15-25) | NR |
| | | | | | | 16(20.00%) | |

| | | | | | | | |
|---|---|---|---|---|---|---|---|
| **Creatine kinase, U/L** | 69.0(40.5-101.0) | NR | NR | NR | 76.5(53.5) | 99(61-191) | NR |
| | > 185 U/L: 5(8%) | | > 200 U/L: 90/657(13.7%) | | > 200 U/L: 12(8.05%)<br>< 40 U/L:19(12.75%) | > 310 U/L: 18(22.50%) | |
| **Lactate dehydrogenase, U/L** | 205.0(184.0-260.5) | NR | NR | NR | 210(94.5) | 226(182-308) | NR |
| | > 245 U/L: 17(27%) | | ≥ 250 U/L: 277/675(41.0%) | | > 250 U/L: 45(30.20%) | > 250 U/L: 17(21.25%) | |
| **Procalcitonin, ng/mL** | 0.04(0.03-0.06) | 0.187(0.0633) | NR | NR | NR | 1.3(0.4-2.6) | NR |
| | ⩾0.1 ng/mL: 7(11%) | | ≥0.5 ng/mL: 35/633(5.5%) | | | ⩾5.0 ng/mL: 1(1.25%) | |
| **Blood urea nitrogen, mmol/L** | NR | NR | NR | NR | 3.8(1.4) | 4.9(3.4-5.9) | NR |
| | | | | | < 2.6 mmol/L: 17(11.41%) | > 9.5 mmol/L: 2(2.50%) | |
| **C-reactive protein, mg/L** | NR | 14.7(10.2) | NR | NR | 7.25(23) | 6.6(5.3-12.3) | NR |
| | NR | NR | ≥10 mg/L: 481/793(60.7%) | > 10 mg/L: 13(40.63%) | > 6 mg/L: 82(55.03%) | > 5 mg/L: 62(77.50%) | Increased: 26(52%) |
| **Others** | | | | Neutrophil percentage, %: 63.80(58.10-72.55); Lymphocyte percentage, %: 25.20(19.33-33.05) | Glucose, mmol/L: 5.83(1.97), > 6.1 mmol/L: 59(39.60%), < 3.9 mmol/L: 1(0.67%) | Glucose, mmol/L: 6.8(4.6-7.7), > 6.1 mmol/L: 19(23.17%); Erythrocyte sedimentation rate, mm/h: 11.9(9.0-17.2), > 15 mm/h: 59(73.75%) | |

Continuous data are median(IQR) or mean(SD); Categorical data are n(N%). NR: not reported.

### Table S31 Laboratory findings of the confirmed patients outside of former epicenter/ nationwide b

| | Huang 2020 C | Chang 2020 | Liu 2020 D | Qian 2020 | Chen 2020 C | Peng 2020 | Xu 2020 D |
|---|---|---|---|---|---|---|---|
| **White blood cell count, × $10^9$/L** | NR | NR | (2.3-8.1), range | 4.99(4.25-5.69) | 4.71(3.80-5.86) | 6.41(1.57), mean±SD | A* |
| **White blood cell count > 10 × $10^9$/L** | NR | NR | 0 | > 9.5 × $10^9$/L: 3(32.97%) | NR | NR | NR |
| **White blood cell count < 4× $10^9$/L** | Decreased: 11(31.43%) | NR | 17(40.5%) | < 3.5× $10^9$/L: 14(15.39%) | NR | NR | 17(33.3%) |
| **Neutrophil count, × $10^9$/L** | NR | NR | NR | 2.91(2.3-3.58) | NR | NR | NR |
| | | | > 5.8× $10^9$/L: 1(2.4%) | > 6.3×$10^9$/L: 3(3.30%) | | | |
| | | | < 2× $10^9$/L: 12(28.6%) | < 1.8×$10^9$/L: 10(10.99%) | | | |
| **Monocyte count, $10^9$/L** | NR | NR | NR | 0.4(0.3-0.58), | NR | NR | NR |
| | | | | > 0.6×$10^9$/L: 18(19.78%) | | | |
| **Eosinophils count, $10^9$/L** | NR | NR | NR | 0.01(0.01-0.05); | NR | 0.02(0.03) | |
| | | | | < 0.02×$10^9$/L: 47(51.65) | | | |
| **Basophils count, $10^9$/L** | NR | NR | NR | 0.01(0.01-0.02), | NR | 0.0007(0.03) | |
| | | | | > 0.06×$10^9$/L: 1(1.10) | | | |
| **Lymphocyte count, × $10^9$/L** | NR | NR | NR | 1.35(0.98-1.66) | 1.12(0.79-1.49) | 1.43(0.60), mean±SD | B* |
| | Decreased: 22(62.86%) | | < 1.1× | < 1.1×$10^9$/L: 28(30.77%) | | | |

| | | | | | | | |
|---|---|---|---|---|---|---|---|
| | | | $10^9$/L: 22(52.4%) | | | | |
| **Hemoglobin, g/L** | NR | 147(12.1) | NR | 135(125-145) | NR | NR | NR |
| | | | | < 115g/L: 33(36.26%) | | | |
| **Platelet count, × $10^9$/L** | NR | 199(72.5) | NR | 196(142-238) | NR | NR | NR |
| | Decreased: 3(8.57%) | | | < 125× $10^9$/L: 10(10.99%) | | | |
| | | | | > 350× $10^9$/L: 3(3.30%) | | | |
| **D-dimer, mg/L** | NR | NR | NR | 0.22(0.28) | NR | NR | NR |
| | | | | > 243ng/L: 22(24.18%) | | | |
| **Albumin, g/L** | NR | NR | NR | 40(37.85-42) | 40.8(37.9-43.0) | NR | NR |
| | | | | < 40 g/L: 43(47.25) | | | |
| **Fibrinogen, g/L** | NR | NR | NR | 3.4(2.7-4.04) | NR | NR | NR |
| | Increased 12(34.29%) | | | > 4g/L: 22(24.18%) | | | |
| | | | | < 2g/L: 3(3.30) | | | |
| **Alanine aminotransferase, U/L** | NR | NR | NR | 18(13-28) | 23(15-33) | NR | C* |
| | | | Increased: 7(16.7%) | > 50 U/L: 7(7.69%) | | | |
| **Aspartate aminotransferase, U/L** | NR | NR | NR | 21(17-28) | 25(20-33) | NR | D* |
| | | | Increased: 9(21.4%) | > 50 U/L: 9(9.89) | | | |
| **Total bilirubin, mmol/L** | NR | NR | NR | NR | NR | NR | NR |
| | | | Increased: 5(11.9%) | | | | |
| **Potassium, mmol/L** | NR | NR | NR | 4.04(3.72-4.23) | NR | NR | NR |
| | Decreased: 9(25.71%) | | Decreased: | < 3.5mmol/L: 7(7.69%) | | | |

| | | | | | | | |
|---|---|---|---|---|---|---|---|
| | | | 6(14.3%) | | | | |
| **Sodium, mmol/L** | NR | NR | NR | 139(137.25-141) | NR | NR | NR |
| | | | | < 137mmol/L: 16(17.58) | | | |
| **Serum creatinine, µmol/L** | NR | NR | NR | 66(57.75-77) | NR | NR | NR |
| | | | | > 73 µmol/L: 3(3.30%)<br>< 57 µmol/L: 21(23.08%) | | | |
| **Creatine kinase, U/L** | NR | NR | NR | NR | NR | NR | NR |
| | | | Increased: 3(7%) | | | | |
| **Lactate dehydrogenase, U/L** | NR | NR | NR | NR | 229(195-291) | NR | NR |
| | | | Increased: 14(33.3%) | | | | |
| **Procalcitonin, ng/mL** | NR | 0.187(0.0633) | NR | 0.02(0-0.04) | NR | NR | NR |
| | | | All in normal range | > 0.04ng/mL: 14(15.39%) | | | |
| **C-reactive protein, mg/L** | NR | 14.7(10.2) | NR | 6.81(1.87-15.30) | 12(4.4-29.4) | 7.66(7.33), mean±SD | E* |
| | Increased: 22(62.86%) | NR | > 8 mg/L: 22(52.4%) | > 5 mg/L: 49(53.85%) | | | |
| **Others** | Chloride: Decreased 15(42.86%); CD3+: Decreased 28(80%); CD4+: Decreased 28(80%); CD8+: Decreased 19(54.29%); CD4+/CD8+: Decreased 7(20%); Arterial partial pressure of | | | Red blood cell, $\times 10^{12}$/L: 4.49(4.11-4.81), < 3.8$\times 10^{12}$/L: 10(10.99%); Thrombocytocrit, %: 0.2(0.15-0.24), > 0.36%: 4(4.40%), < 0.19%: 39(42.86%); Urea, mmol/L: 3.95(3.3-4.49), < 3.1mmol/L: 19(20.88%); Ca2+, mmol/L: 2.16(2.06-2.25), > 2.52mmol/L: 1(1.10%), < 2.11mmol/L: 11(12.09); Cl-, mmol/L: 103(100.55-105), > 110mmol/L: 1(1.10%), < 99mmol/L: | Erythrocyte sedimentation rate, mm/h: 54(33-90); Lactate, mmol/L: 1.4(1.1-2.1); Estimated glomerular filtration rate, mL/min/1.73$m^2$: 109.2(95.3-127.2); CD4 T cells counts, cells/Ul: 431(299-637); | | F*, G*, H*, I* |

| | | |
|---|---|---|
| oxygen: Decreased 9(25.71%). | 8(8.79%); PaO2, mmHg: 75.6(53.5-90.38), > 108mmHg: 6(17.65%), < 83mmHg: 18(52.94%); PaCO2, mmHg: 36.5(28-41), > 48mmHg: 1(2.94%), < 35nnHg: 2(5.88%); Lactic acid, mmol/L: 1.4(1.1-1.93), > 1.6mmol/L: 19(55.88%); PH: 7.44(7.40-7.46), > 7.45: 10(29.41%). | CD4/CD8: 1.68(1.24-1.33). |

Continuous data are median(IQR) or mean(SD); Categorical data are n(N%). NR: not reported. A*: 5.2(4.6-6.3) for imported cases(n=15), 4.8(3.4-6.2) for secondary cases(n=17) and 3.9(3.1-5.7) for tertiary cases(n=19). B*: 1.5(0.9-2.1) for imported cases(n=15), 1.1(1.0-1.9) for secondary cases(n=17) and 1.3(0.9-1.6) for tertiary cases(n=19). C*: 22.9(16.6-31.0) for imported cases(n=15), 13.5(11.6-21.1) for secondary cases(n=17) and 25.2(11.7-36.2) for tertiary cases(n=19). D*: 19.0(17.0-31.0) for imported cases(n=15), 17.0(12.5-19.5) for secondary cases(n=17) and 22.0(17.0-34.0) for tertiary cases(n=19). E*: 4.4(1.3-11.2) for imported cases(n=15), 2.2(1.0-16.2) for secondary cases(n=17) and 7.0(0.8-20.0) for tertiary cases(n=19). F*: CD3+ T lymphocyte, %: 69.9(62.9-78.2) for imported cases(n=15), 65.9(60.0-72.9) for secondary cases(n=17) and 72.2(65.8-76.1) for tertiary cases(n=19). G*: CD4+ T lymphocyte, %: 39.9(38.2-45.7) for imported cases(n=15), 37.5(32.2-43.9) for secondary cases(n=17) and 37.4(32.6-40.4) for tertiary cases(n=19). H*: CD8+ T lymphocyte, %: 28.7(22.7-33.9) for imported cases(n=15),26.7(20.8-31.9) for secondary cases(n=17) and 30.3(26.0-34.0) for tertiary cases(n=19). I*: B lymphocyte, %: 13.9(9.8-18.1) for imported cases(n=15), 11.4(8.4-16.1) for secondary cases(n=17) and 9.1(7.5-10.9) for tertiary cases(n=19).

#### Table S32 Laboratory findings of the confirmed patients outside of former epicenter c

| | Wan 2020 | Li 2020 B | Wu 2020 B | Zheng 2020 A | Lo 2020 | Feng 2020 | Diao 2020 | Gao 2020 | Li 2020 C |
|---|---|---|---|---|---|---|---|---|---|
| **White blood cell count, × 10⁹/L** | 5.4(4.1-7.8) | 5.30(4.2-6.80) | 5.40(4.20–6.95) | 4.36(3.33, 5.42) | 5.0(4.4-5.4) | 5.29(4.22-7.02) | 5.49(4.97-6.48) | NR | NR |
| **White blood cell count ＞ 10 × 10⁹/L** | > 9.5 × 10⁹/L: 9(6.7%) | Increased: 9(10.8%) | Increased: 10(10%) | 3(1.9%) | NR | 49/475(10.3%) | NR | | NR |
| **White blood cell count ＜ 4× 10⁹/L** | < 3.5× 10⁹/L: 28(20.7%) | Decreased: 10(12%) | Decreased: 7(9%) | 66(41.0%) | Leukopenia: 2(20%) | 91/475(19.2%) | Decreased: 1(6.7%) | 9(22.5%) | 15(30.6%) |
| **Neutrophil count, × 10⁹/L** | 3.5(2.6-4.4) | 3.61 A* | 3.74(2.67–5.20) | NR | 2.6(2.3-3.6) | 3.56(2.61-5.42) | 3.48(3.23-4.92) | NR | NR |
| | | Increased: 17(20.5%) A* | Increased: 16(20%) | | | | | | Neutrophil percentage > 75%: 27(55.1%) |
| | | Decreased: 5(6%) A* | Decreased: 5(6%) | | Neutropenia: 1(10%) | | Decreased: 1(6.7%) | | |
| **Monocyte count, 10⁹/L** | NR | 0.39(0.27-0.53) B* | 0.41(0.27–0.53) | NR | 0.5(0.5-0.6) | NR | NR | NR | NR |
| | | Increased: 12(14.5%) B* | Increased: 8(10%) | | | | | | |
| **Lymphocyte count, × 10⁹/L** | 1.1(0.7-1.5) | 1.05(0.75-1.32) | 1.15(0.76–1.40) | 1.07(0.77-1.43) | 1.3(1.0-1.7) | 1.03(0.7-1.45) | 1.24(1.00-1.76) | NR | NR |
| | < 1.1× 10⁹/L: 68(50.4%) | Decreased: 44(53%) | Decreased: 34(43%) | < 0.8×10⁹/L: 42(26.1%) | Lymphopenia: 7(70%) | < 1× 10⁹/L: 225(47.3%) | Decreased: 5(33.3%) | < 1× 10⁹/L: 18(45%) | < 1× 10⁹/L: 30(61.2%) |
| **Hemoglobin, g/L** | 133(122-147) | NR | NR | 130(120-141) | 13.1(12.5-14.0) | 132(121-144) | NR | NR | NR |
| | | | | < 110g/L: 13(8.1%) | | | | | |

| | | | | | | | | | |
|---|---|---|---|---|---|---|---|---|---|
| **Platelet count, × $10^9$/L** | 158(131-230) | NR | NR | 168(131-220) | 178(163.5-206.8) | 184(145-238) | NR | NR | NR |
| | < 125× $10^9$/L: 23(17%) | | | < 100× $10^9$/L: 11(6.8%) | | | | | < 100× $10^9$/L: 5(10.2%) |
| **Prothrombin time, s** | 10.9(10.5-11.4) | NR | NR | NR | 12.2(11.7-12.8) | NR | NR | NR | 11.9(2), M(QR) |
| **Activated partial thromboplastin time, s** | 26.9(24.7-29) | NR | NR | NR | 34.5(32.4-37.2) | NR | NR | NR | 28.5(4.7), M(QR) |
| **D-dimer, mg/L** | 0.4(0.2-0.6) | NR | NR | NR | | 0.58(0.35-1.48) | NR | NR | 0.66(0.99), M(QR) |
| **Albumin, g/L** | 40.5(37-43.4) | NR | NR | NR | NR | 37.87(32.8-41.84) | NR | NR | 35.9(10.8), M(QR) |
| **Alanine aminotransferase, U/L** | 26(12.9-33.15) | NR | NR | 20.3(15.0, 24.5) | 24(19.3-35.3) | 26(16-41) | NR | NR | NR |
| | | | | > 40 U/L: 13(8.1%) | | | | | ⩾40 U/L: 13(26.5%) |
| **Aspartate aminotransferase, U/L** | 33.4(27.8-43.7) | NR | NR | 25.1(19.9, 32.8) | 20(9.0-42.5) | 28(21-39) | NR | NR | NR |
| | > 40 U/L: 30(22.2%) | | | > 40 U/L: 22(13.7%) | | | | | ⩾35U/L: 11(22.4%) |
| **Total bilirubin, mmol/L** | 8.6(5.9-13.7) | NR | NR | 10.9(8.37-15.43) | 6(4.0-7.0) | 10.1(7.5-14) | NR | NR | NR |
| | | | | > 20.5 mmol/L: 9(5.6%) | | | | | |
| **Potassium, mmol/L** | 4(3.55-4.41) | NR | NR | NR | NR | 3.9(3.6-4.2) | NR | NR | NR |
| **Sodium, mmol/L** | 139(137- | NR | NR | NR | NR | 139(137-141) | NR | NR | NR |

| | | | | | | | | | |
|---|---|---|---|---|---|---|---|---|---|
| | 141) | | | | | | | | |
| **Serum creatinine, μmol/L** | 66(57.8-74.5) | NR | NR | 48.2(38.9, 58.2) | 56(46.3-70.8) | 66.77(53.66-78.6) | NR | NR | NR |
| | | | | > 87 μmol/L: 2(1.2%) | | | | | ⩾84μmol/L: 8(16.3%) |
| **Creatine kinase–MB, U/L** | NR | NR | NR | NR | NR | 13(10.49-16.74) | NR | NR | NRN |
| **Creatine kinase, U/L** | 82.2(56.3-146.3) | NR | NR | NR | 87(60.8-138.5) | 82(55-148) | NR | NR | NR |
| | > 200 U/L: 10(7.4%) | | | | Increased: 1(10%) | | | | |
| **Lactate dehydrogenase, U/L** | 320.5(248.5-385.3) | NR | NR | 177.1(141.0-221.9) | 228(206.5-240.5) | 259(202-356) | NR | NR | NR |
| | > 250 U/L: 58(43%) | | | > 225 U/L: 38(23.6%) | Increased: 6(60%) | | | | ⩾214U/L: 43(87.8%) |
| **Procalcitonin, ng/mL** | 0.11(0.08-0.16) | 0.048(0.032-0.085) | 0.04(0.03–0.07) | NR | 0.03(0.03-0.04) | 0.05(0.02-0.08) | NR | NR | 0.02(0.02), M(QR) |
| | ⩾0.5: 1(0.7%) <0.1: 110(81.5%) | Increased: 44(53%) | Increased: 32(40%) | | | | | | |
| **Blood urea nitrogen, mmol/L** | NR | NR | NR | NR | NR | NR | NR | NR | 4.7(1.6), M(QR) |
| **C-reactive protein, mg/L** | 10.5(2.7-51.2) | 16.7(3.58-60.87) | 12.39(2.71–50.61) | 17.9(8.1-36.7) | 0.4(0.1-0.7), mg/dL | 18.8(5.23-57) | NR | NR | NR |
| | | Increased: 50(60.2%) | Increased: 37(46%) | > 8 mg/L: 121(75.2%) | Increased: 4(40%) | ⩾10mg/L: 266/415(64.1%) | | ⩾10mg/L: 22(55%) | |
| **Others** | | Oxyhemoglobin saturation, %: | $SaO_2$, % room air: 97%(96%–98%); | Oxygenation index: | Urea, mmol/L: 3.8(3.0-4.3); | Direct bilirubin, μmol/L: 4(3.1-5.5); Urea, mmol/L: 4.8(3.67-5.89); | Eosinophils percentage, %: Decreased, 9(60.0%); | Serum amyloid A | IL-6, ng/L: 47.9(67.3), M(QR). |

| | | | | | | |
|---|---|---|---|---|---|---|
| 97(95.1-98); Decreased: 16(19.3%) | PH value: 7.45(0.02); PaCO2, mm Hg: 39.21(7.25); PaO2, mm Hg: 85.73(22.11) | 366(316-393); | | eGFR, ml/min/1.73m$^2$: 106(87-125); Calcium, mmol/L: 2.04(1.96-2.15); Myohemoglobin, ng/ml: 18.85(4.8-51.48); Troponin increased,no. /total: 86/384(22.4%); ESR, mm/h: 48(30-80); BNP, pg/ml: 40.85(21.64-79.37); Lactic acid, mmol/L: 2.75(2.23-3.27); Fibrinogen, g/L: 4.4(3.65-5.41); | Eosinophils count,$\times 10^9$/L: 0.01(0.00 ~ 0.04); Neutrophil-to-lymphocyte ratio: 2.81(1.69 ~ 4.53); Lymphocyte percentage, %: Decreased, 5(33.3%); Lymphocyte percentage, %: 23.40(16.80 ~ 32.60). | ≥10mg/L: 29(72.5%) |

Continuous data are median(IQR) or mean(SD); Categorical data are n(N%). NR: not reported. A*: Neutrophil ratio, %: 71.52(11.31), increased: 25(30.1%), decreased: 1(1.2%); B*: Monocyte ratio, %: 7.17(2.93), increased: 15(18.1%);

### Table S33 Laboratory findings of the confirmed patients outside of former epicenter d

| | Liang 2020 B | Mao 2020 B | Wang 2020 D | Xiang 2020 | Xu 2020 E | Yu 2020 A | Yu 2020 B |
|---|---|---|---|---|---|---|---|
| **White blood cell count, × 10⁹/L** | NR | NR | NR | 5.52±3.50 | NR | 5. 17±1. 55,(3.32-8.68, range) | NR |
| **White blood cell count ＞ 10 × 10⁹/L** | Increased: 1(3.6%) | Increased: 2(8.33%) | NR | NR | NR | NR | 5(5.7%) |
| **White blood cell count ＜ 4× 10⁹/L** | Decreased: 7(25%) | Decreased: 4(16.67%) | Decreased: 3(17.6%) | NR | Decreased: 20(62.5%) | Decreased: 3(12%) | 16(18.4%) |
| **Neutrophil count, × 10⁹/L** | NR | NR | NR | 3.84±2.94 | NR | NR | NR |
| | | | Decreased: 2(11.8%) | | | | |
| **Monocyte count, 10⁹/L** | NR | NR | NR | 0.36±0.17 | NR | NR | NR |
| **Lymphocyte count, × 10⁹/L** | NR | NR | NR | 1.16±0.71 | NR | 1.39±0.75,(range, 0.75-3.76) | NR |
| | Decreased: 17(60.7%) | Decreased: 14(58.33%) | Decreased: 7(41.2%) | | Decreased: 21(65.6%) | Decreased: 8(32%) | < 1× 10⁹/L: 30(34.5%) |
| **Hemoglobin, g/L** | NR | NR | NR | 143.3±18.2 | NR | NR | NR |
| **Platelet count, × 10⁹/L** | NR | NR | NR | 163.9±65.7 | NR | NR | NR |
| | | | | | | | < 125× 10⁹/L: 15(17.2%) |
| **Prothrombin time, s** | NR | NR | NR | 12.76±1.49 | NR | NR | NR |
| **Activated partial thromboplastin time, s** | NR | NR | NR | 32.68±13.41 | NR | NR | NR |
| **ESR, mm/h** | NR | NR | NR | 47.00±26.50 | NR | 13. 89±10. 61,(range, | NR |

| | | | | | | | |
|---|---|---|---|---|---|---|---|
| | | | | | | 2-85) | |
| | | | Increased, 7(41.2%) | | | Increased: 6(24%) | |
| **D-dimer, mg/L** | NR | NR | NR | 2.51±12.99 | | NR | NR |
| | | | Increased: 1(5.9%) | | | | |
| **Albumin, g/L** | NR | NR | NR | 42.9±5.08 | NR | NR | NR |
| **Alanine aminotransferase, U/L** | NR | NR | NR | 22.7±12.4 | NR | NR | NR |
| **Aspartate aminotransferase, U/L** | NR | NR | NR | 27.4±12.3 | NR | NR | NR |
| | | | Increased: 2(11.8%) | | | | |
| **Total bilirubin, mmol/L** | NR | NR | NR | 8.23±3.76 | NR | NR | NR |
| **Potassium, mmol/L** | NR | NR | NR | 3.83±0.55 | NR | NR | NR |
| **Sodium, mmol/L** | NR | NR | NR | 136.3±3.5 | NR | NR | NR |
| **Serum creatinine, μmol/L** | NR | NR | Decreased: 2(11.8%) | 68.8±23.6 | NR | NR | NR |
| **Creatine kinase–MB, U/L** | NR | NR | NR | NR | | NR | NR |
| | | | Increased: 3(17.6%) | | | | |
| **Creatine kinase, U/L** | NR | NR | NR | 138.2±137.7 | NR | NR | NR |
| | | | Increased: 2(11.8%) | | | | |
| | | | Decreased: 2(11.8%) | | | | |
| **Lactate** | NR | NR | NR | 276.7±123.1 | NR | NR | NR |

| | | | | | | | |
|---|---|---|---|---|---|---|---|
| **dehydrogenase, U/L** | | | | | | | |
| | | | Increased: 3(17.6%) | | | | |
| **Procalcitonin, ng/mL** | NR | NR | 0.04-0.157 | 0.06±0.08 | NR | NR | NR |
| | | Increased: 1(4.17%) | | | | | |
| **Blood urea nitrogen, mmol/L** | NR | NR | NR | NR | NR | NR | NR |
| | | | Decreased: 1(5.9%) | | | | |
| **Urea, mmol/L** | NR | NR | NR | 4.44±1.59 | 3.8(3.0-4.3) | NR | NR |
| **C-reactive protein, mg/L** | NR | NR | NR | 27.77±38.67 | | NR | NR |
| | | Increased: 9(37.5%) | Increased: 7(41.2%) | | Increased: 23(71.9%) | | > 10mg/L: 48(55.2%) |
| **Others** | | Hypoproteinemia: 13(54.17%) | | Red blood cell,$\times 10^{12}$/L: 4.56±0.53; Hematocrit, L/L: 0.41±0.05; Eosinophils (Absolute value), $\times 10^{9}$/L: 0.15±0.89; Globulin, g/L: 27.74±4.56; Direct bilirubin, μmol/L: 2.84±1.40; Alkaline phosphatase, U/L: 65.2±22.9; Uric acid, umol/L: 251.4±75.9; Triglycerides, mmol/L: 1.14±0.96; Total cholesterol, mmol/L: 3.69±0.74; High-density lipoprotein, mmol/L: 1.05±0.31; Low density lipoprotein, mmol/L: 2.35±0.72; Amylase, U/L: 65.2±33.6; $Cl^{-}$, mmol/L: 97.5±15.3; $Ca2^{+}$, mmol/L: 2.0±0.1; Serum amyloid protein A, mg/L: 113.05±90.82; Fibrinogen, g/L: 3.84±2.05. | | IL-6, pg/mL: 18. 66± 14. 86,(range, 4. 52-55. 5); IL-6 increased: 20(80. 0%). | |

Continuous data are median(IQR) or mean(SD); Categorical data are n(N%). NR: not reported.

#### Table S34 Treatment of the confirmed patients in former epicenter

| | Huang 2020 A | Chen 2020 A | Liu 2020 A | Wang 2020 A | Huang 2020 B | Cao2020 | Bin 2020 |
|---|---|---|---|---|---|---|---|
| **Oxygen therapy** | 27(66%) | 75(76%) | 85(62.0%) | 106(76.81%) | 25(73.5%) | 76(74.5%) | NR |
| **Invasive mechanical ventilation** | 2(5%) | 4(4%) | 0 | 17(12.32%) | 3(8.8%) | 14(13.7%) | NR |
| **Non-invasive mechanical ventilation** | 10(24%) | 13(13%) | 34(24.8%) | 15(10.9%) | 2(5.9%) A* | 5(4.9%) | NR |
| **CRRT** | 3(7%) | 9(9%) | NR | 2(1.45%) | NR | 6(5.9%) | NR |
| **ECMO** | 2(5%) | 3(3%) | 0 | 4(2.9%) | NR | 3(2.9%) | NR |
| **Antibiotic treatment** | 41(100%) | 70(71%) | 119(86.9%) | B* | 31(91.2%) | 101(99%) | NR |
| **Antiviral treatment** | 38(93%) | 75(76%) | 105(76.6%) | 124(89.9) | 32(94.1%) C* | 100(98%) | 55(100%) |
| **Corticosteroid therapy** | 9(22%) | 19(19%) | 40(29.2%) | 62(44.9) | 21(61.8%) | 51(50%) | 17(30.9%) |
| **Intravenous immunoglobulin therapy** | NR | 27(27%) | 44(32.1%) | NR | NR | 11(10.8%) | NR |
| **Antifungal treatment** | NR | 15(15%) | NR | NR | NR | NR | NR |
| **ICU** | 13(32%) | 23(23%) | NR | 36(26.1%) | 8(23.5%) | 18(17.6%) | NR |
| **Others** | NR | NR | NR | Vasopressors: 13(9.4%) | NR | Chinese medicine treatment: 3(2.9%). | Chinese patent medicine: 46(83.6%) D* |

Categorical data are n(N%). NR: not reported. CRPT: continuous renal replacement therapy. ECMO: extracorporeal membrane oxygenation. ICU: intensive unit care; A*: Non-invasive ventilation or high flow nasal cannula; B*: Wnag2020 reported that many patients received antibacterial therapy(moxifloxacin, 89 [64.4%]; ceftriaxone, 34 [24.6%]; azithromycin, 25 [18.1%]); C*: Other drugs but not lopinavir/ritonavir, and antiviral therapy switched to lopinavir/ritonavir later: 9(26.5%). D*: Oral Lianhua Qingwen capsule;

#### Table S35 Treatment of the confirmed patients outside of former epicenter a

| | Qian 2020 | Chen 2020 C | Dai 2020 | Xu 2020 D | Wan 2020 | Lo 2020 | Feng 2020 | Gao 2020 | Li 2020 C |
|---|---|---|---|---|---|---|---|---|---|
| **Oxygen therapy** | NR | NR | NR | NR | 90(66.7%) | 4(40%) | B* | 1(2.5%) | M* |
| **Invasive mechanical ventilation** | NR | | NR | 0 | 1(0.7%) | 0 | 39(8.2%) | NR | |
| **Non-invasive mechanical ventilation** | NR | | 11(4.7%) | 0 | 34(25.2%) A* | 0 | 34(7.1%) | NR | |
| **ECMO** | NR | | NR | NR | 0 | NR | 4(0.8%) | NR | NR |
| **Antibiotic treatment** | NR | | 118(50.4%) | NR | 59(43.7%) | 10(100%) | 319(67%) | 25(62.5%) | 32(65.3%) |
| **Antiviral treatment** | 91(100%) | | 234(100%) | NR | 135(100%) | 10(100%) | 286(60.1%) | 40(100%) | 30 (62.1%) D* |
| **Corticosteroid therapy** | 9(9.9%) | | 34(14.5%) | NR | 36(26.7%) | 3(30%) | 127(26.7%) | 6(15%) | 29(59.2%) |
| **Intravenous immunoglobulin therapy** | NR | | 34(14.5%) | NR | NR | NR | NR | NR | 24(49%) |
| **Antifungal treatment** | NR | | NR | NR | NR | 0 | 8(1.7%) | NR | NR |
| **Hemofiltration therapy** | NR | NR | NR | NR | R | NR | NR | 1(2.5%) | 2(4.1%) |
| **Interferon therapy** | NR | NR | NR | NR | NR | 3(30%) | NR | Aerosol inhalation of α interferon: 40(100%) | Aerosol inhalation: 37(75.5%) |
| **Traditional Chinese medicine** | NR | NR | NR | NR | 124(91.8%) | NR | NR | NRN | Chinese patent medicine:42(85.7%) |
| **ICU** | 9(9.89%) | 22(8.8%) | 0 | NR | NR | NR | NR | NR | NR |
| **Others** | | | | | Continuous renal replacement therapy: 5(3.7%); | | | | |

Categorical data are n(N%). NR: not reported. ECMO: extracorporeal membrane oxygenation. ICU: intensive unit care; A*: Non-invasive ventilation or high-flow nasal cannula. B*: Nasal cannula or no oxygen therapy 368(77.3%), and high-flow nasal cannula 31(6.5%). C*: Air suction 6(12.2%), intranasal oxygen inhalation 17(34.7%), oxygen therapy / assisted ventilation 26(53.1%). D*: One antiviral drug alone 30(62.1%), combined two antiviral drugs 19(38.8%).

#### Table S36 Treatment of the confirmed patients outside of former epicenter/ nationwide b

| | Wang 2020 D | Yu 2020 A | Xu 2020 A | Guan 2020 | Yang 2020 | Wu 2020 A |
|---|---|---|---|---|---|---|
| **Oxygen therapy** | NR | NR | NR | 454(41.3%) | 134(89.93%) | 35(43.75%) |
| **Invasive mechanical ventilation** | NR | NR | NR | 25(2.3%) | 0 | 0 |
| **Non-invasive mechanical ventilation** | NR | NR | NR | 56(5.1%) | 2(1.34%) | 0 |
| **ECMO** | NR | NR | NR | 5(0.5%) | NR | 0 |
| **Antibiotic treatment** | NR | NR | 28(45%) | 637(58.0%) | 34(22.82%) | 73(91.25%) |
| **Antiviral treatment** | NR | 25(100%) | 55(89%) | 393(35.8%) | 140(93.96%) | 80(100.00%) |
| **Corticosteroid therapy** | NR | 2(8%) | 16(26%) | 204(18.6%) | 5(3.36%) | NR |
| **Intravenous immunoglobulin therapy** | NR | NR | NR | 144(13.1%) | 19(12.75%) | 16(20.00%) |
| **Antifungal treatment** | NR | NR | NR | 31(2.8%) | 0 | NR |
| **Interferon therapy** | NR | Recombinant human interferon α 2B injection: 25(100%) | NR | NR | 144(96.64%) | NR |
| **Traditional Chinese medicine** | 17(100%) | NR | NR | NR | NR | 3(3.75%) |
| **ICU** | NR | NR | 1(2%) | 55(5%) | 0 | NR |
| **Others** | | | | CRRT: 9(0.8%) | | Hormone therapy: 12(14.63%). |

Categorical data are n(N%). NR: not reported. CRPT: continuous renal replacement therapy. ECMO: extracorporeal membrane oxygenation. ICU: intensive unit care.

**Table S37 Percentages of epidemiological and clinical characteristics among the confirmed patients: Non-severe/severe (median, IQR)**

| Subject | Non-severe, % | Severe, % | *p* |
|---|---|---|---|
| **Epidemiological characteristics and comorbidities** | | | |
| Male | 51(40-59) | 57(52-69) | 0.04 |
| Age>45-60, y | 48(31-55) | 78(58-78) | 0.017 |
| Current smoker | 8.1(3.8-10) | 3.4(1.3-15) | 1 |
| Former smoker | 3.7(1.3-17) | 6.9(5.2-25) | 0.4 |
| Exposure former epicenter | 62(49-83) | 59(18-85) | 0.721 |
| Exposure to confirmed patients | 44(19-68) | 41(13-70) | 1 |
| Medical staff | 3.7(0-62) | 0(0-22) | 0.7 |
| Cluster/ gathering infection | 37(16-54) | 32(8.2-58) | 0.886 |
| Death | 0(0-0.9) | 6.5(1.3-17) | 0.056 |
| Any comorbidities | 21(17-35) | 50(41-64) | 0.003 |
| Hypertension | 9.4(5.2-21) | 32(11-40) | 0.007 |
| Cardiovascular disease | 1.8(1.2-4.5) | 6.7(4-14) | 0.026 |
| Cerebrovascular disease | 2.3(1.2-2.3) | 3.3(2.3-7.3) | 0.2 |
| Cardio-cerebrovascular disease | 6.3(1-19) | 11(8-15) | 0.7 |
| COPD | 1.7(0-3.1) | 10(3.5-16) | 0.002 |
| Other chronic respiratory disease | 4.8(0-9.5) | 11(2.5-20) | 1 |
| Diabetes | 3.8(0-11) | 16(11-23) | 0.001 |
| Chronic liver disease | 4.1(1.5-13) | 1.3(0-5.8) | 0.343 |
| Immunodeficiency/HIV | 0.4(0.2-0.6) | 2(0-4) | 1 |
| Hepatitis B infection | 1.2(0-2.4) | 5.9(0.6-11) | 1 |
| Malignancy | 1.2(0.85-5.1) | 5.7(2.7-7.1) | 0.2 |
| Chronic kidney disease | 0.55(0.13-2.6) | 2(1.6-3.1) | 0.2 |
| **Signs, symptoms and complications** | | | |
| Fever | 84(72-91) | 87(69-96) | 0.724 |

| | | | |
|---|---|---|---|
| Highest temperature > 39℃ | 4.9(3.4-6.2) | 3.5(2.4-4.5) | 0.343 |
| Chills | 11(6.6-12) | 15(8.6-25) | 0.222 |
| Cough | 67(61-71) | 70(57-87) | 0.381 |
| Sputum production | 23(8.8-35) | 44(26-57) | 0.063 |
| Dyspnea | 7.7(2.4-15) | 33(23-57) | 0.001 |
| Chest tightness | 9.3(8.6-10) | 7.7(0-15.4) | 0.222 |
| Chest pain | 2.8(1.7-3.9) | 11.8(7.6-16) | 0.333 |
| Hemoptysis | 0.8(0.6-1.3) | 2.6(0.58-6.4) | 0.343 |
| Fatigue | 36(20-55) | 33(30-79) | 0.436 |
| Myalgia | 15(9.4-22) | 17(11-34) | 0.436 |
| Headache | 10(6.3-16) | 13(8.4-19) | 0.546 |
| Sore throat | 17(7.9-18) | 8.7(0-13) | 0.053 |
| Diarrhea | 11(5.2-20) | 11(1.7-24) | 0.863 |
| Nausea or vomiting | 4.6(3.9-35) | 5.2(0-10) | 0.818 |
| Abdominal pain | 2.8(1.65-7.25) | 7.4(1.7-9.88) | 0.686 |
| Dizziness | 11.1(7.1-15.1) | 9.65(0-19.3) | 1 |
| Inappetence | 11(3.55-26.8) | 15(10.85-36.95) | 0.421 |
| ARDS | 1.1(1.1-1.1) | 32.8(15.6-50) | 0.333 |
| Acute cardiac injury | 8.4(8.4-8.4) | 5(5-5) | 1 |
| Acute kidney injury | 2.05(0.1-4) | 2.7(2.5-2.9) | 1 |
| Shock | 0.05(0-0.1) | 4.45(2.5-6.4) | 0.333 |
| Secondary infection | 1.95(0-3.9) | 19.9(17.5-22.3) | 0.333 |
| **Radiological and laboratory findings** | | | |
| Bilateral involvement | 79.3(46.8-90.15) | 93(89.5-100) | 0.011 |
| Local patchy opacity | 39.2(11.5-71.8) | 55.1(7-63.3) | 1 |
| Ground glass opacity | 6.55(0.8-12.3) | 99.1(66.7-100) | 0.259 |
| Consolidation | 31.4(15.6-49.65) | 45.2(27.23-73.9) | 0.31 |
| Pleural effusion | 2.35(0-7.9) | 15.4(12.88-29.93) | 0.002 |
| Interlobular septal thickening | 59.25(43.5-75) | 69.2(96.2-69.2) | 1 |
| Crazy-paving pattern | 29.1(27.6-30.6) | 55.1(54.2-56) | 0.333 |

| | | | |
|---|---|---|---|
| Bronchial Wall Thickening | 5.2(4.1-21.3) | 64(42.8-70.8) | 0.1 |
| Air bronchogram | 40.5(27.4-53.6) | 50(46.2-53.8) | 1 |
| Lymph node enlargement | 1.8(0-6.98) | 15.2(1.78-26.83) | 0.343 |
| Upper lobes involvement | 64.7(18.58-83.6) | 92.3(23.08-95.08) | 0.343 |
| Lower lobes involvement | 75(45.98-97.5) | 96.15(60.58-100) | 0.486 |
| No abnormality | 7.65(2.5-15.73) | 0(0-2.18) | 0.057 |
| White blood cell count > 9.5-10 × $10^9$/L | 6.6(4.8-8.6) | 16(7.5-23.2) | 0.209 |
| White blood cell count < 3.5-4× $10^9$/L | 25(19.1-31.3) | 16.1(11.1-30) | 0.383 |
| Lymphocyte count < 1-1.5×$10^9$/L | 38(16-79.3) | 82.1(71.8-88.9) | 0.073 |
| Platelet count < 100-150 g/L | 11.6(6.1-31.6) | 30(10-57.7) | 0.7 |
| D-dimer > 0.5 mg/L | 35.9(28.6-43.2) | 59.45(59.3-59.6) | 0.333 |
| Alanine aminotransferase > 40 U/L | 12.95(6.1-19.8) | 22.4(16.7-28.1) | 1 |
| Aspartate aminotransferase >40 U/L | 16(7.2-18.2) | 39.4(37.5-40) | 0.1 |
| LDH > 225-250 U/L | 33.1(20.45-46.8) | 62.4(52.03-72.93) | 0.029 |
| Total bilirubin > 21-26 mmol/L | 7.25(4.6-9.9) | 11.65(10-13.3) | 0.333 |
| Creatine kinase >190-200 U/L | 4.55(2.85-10.9) | 18.25(13.38-27.25) | 0.057 |
| Serum creatinine > 87-97 μmol/L | 1.9(0.8-3) | 5.4(3.3-7.5) | 0.333 |
| Procalcitonin > 0.5 ng/mL | 3.7(0-39.7) | 13.7(2.5-84) | 0.7 |
| C-reactive protein > 10 mg/L | 56.3(41.3-62.95) | 81.5(46.9-96) | 0.151 |
| **Treatment** | | | |
| Oxygen therapy | 48.35(8.93-90.25) | 48.05(15.93-77.78) | 1 |
| Invasive mechanical ventilation | 0(0-0) | 14.5(2.5-31.5) | 0.267 |
| Non-invasive mechanical ventilation | 0(0-7.4) | 32.4(27.4-67.5) | 0.267 |
| CRRT | 0.5(0-1) | 7.6(5.2-10) | 0.333 |
| ECMO | 0(0-0) | 2.9(0-3.2) | 0.2 |
| Antibiotic treatment | 53.8(25-59.4) | 87.5(80.3-88.7) | 0.1 |
| Antiviral treatment | 68.25(39.48-95) | 79.55(52.2-97.23) | 0.686 |
| Corticosteroid therapy | 13.7(13.4-15.8) | 52.5(44.5-64.5) | 0.1 |
| Intravenous immunoglobulin therapy | 9.3(9.3-9.3) | 33.5(33.5-33.5) | 1 |
| Antifungal treatment | 1.25(0.6-1.9) | 6.15(4.8-7.5) | 0.333 |

| | | | |
|---|---|---|---|
| TCM | 83.55(75.6-91.5) | 90.7(88.9-92.5) | 1 |
| Percentage of patients in severe condition or not of each subject is in median and interquartile ranges (IQR). COPD: Chronic obstructive pulmonary disease; ARDS: Acute respiratory distress syndrome; LDH: Lactate dehydrogenase; CRRT: Continuous renal replacement therapy; ECMO: Extracorporeal membrane oxygenation; TCM: Traditional Chinese medicine. | | | |

#### Table S38 Epidemiological and clinical characteristics of the confirmed patients with severe condition or not

| | Non severe | Severe |
|---|---|---|
| **Epidemiological characteristics** | | |
| **Total** | | |
| **Guan 2020** | 926/1099(84.3%) | 173/1099(15.7%) |
| **Mao 2020 A** | 126(58.9%) | 88(41.1%) |
| **Zhang 2020 B** | 82(58.6%) | 58(41.4%) |
| **Xu 2020 B** | 37(74%) | 13(26%) |
| **Tian 2020** | 216(42.4%) | 46(17.6%) |
| **Li 2020 A** | 16(64%) | 9(36%) |
| **Qian 2020** | 82(90.1%) | 9(9.9%) |
| **Liu 2020 C** | 49(67.1%) | 24(32.9%) |
| **Wan 2020** | 95(70.4%) | 40(29.6%) |
| **Li 2020 B** | 58(69.9%) | 25(30.1%) |
| **Tang 2020** | 70(84.3%) | 13(15.7%) |
| **Zheng 2020** | 131(81.4%) | 30(18.6%) |
| **Zhao 2020** | 87(86.1%) | 14(13.9%) |
| **Lo 2020** | 6(60%) | 4(40%) |
| **Feng 2020** | 352(73.9%) | 124(26.1%) |
| **Bin 2020** | 45(81.8%) | 9(16.4%) |

| | | |
|---|---|---|
| **Wang 2020 C** | 42(43.8%) | 54(56.2%) |
| **Li 2020 C** | 22(44.9%) | 27(55.1%) |
| **Lv 2020** | 13(76.5%) | 4(23.5%) |
| **Xiang 2020** | 40(81.6%) | 9(18.4%) |
| | **Male** | |
| **Guan 2020** | 537/923(58.2%) | 100(57.8%) |
| **Mao 2020 A** | 43(34.1%) | 44(50.0%) |
| **Zhang 2020 B** | 38(46.3%) | 33(56.9%) |
| **Xu 2020 B** | 22(59.5%) | 7(53.8%) |
| **Tian 2020** | 101(46.8%) | 26(56.5%) |
| **Li 2020 A** | 6(37.5%) | 6(66.7%) |
| **Liu 2020 C** | 31(63.3%) | 10(41.7%) |
| **Wan 2020** | 52(54.7%) | 21(52.5%) |
| **Li 2020 B** | 29(50%) | 15(60%) |
| **Zheng 2020** | 66(50.4%) | 14(46.7%) |
| **Zhao 2020** | 48(55.2%) | 8(57.1%) |
| **Lo 2020** | 2(33%) | 1(25%) |
| **Feng 2020** | 190(54%) | 81(65.3%) |
| **Bin 2020** | 23(51.1%) | 7(77.8%) |
| **Wang 2020 C** | 15(35.7%) | 31(57.4%) |
| **Li 2020 C** | 9(40.9%) | 19(70.4%) |
| **Lv 2020** | 8(61.5%) | 4(100%) |
| **Xiang 2020** | 25(62.5%) | 8(88.9%) |
| | **Female** | |
| **Guan 2020** | 386/923(41.8%) | 73(42.2%) |
| **Mao 2020 A** | 83(65.9%) | 44(50.0%) |
| **Zhang 2020 B** | 44(53.7%) | 25(43.1%) |
| **Xu 2020 B** | 15(40.5%) | 6(46.2%) |
| **Tian 2020** | 115(53.2%) | 20(43.5%) |
| **Li 2020 A** | 10(62.5%) | 3(33.3%) |
| **Liu 2020 C** | 18(36.7%) | 14(58.3%) |

| | | |
|---|---|---|
| **Wan 2020** | 43(45.3%) | 19(47.5) |
| **Li 2020 B** | 29(50%) | 10(40%) |
| **Zheng 2020** | 65(49.6%) | 16(53.3%) |
| **Zhao 2020** | 39(44.8%) | 39(44.8%) |
| **Lo 2020** | 4(67%) | 3(75%) |
| **Feng 2020** | 162(46%) | 43(34.7%) |
| **Bin 2020** | 22(48.9%) | 2(22.2%) |
| **Li 2020 C** | 13(59.1%) | 8(29.6%) |
| **Lv 2020** | 5(38.5%) | 0 |
| **Xiang 2020** | 15(37.5%) | 1(11.1%) |
| | **Age, Median(IQR)** | |
| **Guan 2020** | 45.0(34.0–57.0) | 52.0(40.0–65.0) |
| **Zhang 2020 B** | 51.5, range: 26-78 | 64, range:25-87 |
| **Xu 2020 B** | 40.7, range: 3-62 | 57.2, range: 34-85 |
| **Tian 2020** | 44.5, range: 1-93 | 61.4, range: 1-94 |
| **Wan 2020** | 44(33-49) | 56(52-73) |
| **Zheng 2020** | 40(31-51) | 57(46.5-66) |
| **Feng 2020** | 51(37-63) | Severe: 58(48-67), n=54; Critical: 61(49-68), n=70 |
| **Li 2020 C, M(QR)** | 31.5(17.5) | 54(23.5) |
| | **Age, ≥50(y)** | |
| **Guan 2020** | 350/848(54.7%) | 95/163(58.3%) |
| **Mao 2020 A** | 60(47.6%) | 64(72.7%) |
| **Zhang 2020 B** | 50(61%) | 48(82.8%) |
| **>50, Xu 2020 B** | 10(27%) | 5(38.5%) |
| **≥45, Tian 2020** | 103(47.7%) | 36(78.2%) |
| **＞51, Li 2020 A** | 5(31.3%) | 7(77.8%) |
| **≥65, Feng 2020** | 73(20.7%) | 45(36.3%) |
| **＞60, Wang 2020 C** | 17(40.5%) | 42(77.8%) |
| | **Age, Mean(SD)** | |
| **Mao 2020 A** | 48.9(14.7) | 58.2(15.0) |
| **Qian 2020** | 49(35.3-56) | 66(54-80) |

| | | |
|---|---|---|
| **Liu 2020 C** | Mild: 29.2(10.9); Common: 33.4(12.2) | Severe: 44.2(12.0); Critical: 63.0(21.2) |
| **Li 2020 B** | 41.9(10.6) | 53.7(12.3) |
| **Tang 2020** | 43.7(15) | 57.4(11.8) |
| **Zhao 2020** | 42.94(11.7) | 53.71(12.6) |
| **Lo 2020** | 37(19) | 61(5) |
| **Bin 2020** | 52.1(17.6) | 64.4(9.9) |
| **Lv 2020** | 53.4(12.5) | 47.8(8.3) |
| **Xiang 2020** | 40.6(14.3) | 53(14) |
| | **Current smoker** | |
| **Guan 2020** | 108/913(11.8%) | 29/172(16.9%) |
| **Zhang 2020 B** | 0 | 2(3.4%) |
| **Wan 2020** | 8(8.4%) | 1(2.5%) |
| **Feng 2020** | 27/333(8.1) | 17/121(14%) |
| **Xiang 2020** | 3(7.5%) | 0 |
| | **Former smoker** | |
| **Guan 2020** | 12/913(1.3%) | 9/172(5.2%) |
| **Zhang 2020 B** | 3(3.7%) | 4(6.9%) |
| **Lo 2020** | 1(17%) | 1(25%) |
| | **Never smoking** | |
| **Guan 2020** | 793/913(86.9%) | 134/172(77.9%) |
| **Li 2020 A** | 14(87.5%) | 4(44.4%) |
| | **BMI** | |
| **＞24kg/m$^2$, Wang 2020 C** | 14(33.3%) | 30(55.6%) |
| **Xiang 2020** | 24.55±4.04 | 26.17±3.93 |
| | **Exposure to the former epicenter** | |
| **Xu 2020 B** | 24(48%) | 6(12%) |
| **Tian 2020** | 48(22.2%) | 5(10.9%) |
| **Liu 2020 C** | 33(67.3%) | 21(87.5%) |
| **Wan 2020** | 56(58.9%) | 15(37.5%) |
| **Zheng 2020** | 84(64.1%) | 20(66.7%) |
| **Feng 2020** | 312(88.6) | 113(91.1%) |

| | | |
|---|---|---|
| **Li 2020 C** | 11(50%) | 14(51.9%) |
| **Xiang 2020** | 39(97.5%) | 7(77.7%) |
| **Exposure to confirmed patients** | | |
| **Wan 2020** | 18(18.9%) | 5(12.5%) |
| **Li 2020 C** | 15(68.2%) | 19(70.4%) |
| **Contact with wildlife** | | |
| **Guan 2020** | 10/559(1.8%) | 3/128(2.3%) |
| **Onset of symptoms to hospital admission(d)** | | |
| **Zhang 2020 B** | 8(5-11) | 7(6-12) |
| **Tian 2020** | 4.4(3.5) | 5.2(4.6) |
| **Li 2020 B** | 6(3-8.5) | 8(6-12) |
| **Zheng 2020** | 6(3-8) | 6(3.75-8) |
| **Feng 2020** | 6(3-10) | Severe: 7(4-10), n=54; Critical: 9(7-13), n=70 |
| **Incubation period** | | |
| **Guan 2020** | 4.0(2.8–7.0) | 4.0(2.0–7.0) |
| **Tian 2020** | 2.1(1.9) | 2.1(1.6) |
| **Li 2020 C, M(QR)** | 7(3) | 6(2.5) |
| **Xiang 2020** | 4.7(4.4) | 4.5(0.7) |
| **Medical staff** | | |
| **Zhang 2020 B** | 3(3.7%) | 0 |
| **Li 2020 A** | 10(62%%) | 2(22.2%) |
| **Lo 2020** | 0 | 0 |
| **Familiar/cluster/ Close contact infections** | | |
| **Zhang 2020 B** | 11(13.4) | 4(6.9) |
| **Xu 2020 B** | 12(24%) | 6(12%) |
| **Tian 2020** | 109(50.5%) | 24(52.2%) |
| **Li 2020 C** | 12(54.5%) | 16(59.3%) |
| **Clinical outcomes** | | |
| **Discharge** | | |
| **Guan 2020** | 50(5.4%) | 5(2.9%) |
| **Tian 2020** | 43(19.9%) | 2(4.3%) |

| | | |
|---|---|---|
| **Wan 2020** | 10(1.05%) | 5/(12.5%) |
| **Feng 2020** | 334(94.9%) | 69(55.6%) |
| | **Staying in hospital** | |
| **Guan 2020** | 875(94.5%) | 154(89.0%) |
| **Tian 2020** | 173(80.1%) | 41(89.1%) |
| **Wan 2020** | 85(89.5%) | 35(87.5%) |
| **Feng 2020** | 6(1.7%) | 17(13.7%) |
| | **Death** | |
| **Guan 2020** | 1(0.1%) | 14(8.1%) |
| **Tian 2020** | 0 | 3(6.5%) |
| **Wan 2020** | 0 | 1/(2.5%) |
| **Lo 2020** | 0 | 0 |
| **Feng 2020** | 6(1.7%) | 32(25.8%) |
| | **Comorbidity** | |
| | **Any comorbidity** | |
| **Guan 2020** | 194(21.0%) | 67(38.7%) |
| **Mao 2020 A** | 41(32.5%) | 42(47.7%) |
| **Zhang 2020 B** | 44(53.7%) | 46(79.3%) |
| **Wan 2020** | 15(16.3%) | 28(70%) |
| **Li 2020 B** | 4(6.9%) | 11(44%) |
| **Tang 2020** | 12(17.1%) | 4(30.8%) |
| **Zhao 2020** | 23(26.4%) | 7(50%) |
| **Lo 2020** | 1(17%) | 2(50%) |
| **Feng 2020** | 133(37.8%) | 72(58.1%) |
| | **Hypertension** | |
| **Guan 2020** | 124(13.4%) | 41(23.7%) |
| **Mao 2020 A** | 19(15.1%) | 32(36.4%) |
| **Zhang 2020 B** | 20(24.4%) | 22(37.9%) |
| **Li 2020 A** | 1(6%) | 1(11.1%) |
| **Wan 2020** | 9(9.4%) | 4(10%) |
| **Li 2020 B** | 3(5.2%) | 2(8%) |

| | | |
|---|---|---|
| **Zheng 2020** | 10(7.6%) | 12(40%) |
| **Feng 2020** | 73(20.7%) | 40(32.3%) |
| **Wang 2020 C** | 15(35.7%) | 35(64.8%) |
| **Li 2020 C** | 1(4.5%) | 8(29.6%) |
| **Xiang 2020** | 2(5%) | 4(44.4%) |
| **Diabetes** | | |
| **Guan 2020** | 53(5.7%) | 28(16.2%) |
| **Mao 2020 A** | 15(11.9%) | 15(17.0%) |
| **Zhang 2020 B** | 9(11.0%) | 8(13.8%) |
| **Li 2020 A** | 0 | 1(11.1%) |
| **Wan 2020** | 3(3.1%) | 9(22.5%) |
| **Li 2020 B** | 0 | 7(28%) |
| **Zheng 2020** | 5(3.8%) | 2(6.7%) |
| **Feng 2020** | 32(9.1%) | 17(13.7%) |
| **Wang 2020 C** | 6(14.3%) | 19(35.2%) |
| **Li 2020 C** | 0 | 1(3.7%) |
| **Xiang 2020** | 0 | 2(22.2%) |
| **Cardiovascular disease** | | |
| **Feng 2020** | 21(6%) | 17(13.7%) |
| **Li 2020 C** | 1(4.5%) | 4(14.8%) |
| **Guan 2020（Coronary heart disease)** | 17(1.8%) | 10(5.8%) |
| **Zhang 2020 B (Coronary heart disease)** | 3(3.7%) | 4(6.9%) |
| **Li 2020 B (Coronary heart disease)** | 0 | 1(4%) |
| **Zheng 2020 (Coronary heart disease)** | 2(1.5%) | 2(6.7%) |
| **Zhang 2020 B (Aorta sclerosis)** | 1(1.2%) | 1(1.7%) |
| **Cerebrovascular disease** | | |
| **Guan 2020** | 11(1.2%) | 4(2.3%) |
| **Zheng 2020** | 3(2.3%) | 1(3.3%) |
| **Feng 2020** | 8(2.3%) | 9(7.3%) |
| **Cardio cerebrovascular disease** | | |
| **Mao 2020 A** | 8(6.3) | 7(8.0) |

| | | |
|---|---|---|
| **Li 2020 A** | 3(18.8%) | 1(11.1%) |
| **Wan 2020** | 1(1%) | 6(15%) |
| **Chronic gastritis and gastric ulcer** | | |
| **Zhang 2020 B** | 5(6.1%) | 2(3.4%) |
| **Cholelithiasis** | | |
| **Zhang 2020 B** | 2(2.4%) | 4(6.9%) |
| **Hyperlipidemia** | | |
| **Zhang 2020 B** | 5(6.1%) | 2(3.4%) |
| **COPD** | | |
| **Guan 2020** | 6(0.6%) | 6(3.5%) |
| **Zhang 2020 B** | 0 | 2(3.4%) |
| **Li 2020 A** | 1(6%) | 4(44.4%) |
| **Wan 2020** | 0 | 4(10%) |
| **Li 2020 B** | 1(1.7%) | 4(16%) |
| **Zheng 2020** | 4(3.1%) | 2(6.7%) |
| **Feng 2020** | 8(2.3%) | 14(11.3%) |
| **Malignancy** | | |
| **Guan 2020** | 7(0.8%) | 3(1.7%) |
| **Mao 2020 A** | 8(6.3%) | 5(5.7%) |
| **Wan 2020** | 1(1%) | 3(7.5%) |
| **Feng 2020** | 5(1.4%) | 7(5.6%) |
| **Chronic kidney disease** | | |
| **Guan 2020** | 5(0.5%) | 3(1.7%) |
| **Mao 2020 A** | 4(3.2%) | 2(2.3%) |
| **Zhang 2020 B** | 0 | 2(3.4%) |
| **Feng 2020** | 2(0.6%) | 2(1.6%) |
| **Chronic liver disease** | | |
| **Wan 2020** | 1(1%) | 1(2.5%) |
| **Zheng 2020** | 4(3.1%) | 0 |
| **Xiang 2020** | 6(15%) | 0 |
| **Zhang 2020 B (Fatty liver and abnormal liver function)** | 4(5.0%) | 4(6.9%) |

| | | |
|---|---|---|
| | **Immunodeficiency/HIV** | |
| **Guan 2020** | 2(0.2%) | 0 |
| **Feng 2020(Immunosuppression)** | 2(0.6%) | 5(4%) |
| | **Hepatitis B infection** | |
| **Guan 2020** | 22(2.4%) | 1(0.6%) |
| **Li 2020 C** | 0 | 3(11.1%) |
| | **Arrhythmia** | |
| **Zhang 2020 B** | 1(1.2%) | 4(6.9%) |
| | **Thyroid diseases** | |
| **Zhang 2020 B** | 1(1.2%) | 4(6.9%) |
| | **Electrolyte imbalance** | |
| **Zhang 2020 B** | 0 | 4(6.9%) |
| | **Urolithiasis** | |
| **Zhang 2020 B** | 2(2.4%) | 1(1.7%) |
| | **Stroke** | |
| **Zhang 2020 B** | 1(1.2%) | 2(3.4%) |
| | **Aorta sclerosis** | |
| **Zhang 2020 B** | 1(1.2%) | 1(1.7%) |
| | **Secondary pulmonary tuberculosis** | |
| **Zhang 2020 B** | 0 | 2(3.4%) |
| | **Other pulmonary disease** | |
| **Wan 2020** | 0 | 1(2.5%) |
| **Wang 2020 C** | 4(9.5%) | 11(20.4%) |
| | **Signs and symptoms on admission** | |
| | **Fever** | |
| **Guan 2020** | 391/910(43%) | 82/171(48%) |
| **Mao 2020 A** | 92(73.0%) | 40(45.5%) |
| **Zhang 2020 B** | 59/67(88.1%) | 51/53(96.2%) |
| **Xu 2020 B** | 33(66%) | 10(20%) |
| **Tian 2020** | 178(82.4%) | 37(80.4%) |
| **Li 2020 A** | 15(93.8%) | 9(100%) |

| | | |
|---|---|---|
| **Liu 2020 C** | 45(91.8%) | 23(95.8%) |
| **Wan 2020** | 86(90.1%) | 34(85%) |
| **Li 2020 B** | 50(86.2%) | 22(88%) |
| **Tang 2020** | 61(87.1%) | 11(84.6%) |
| **Zheng 2020** | 93(71%) | 29(96.7%) |
| **Feng 2020** | 277/337(82.2%) | 113/117(91.1%) |
| **Wang 2020 C** | 30(71.4%) | 41(75.9%) |
| **Li 2020 C** | 22(100%) | 25(92.6%) |
| **Lv 2020** | 13(100%) | 4(100%) |
| **Xiang 2020** | 32(80%) | 6(66.7%) |
| **Fever during hospitalization** | | |
| **Guan 2020** | 816/926(88.1%) | 159/173(91.9%) |
| **Highest temperature＞39℃** | | |
| **Guan 2020** | 30/910(3.3%) | 8/171(4.7%) |
| **Xu 2020 B** | 3(6%) | 2(4%) |
| **Tian 2020** | 8(3.7%) | 1(2.2%) |
| **Wan 2020** | 6(6.3%) | 1(2.9%) |
| **Highest temperature＞39℃during hospitalization** | | |
| **Guan 2020** | 88/774(11.4%) | 26/152(17.1%) |
| **Highest temperature＜37.5℃** | | |
| **Guan 2020** | 519/910(57.0%) | 89/171(52.0%) |
| **<37.3℃, Xu 2020 B** | 7(19%) | 3(23%) |
| **<37.3°C, Tian 2020** | 38(17.6%) | 9(19.6%) |
| **<37.3°C, Li 2020 A** | 1(6.2%) | 0 |
| **<37.3°C, Wan 2020** | 11(11.6%) | 5(14.3%) |
| **Highest temperature＜37.5℃ during hospitalization** | | |
| **Guan 2020** | 79/774(10.2%) | 13/152(8.6%) |
| **Cough** | | |
| **Guan 2020** | 623(67.3%) | 122(70.5%) |
| **Mao 2020 A(dry cough)** | 77(61.1%) | 30(34.1%) |
| **Zhang 2020 B** | 45/67(67.2%) | 45/53(84.9%) |

| | | |
|---|---|---|
| **Xu 2020 B** | 14(28%) | 6(12%) |
| **Tian 2020** | 95(44.0%) | 25(54.3%) |
| **Li 2020 A** | 11(68.8%) | 6(66.7%) |
| **Liu 2020 C** | 40(81.6%) | 20(83.3%) |
| **Wan 2020** | 67(70.5%) | 35(87.5%) |
| **Li 2020 B** | 41(70.7%) | 24(96%) |
| **Tang 2020** | 50(71.4%) | 10(76.9%) |
| **Zheng 2020** | 80(61.1%) | 21(70%) |
| **Feng 2020(Dry cough)** | 220/336(65.5%) | 49/117(39.5%) |
| **Wang 2020 C** | 26(61.9%) | 44(81.5%) |
| **Li 2020 C** | 19(86.4%) | 18(66.7%) |
| **Lv 2020** | 11(84.6%) | 4(100%) |
| **Xiang 2020** | 13(32.5%) | 6(66.7%) |
| | **Sputum production** | |
| **Guan 2020** | 309(33.4%) | 61(35.3%) |
| **Xu 2020 B** | 5(10%) | 2(4%) |
| **Liu 2020 C** | 23(46.9%) | 16(66.7%) |
| **Wan 2020** | 5(5.3%) | 7(17.5%) |
| **Li 2020 B** | 6(10.3%) | 9(36%) |
| **Tang 2020** | 26(37.1%) | 8(61.5%) |
| **Feng 2020** | 100/336(29.8%) | 61/117(52.1%) |
| **Li 2020 C** | 5(22.7%) | 14(51.9%) |
| **Xiang 2020** | 3(7.5%) | 4(44.4%) |
| | **Dyspnea** | |
| **Guan 2020** | 140(15.1%) | 65(37.6%) |
| **Zhang 2020 B** | 20/67(29.9%) | 24/53(45.3%) |
| **Xu 2020 B(chest tightness or dyspnea)** | 0 | 4(8%) |
| **Tian 2020** | 3(1.4%) | 15(32.6%) |
| **Li 2020 A** | 12(75%) | 8(88.9%) |
| **Wan 2020** | 0 | 18(18.9%) |
| **Li 2020 B** | 2(3.4%) | 7(28%) |

| | | |
|---|---|---|
| **Tang 2020** | 3(4.3%) | 4(30.8%) |
| **Tang 2020(气促)** | 11(15.7%) | 8(61.5%) |
| **Zheng 2020** | 14(10.7%) | 9(30%) |
| **Feng 2020** | 50/335(14.9%) | 59/112(52.7%) |
| **Li 2020 C** | 1(4.5%) | 3(11.1%) |
| **Lv 2020** | 1(7.7%) | 3(75%) |
| | **Fatigue** | |
| **Guan 2020** | 350(37.8%) | 69(39.9%) |
| **Xu 2020 B** | 2(4%) | 6(12%) |
| **Tian 2020** | 54(25.0%) | 15(32.6%) |
| **Li 2020 A(fatigue or muscular soreness)** | 13(81.3%) | 4(44.4%) |
| **Liu 2020 C** | 35(71.4%) | 20(83.3%) |
| **Tang 2020** | 21(30%) | 4(30.8%) |
| **Zheng 2020** | 49(37.4%) | 15(30%) |
| **Li 2020 C** | 8(36.4%) | 24(88.9%) |
| **Lv 2020** | 10(76.9%) | 3(75%) |
| **Xiang 2020** | 6(15%) | 3(33.3%) |
| | **Myalgia** | |
| **Guan 2020(myalgia or arthralgia)** | 134(14.5%) | 30(17.3%) |
| **Mao 2020 A** | 6(4.8%) | 17(19.3%) |
| **Zhang 2020 B** | 51/67(76.1%) | 39/53(73.6%) |
| **Xu 2020 B** | 4(8%) | 4(8%) |
| **Wan 2020(myalgia or arthralgia)** | 25(26.3%) | 19(47.5%) |
| **Li 2020 B** | 10(17.2%) | 5(20%) |
| **Tang 2020** | 11(15.7%) | 1(7.7%) |
| **Zheng 2020** | 14(10.7%) | 4(13.3%) |
| **Feng 2020** | 38/333(11.4%) | 17/105(16.2%) |
| | **Headache** | |
| **Guan 2020** | 124(13.4%) | 26(15.0%) |
| **Mao 2020 A** | 15(17.0%) | 13(10.3%) |
| **Xu 2020 B** | 3(6%) | 2(4%) |

| | | |
|---|---|---|
| **Tian 2020** | 14(6.5%) | 3(6.5%) |
| **Wan 2020** | 23(24.2%) | 11(27.5%) |
| **Li 2020 B** | 6(10.3%) | 3(12%) |
| **Tang 2020** | 10(14.3%) | 2(15.4%) |
| **Zheng 2020** | 8(6.1%) | 4(13.3%) |
| **Xiang 2020(headache/dizziness)** | 4(10%) | 2(22.2%) |
| | **Hemoptysis** | |
| **Guan 2020** | 6(0.6%) | 4(2.3%) |
| **Wan 2020** | 1(1%) | 3(7.5%) |
| **Tang 2020** | 1(1.4%) | 0 |
| **Feng 2020** | 2/332(0.6%) | 3/103(2.9%) |
| | **Diarrhea** | |
| **Guan 2020** | 32(3.5%) | 10(5.8%) |
| **Mao 2020 A** | 28(22.2%) | 13(14.8%) |
| **Zhang 2020 B** | 9/82(11.0%) | 9/57(15.8%) |
| **Li 2020 A** | 4(25%) | 1(11.1%) |
| **Wan 2020** | 5(5.3%) | 13(32.5%) |
| **Tang 2020** | 5(7.1%) | 0 |
| **Zheng 2020** | 16(12.2%) | 1(3.3%) |
| **Li 2020 C** | 4(18.2%) | 27(100%) |
| **Xiang 2020** | 2(5%) | 0 |
| | **Gastrointestinal reaction** | |
| **Xu 2020 B** | 1(2%) | 0 |
| **Feng 2020(Digestive symptoms)** | 39/336(11.6%) | 10/110(9.1%) |
| | **Sore throat** | |
| **Guan 2020** | 130(14.0%) | 23(13.3%) |
| **Mao 2020 A** | 21(16.7%) | 10(11.4) |
| **Xu 2020 B** | 3(6%) | 1(2%) |
| **Wan 2020** | 24(25.3%) | 0 |
| **Tang 2020** | 12(17.1%) | 2(15.4%) |
| **Feng 2020** | 26/330(7.9%) | 9/103(8.7%) |

| | | |
|---|---|---|
| **Xiang 2020** | 7(17.5%) | 0 |
| | **Respiratory rate** | |
| **Tian 2020** | 123.7(14.4) | 126.1(16.4) |
| **Wan 2020(＞24/min)** | 3(3.2%) | 9(22.5%) |
| **Li 2020 B** | 20(20-22) | 21(19.5-24.5) |
| **Xiang 2020** | 20.0±2.4 | 23.2±6.2 |
| | **Nausea or vomiting** | |
| **Guan 2020** | 43(4.6%) | 12(6.9%) |
| **Zhang 2020 B(nausea)** | 19/82(23.2%) | 5/57(8.8%) |
| **Zhang 2020 B(vomiting)** | 5/82(6.1%) | 2/57(3.5%) |
| **Wan 2020(retching)** | 4(4.2%) | 0 |
| **Tang 2020** | 2(2.9%) | 2(15.4%) |
| **Zheng 2020** | 6(4.6%) | 0 |
| | **Chill** | |
| **Guan 2020** | 100(10.8%) | 26(15.0%) |
| **Wan 2020** | 7(7.4%) | 7(17.5%) |
| **Tang 2020** | 8(11.4%) | 1(7.7%) |
| **Feng 2020** | 17/300(5.7%) | 7/74(9.5%) |
| **Xiang 2020** | 5(12.5%) | 3(33.3%) |
| | **Dizziness** | |
| **Mao 2020 A** | 19(15.1%) | 17(19.3%) |
| **Tang 2020** | 5(7.1%) | 0 |
| | **Anorexia** | |
| **Mao 2020 A** | 47(37.3%) | 21(23.9%) |
| **Zhang 2020 B** | 9/82(11.0%) | 8/57(14.0%) |
| **Liu 2020 C** | 8(16.3%) | 12(50%) |
| **Wan 2020** | 0 | 6(15%) |
| **Tang 2020** | 5(7.1%) | 1(7.7%) |
| | **Abdominal pain** | |
| **Mao 2020 A** | 4(3.2%) | 6(6.8%) |
| **Zhang 2020 B** | 2/82(2.4%) | 6/57(10.5%) |

| | | |
|---|---|---|
| **Li 2020 B(Abdominal pain/ Diarrhea)** | 5(8.6%) | 2(8%) |
| **Tang 2020** | 1(1.4%) | 0 |
| **Tonsil swelling** | | |
| **Guan 2020** | 17(1.8%) | 6(3.5%) |
| **Nasal congestion** | | |
| **Guan 2020** | 47(5.1%) | 6(3.5%) |
| **Conjunctival congestion** | | |
| **Guan 2020** | 5(0.5) | 4(2.3) |
| **Enlargement of lymph nodes** | | |
| **Guan 2020** | 1(0.1%) | 1(0.6%) |
| **Impaired consciousness** | | |
| **Mao 2020 A** | 3(2.4%) | 13(14.8%) |
| **Acute cerebrovascular disease** | | |
| **Mao 2020 A** | 1(0.8%) | 5(5.7%) |
| **Ataxia** | | |
| **Mao 2020 A** | 0 | 1(1.1%) |
| **Seizure** | | |
| **Mao 2020 A** | 0 | 1(1.1%) |
| **Taste impairment** | | |
| **Mao 2020 A** | 9(7.1%) | 3(3.4%) |
| **Smell impairment** | | |
| **Mao 2020 A** | 8(6.3%) | 3(3.4%) |
| **Vision impairment** | | |
| **Mao 2020 A** | 1(0.8%) | 2(2.3%) |
| **Nerve pain** | | |
| **Mao 2020 A** | 1(0.8%) | 4(4.5%) |
| **Neurological symptoms** | | |
| **Feng 2020** | 35/334(10.5%) | 12/106(11.3%) |
| **Rash** | | |
| **Guan 2020** | 0 | 2(1.2%) |
| **Throat congestion** | | |

| | | |
|---|---|---|
| **Guan 2020** | 17(1.8%) | 2(1.2%) |
| **Belching** | | |
| **Zhang 2020 B** | 4/82(4.9%) | 3/57(5.3%) |
| **Chest operation history** | | |
| **Li 2020 A** | 6(37.5%) | 7(77.8%) |
| **Palpitation** | | |
| **Wan 2020** | 2(2.1%) | 3(7.5%) |
| **Chest tightness and shortness of breath** | | |
| **Wan 2020** | 9(9.5%) | 3(7.5%) |
| **Diastolic pressure, mm Hg** | | |
| **Wan 2020** | 76(71-86) | 76(70-80) |
| **Xiang 2020** | 80.7±12.7 | 69.1±10.8 |
| **Systolic pressure, mm Hg** | | |
| **Wan 2020** | 119(111-128) | 121(112-133) |
| **Xiang 2020** | 123.8±14.9 | 122.1±16.9 |
| **Median arterial pressure(mmHg)** | | |
| **Li 2020 B** | 93.3(87.2-103.3) | 91(84.3-100.5) |
| **Pharyngeal discomfort** | | |
| **Li 2020 B** | 4(6.9%) | 2(8%) |
| **Chest pain** | | |
| **Li 2020 B** | 1(1.7%) | 4(16%) |
| **Feng 2020** | 13/335(3.9%) | 8/105(7.6%) |
| **Tang 2020(Chest distress)** | 6(8.6%) | 2(15.4%) |
| **Xiang 2020(Chest distress)** | 4(10%) | 0 |
| **Others** | | |
| **Tang 2020(Catarrhal symptoms)** | 7(10%) | 0 |
| **Tang 2020(Thirst)** | 1(1.4%) | 1(7.7%) |
| **Xiang 2020(Asymptomatic infection)** | 3(7.5%) | 0 |
| **Complications** | | |
| **ARDS** | | |
| **Guan 2020** | 10(1.1%) | 27(15.6%) |

| | | |
|---|---|---|
| **Wan 2020** | 1(1.1%) | 20(50%) |
| **Acute kidney injury** | | |
| **Guan 2020** | 1(0.1%) | 5(2.9%) |
| **Wan 2020** | 4(4%) | 1(2.5%) |
| **Acute cardiac injury** | | |
| **Wan 2020** | 8(8.4%) | 2(5%) |
| **Shock** | | |
| **Guan 2020** | 1(0.1%) | 11(6.4%) |
| **Wan 2020** | 0 | 1(2.5%) |
| **Rhabdomyolysis** | | |
| **Guan 2020** | 2(0.2%) | 0 |
| **Disseminated intravascular coagulation** | | |
| **Guan 2020** | 0 | 1(0.6%) |
| **Pneumonia** | | |
| **Guan 2020** | 800/894(89.5%) | 172(99.4%) |
| **Secondary infection** | | |
| **Wan 2020** | 0 | 7(17.5%) |
| **Feng 2020** | 12/307(3.9%) | 23/103(22.3%) |
| **Main radiological findings** | | |
| **Bilateral involvement of chest CT on admission** | | |
| **Guan 2020** | 368/808(45.5%) | 137/167(82.0%) |
| **Zhang 2020 B** | 68/78(87.2%) | 53/57(93.0%) |
| **Xu 2020 B(Bilateral upper lobes)** | 15(53.6%) | 12(92.3%) |
| **Xu 2020 B(Bilateral lower lobes)** | 12(42.9%) | 13(100.0%) |
| **Liu 2020 C** | 31(63.3%) | 24(100%) |
| **Li 2020 B** | 54(93.1%) | 25(100%) |
| **Tang 2020** | 58/62(93.5%) | 12(92.3%) |
| **Zheng 2020** | 63(48.1%) | 26(86.7%) |
| **Zhao 2020** | 69(79.3) | 14(100) |
| **Feng 2020** | 266/327(81.3%) | 107/115(93%) |
| **Li 2020 C** | 7(31.8%) | 27(100%) |

| | | |
|---|---|---|
| | **Local patchy shadowing of chest CT on admission** | |
| **Guan 2020** | 317/808(39.2%) | 92/167(55.1%) |
| **Zhang 2020 B** | 9/78(11.5%) | 4/57(7%) |
| **Zheng 2020** | 94(71.8%) | 19(63.3%) |
| | **Number of lobs involved of chest CT on admission** | |
| **≤ 3: Xu 2020 B** | 11(39.3%) | 1(7.7%) |
| **4-5: Xu 2020 B** | 17(60.7%) | 12(92.3%) |
| **Li 2020 B** | 5(3-5) | 5(5-5) |
| **Feng 2020** | 5(3-5) | Severe: 5(5-5), n=54; Critical: 5(5-5), n=61 |
| **Xiang 2020** | 2.5±1.4 | 3.4±1.8 |
| | **Ground glass opacity of chest CT on admission** | |
| **Guan 2020** | 449/808(55.6%) | 101/167(60.5%) |
| **Xu 2020 B** | 21(75.0%) | 9(69.2%) |
| **Liu 2020 C(Unique)** | 12(24.5%) | 3(12.5%) |
| **Liu 2020 C(Multiple)** | 31(63.3%) | 19(79.2%) |
| **Li 2020 B** | 56(96.6%) | 25(100%) |
| **Tang 2020** | 61/62(98.4%) | 13(100%) |
| **Zheng 2020** | 62(47.3%) | 20(66.7%) |
| **Zhao 2020** | 73(83.9%) | 14(100%) |
| **Feng 2020** | 311/327(95.1%) | 114/115(99.1%) |
| | **Interstitial abnormalities of chest CT on admission** | |
| **Guan 2020** | 99/808(12.3%) | 44/167(26.3%) |
| **Zheng 2020** | 1(0.8%) | 1(3.3%) |
| | **No abnormality of chest CT on admission** | |
| **Guan 2020** | 157/877(17.9%) | 5/173(2.9%) |
| **Zhang 2020 B** | 1/78(1.3%) | 0 |
| **Liu 2020 C** | 3(6.1%) | 0 |
| **Zhao 2020** | 8(9.2%) | 0 |
| | **Paving Stone Sign** | |
| **Liu 2020 C** | 15(30.6%) | 13(54.2%) |
| **Li 2020 B** | 16(27.6%) | 14(56%) |

| | | |
|---|---|---|
| | **Consolidation** | |
| **Liu 2020 C** | 0 | 8(33.3%) |
| **Li 2020 B** | 31(53.4%) | 22(88%) |
| **Tang 2020** | 30(48.4%) | 4(30.8%) |
| **Zhao 2020** | 36(41.4%) | 8(57.1%) |
| **Feng 2020** | 68/327(20.8%) | 19/115(16.5%) |
| **Xu 2020 B** | 6(21.4) | 9(69.2) |
| | **Bronchial Wall Thickening** | |
| **Liu 2020 C** | 2(4.1%) | 17(70.8%) |
| **Li 2020 B** | 3(5.2%) | 16(64%) |
| **Zhao 2020** | 22(25.3%) | 6(42.8%) |
| | **Intrathoracic lymph node enlargement** | |
| **Xu 2020 B(Mediastinal nodes)** | 1(3.6) | 0 |
| **Zhao 2020** | 0 | 1(7.1) |
| **Tang 2020(Mediastinal nodes)** | 5(8.1%) | 3(23.3%) |
| **Li 2020 B** | 0 | 7(28%) |
| | **Pleural Effusion** | |
| **Liu 2020 C** | 0 | 3(12.5%) |
| **Li 2020 B** | 0 | 7(28%) |
| **Tang 2020** | 1/62(1.6%) | 2(15.4%) |
| **Feng 2020** | 10/327(3.1%) | 15/115(13%) |
| **Xu 2020 B** | 2(7.1%) | 2(15.4%) |
| **Zhao 2020** | 9(10.3%) | 5(35.7%) |
| | **Interlobular septa thickening** | |
| **Xu 2020 B** | 21(75.0%) | 9(69.2%) |
| **Tang 2020** | 27(43.5%) | 9(69.2%) |
| | **Air bronchogram** | |
| **Xu 2020 B** | 15(53.6) | 7(53.8) |
| **Tang 2020** | 17(27.4) | 6(46.2) |
| | **Upper lobes involvement** | |
| **Xu 2020 B** | 15(53.6) | 12(92.3) |

| | | |
|---|---|---|
| **Zhao 2020** | 6(6.9) | 0 |
| **Tang 2020** | 47(75.8) | 12(92.3) |
| **Li 2020 B** | 50(86.2%) | 24(96%) |
| | **Lower lobes involvement(CT)** | |
| **Xu 2020 B** | 12(42.9) | 13(100.0) |
| **Zhao 2020** | 48(55.2) | 7(50) |
| **Tang 2020** | 61(98.4) | 12(92.3) |
| **Li 2020 B** | 55(94.8%) | 25(100%) |
| | **Main laboratory findings** | |
| | **White blood cell count, × $10^9$/L** | |
| **Guan 2020** | 4.9(3.8–6.0) | 3.7(3.0–6.2) |
| **Mao 2020 A** | 4.5(1.8-14.0) | 5.4(0.1-20.4) |
| **Zhang 2020 B** | 4.5(3.5-5.9) | 5.3(4.0-9.0) |
| **Qian 2020** | 4.97(4.02-5.65) | 5.23(4.74-6.8) |
| **Wan 2020** | 5.5(4.0-8.0) | 5.2(4.9-6.9) |
| **Li 2020 B** | 5.27(4.2-6.3) | 5.5(4.1-7.7) |
| **Zheng 2020** | 4.17(3.33-5.24) | 4.76(3.18-5.74) |
| **Lo 2020** | 4.51(1.18) | 5.57(1.99) |
| **Feng 2020** | 5.15(4.17-6.54) | Severe: 5.42(3.69-8.17); Critical: 7.19(4.61-11.19) |
| **Bin 2020** | 4.94±1.49 | 4.62±1.77 |
| **Li 2020 C, M(QR)** | 5.21(2.54) | 5.74(5.17) |
| **Xiang 2020** | 4.61±1.88 | 9.56±6.01 |
| | **White blood cell count ＞ 10 × $10^9$/L** | |
| **Guan 2020** | 39/811(4.8%) | 19/167(11.4%) |
| **Zhang 2020 B(＞9.5 × $10^9$/L)** | 4/82(4.9%) | 13/56(23.2%) |
| **Li 2020 A(＞9.5 × $10^9$/L)** | 9(56.3%) | 4(44.5%) |
| **Wan 2020(＞9.5 × $10^9$/L)** | 6(7%) | 3(7.5%) |
| **Li 2020 B(Increased)** | 5(8.6%) | 4(16%) |
| **Zheng 2020** | 3(2.3%) | 0 |
| **Feng 2020** | 23/351(6.6%) | 26(21%) |
| | **White blood cell count ＜ 4× $10^9$/L** | |

| | | |
|---|---|---|
| **Guan 2020** | 228/811(28.1%) | 102/167(61.1%) |
| **Zhang 2020 B(＜3.5× 10⁹/L)** | 18/82(22.0%) | 9/56(16.1%) |
| **Li 2020 A** | 5(31.3%) | 1(11.1%) |
| **Wan 2020(＜3.5× 10⁹/L)** | 24(25%) | 4(10%) |
| **Li 2020 B(Decreased)** | 6(10.3%) | 4(16%) |
| **Zheng 2020** | 57(43.5%) | 9(30%) |
| **Feng 2020** | 67/351(19.1%) | 24/124(19.4%) |
| | **Neutrophil count, × 10⁹/L** | |
| **Mao 2020 A** | 2.6(0.7-11.8) | 3.8(0.0-18.7) |
| **Qian 2020** | 2.8(2.18-3.49) | 3.32(3-5.82) |
| **Wan 2020** | 3.6(3.0-3.9) | 4.1(3.1-5.7) |
| **Li 2020 B** | 3.5(2.64-4.46) | 4.36(2.87-6.48) |
| **Lo 2020** | 2.49(0.9) | 3.82(1.81) |
| **Feng 2020** | 3.39(2.5-4.64) | Severe: 3.6(2.59-5.99); Critical: 5.99(3.47-9.55) |
| **Li 2020 C(Percentage), M(QR)** | 68.7(15.1%) | 83.1(8%) |
| **Xiang 2020** | 2.96±1.41 | 7.76±4.61 |
| | **Eosinophils, × 10⁹/L** | |
| **Zhang 2020 B** | 0.02(0.008-0.05) | 0.01(0.0-0.06) |
| **Xiang 2020** | 0.02±0.07 | 0.70±2.08 |
| | **Eosinophils <0.02 × 10⁹/L** | |
| **Zhang 2020 B** | 39/82(47.6%) | 34/56(60.7%) |
| | **Eosinophils percentage, %** | |
| **Zhang 2020 B** | 0.5(0.08-1.0) | 0.2(0.0-0.8) |
| **Qian 2020** | 0.02(0.01-0.06) | 0.01(0-0.01) |
| | **Basophil** | |
| **Qian 2020** | 0.01(0.01-0.02) | 0(0-0.01) |
| | **Lymphocyte count, × 10⁹/L** | |
| **Guan 2020** | 1.0(0.8–1.4) | 0.8(0.6–1.0) |
| **Mao 2020 A** | 1.3(0.4-2.6) | 0.9(0.1-2.6) |
| **Zhang 2020 B** | 0.8(0.6-1.2) | 0.7(0.5-1.0) |
| **Qian 2020** | 1.4(1.05-1.75) | 0.9(0.7-1.3) |

| | | |
|---|---|---|
| **Wan 2020** | 1.2(0.8-1.6) | 0.8(0.6-1.0) |
| **Li 2020 B** | 1.23(0.93-1.42) | 0.7(0.44-0.95) |
| **Zheng 2020** | 1.12(0.84, 1.45) | 0.85(0.69, 1.06) |
| **Lo 2020** | 1.4(0.54) | 1.18(0.49) |
| **Feng 2020** | 1.13(0.79-1.53) | Severe: 0.78(0.52-1.08); Critical: 0.82(0.49-1.08) |
| **Bin 2020** | 1.25±0.43 | 0.83±0.32 |
| **Li 2020 C, M(QR)** | 1.17(0.31) | 0.59(0.24) |
| **Xiang 2020** | 1.26±0.69 | 0.72±0.66 |
| **Lymphocyte percentage, %** | | |
| **Zhang 2020 B** | 20.0(12.5-28.4) | 12.7(7.7-22.0) |
| **Lymphocyte count＜1.5× 10⁹/L** | | |
| **Guan 2020** | 584/736(79.3%) | 147/154(95.5%) |
| **Zhang 2020 B(＜1.1× 10⁹/L)** | 58/82(70.7%) | 46/56(82.1%) |
| **Xu 2020 B(Decreased)** | 8(16%) | 6(12%) |
| **Li 2020 A(＜1.1× 10⁹/L)** | 14(87.5%) | 8(88.9%) |
| **Wan 2020** | 36(38%) | 32(80%) |
| **Li 2020 B(Decreased)** | 22(37.9%) | 22(88%) |
| **Zheng 2020(＜0.8× 10⁹/L)** | 29(22.1%) | 13(43.3%) |
| **Feng 2020(＜1× 10⁹/L)** | 136(38.6%) | 89(71.8%) |
| **Mononuclear leucocyte count, ×10⁹/L** | | |
| **Qian 2020** | 0.4(0.3-0.59) | 0.35(0.3-0.7) |
| **Li 2020 B** | 0.42(0.29-0.53) | 0.34(0.15-0.51) |
| **Li 2020 B(Increased)** | 10(17.2%) | 2(8%) |
| **Li 2020 B(Decreased)** | 0 | 2(8%) |
| **Xiang 2020** | 0.35±0.14 | 0.37±0.25 |
| **Red blood cell** | | |
| **Qian 2020** | 4.49(4.17-4.81) | 4.26(3.8-4.82) |
| **Xiang 2020** | 4.56±0.55 | 4.55±0.41 |
| **Hemoglobin, g/L** | | |
| **Guan 2020** | 135.0(120.0–148.0) | 128.0(111.8–141.0) |
| **Qian 2020** | 135(126-147) | 130(118-142) |

| | | |
|---|---|---|
| **Wan 2020** | 134(124-147) | 130(120-143) |
| **Feng 2020** | 133(121-144) | Severe: 132(123-144); Critical: 131(118-143) |
| **Xiang 2020** | 143.0±19.3 | 144.8±12.6 |
| **Hemoglobin＜110 g/L** | | |
| **Zheng 2020** | 11(8.4%) | 2(6.7%) |
| **Oxyhemoglobin saturation, %** | | |
| **Li 2020 B** | 97(96-98) | 95.1(92.9-97.45) |
| **Li 2020 B(Decreased)** | 5(8.6%) | 11(44%) |
| **Platelet count, × $10^9$/L** | | |
| **Guan 2020** | 172.0(139.0–212.0) | 137.5(99.0–179.5) |
| **Mao 2020 A** | 219.0(42.0 - 583.0) | 204.5(18.0 - 576.0) |
| **Qian 2020** | 198(144-248) | 152(127-208) |
| **Wan 2020** | 170(136-234) | 147(118-213) |
| **Zheng 2020** | 171(137,221) | 160(133,214) |
| **Feng 2020** | 185(146-238) | Severe: 184(138-216); Critical: 181(135-246) |
| **Li 2020 C, M(QR)** | 172(81) | 153(173) |
| **Xiang 2020** | 161.3±67.7 | 175.4±57.9 |
| **Platelet count＜150× $10^9$/L** | | |
| **Guan 2020** | 225/713(31.6%) | 90/156(57.7%) |
| **Wan 2020(＜125× $10^9$/L)** | 11(11.6%) | 12(30%) |
| **Zheng 2020(＜100× $10^9$/L)** | 8(6.1%) | 3(10%) |
| **Prothrombin time, s** | | |
| **Wan 2020** | 10.8(10.4-11.3) | 11.3(10.7-11.8) |
| **Li 2020 C, QR** | 12.8(2) | 11.5(1.6) |
| **Xiang 2020** | 12.47±0.66 | 14.02±2.98 |
| **Activated partial thromboplastin time,s** | | |
| **Wan 2020** | 26.6(24.5-28.8) | 29.7(226.2-39.4) |
| **Li 2020 C, M(QR)** | 29.6(5.3) | 28.1(6.1) |
| **Xiang 2020** | 30.86±3.80 | 40.57±30.04 |
| **Thrombocytocrit** | | |
| **Qian 2020** | 0.21(0.16-0.25) | 0.16(0.13-0.21) |

| | | |
|---|---|---|
| | **Fibrinogen** | |
| **Qian 2020** | 3.36(2.64-4.08) | 3.8(3.71-4.15) |
| **Xiang 2020** | 3.71±2.16 | 4.38±1.51 |
| **Feng 2020** | 4.31(3.55-5.33) | Severe: 4.78(4.33-5.74); Critical: 4.71(3.89-5.74) |
| | **Albumin, g/L** | |
| **Liu 2020 B** | 39.70(36.50-46.10) | 35.75(28.68-42.00) |
| **Qian 2020** | 40.2(38-42.4) | 38.55(36.33-39.25) |
| **Wan 2020** | 49.9(37.4-43.6) | 36(33-38.5) |
| **Feng 2020** | 39.14(35.15-42.7) | Severe: 35.93(32.05-39.56); Critical: 32.25(27.88-34.35) |
| **Li 2020 C, M(QR)** | 41.7(3.4) | 30.8(5.4) |
| **Xiang 2020** | 44.28±4.46 | 37.23±3.30 |
| | **D-dimer, mg/L** | |
| **Mao 2020 A** | 0.4(0.2-8.7) | 0.9(0.1-20.0) |
| **Zhang 2020 B** | 0.2(0.1-0.3) | 0.4(0.2-2.4) |
| **Qian 2020** | 300(106-400) | 450(160-485) |
| **Wan 2020** | 0.3(0.2-0.5) | 0.6(0.4-1.1) |
| **Feng 2020** | 0.51(0.32-1.08) | Severe: 0.89(0.44-2.33); Critical: 1.11(0.51-4) |
| **Li 2020 C, M(QR)** | 0.25(0.09) | 0.98(3.35) |
| **Xiang 2020** | 0.53±1.34 | 11.11±29.69 |
| | **D-dimer＞0.5mg/L** | |
| **Guan 2020** | 195/451(43.2%) | 65/109(59.6%) |
| **Zhang 2020 B(＞0.243mg/L)** | 12/43(27.9%) | 23/38(60.5%) |
| **Wang 2020 C(Increased)** | 12(28.6%) | 32(59.3%) |
| | **Alanine aminotransferase, U/L** | |
| **Mao 2020 A** | 23.0(6.0-261.0) | 32.5(5.0-1933.0) |
| **Liu 2020 B** | 22.75(16.31-37.25) | 60.25(40.88-68.90) |
| **Qian 2020** | 18(13-29) | 19.9(14-26) |
| **Wan 2020** | 21.7(14.8-36.9) | 26.6(14.5-33.3) |
| **Zheng 2020A** | 19.3(14.6-17.8) | 23.9(17.6, 35.3) |
| **Feng 2020** | 23(15-38) | Severe: 32(21-47); Critical: 35(25-53) |
| **Li 2020 C, M(QR)** | 27(34) | 29(19) |

| | | |
|---|---|---|
| **Xiang 2020** | 20.7±11.8 | 32.0±11.2 |
| | **Alanine aminotransferase＞40 U/L** | |
| **Guan 2020** | 120/606(19.8%) | 38/135(28.1%) |
| **Zheng 2020** | 8(6.1%) | 5(16.7%) |
| | **Aspartate aminotransferase, U/L** | |
| **Mao 2020 A** | 23.0(9.0-244.0) | 34.0(8.0-8191.0) |
| **Liu 2020 B** | 23.63(18.71-26.50) | 37.00(20.88-64.45) |
| **Qian 2020** | 21(17-29) | 27(23.75-27) |
| **Wan 2020** | 22.4(16.9-30.5) | 33.6(25.7-44.2) |
| **Zheng 2020** | 23.4(19.0-28.8) | 31.6(25.9-49.36) |
| **Lo 2020** | 19(8) | 24(9) |
| **Feng 2020** | 25(19-34) | Severe: 34(26-53); Critical: 39(30-54) |
| **Li 2020 C, M(QR)** | 23(11) | 29(11) |
| **Xiang 2020** | 25.1±10.3 | 38.1±15.7 |
| | **Aspartate aminotransferase＞40 U/L** | |
| **Guan 2020** | 112/615(18.2%) | 56/142(39.4%) |
| **Wan 2020** | 15(16%) | 15(37.5%) |
| **Zheng 2020** | 10(7.6%) | 12(40%) |
| | **Total bilirubin, mmol/L** | |
| **Liu 2020 B** | 15.95(11.34-20.83) | 20.50(11.28-25.00) |
| **Zheng 2020** | 10.7(8.18-15.3) | 12.7(9.2-16.9) |
| **Feng 2020** | 9.5(7.3-13.3) | Severe: 11.9(8.9-15.6); Critical: 12.2(8.6-16.7) |
| **Xiang 2020** | 7.50±3.35 | 11.48±3.97 |
| **Wan 2020** | 8.6(5.6-14) | 9.8(7.8-15.6) |
| | **Total bilirubin＞17.1 mmol/L** | |
| **Guan 2020** | 59/594(9.9%) | 17/128(13.3%) |
| **Zheng 2020** | 6(4.6%) | 3(10%) |
| | **Potassium, mmol/L** | |
| **Guan 2020** | 3.9(3.6–4.2) | 3.8(3.5–4.1) |
| **Qian 2020** | 4.09(3.72-4.27) | 3.82(3.76-3.89) |
| **Wan 2020** | 4(3.7-4.5) | 3.8(3.5-4.3) |

| | | |
|---|---|---|
| **Feng 2020** | 3.9(3.6-4.1) | Severe: 4(3.5-4.2); Critical: 4(3.7-4.6) |
| **Xiang 2020** | 3.80±0.52 | 3.92±0.68 |
| **Sodium, mmol/L** | | |
| **Guan 2020** | 138.4(136.6–140.4) | 138.0(136.0–140.0) |
| **Qian 2020** | 139.6(137.6-142) | 137.85(135.3-139.38) |
| **Wan 2020** | 139(137.2-141) | 136.5(133-138) |
| **Feng 2020** | 139(137-141) | Severe: 140(137-141); Critical: 140(137-142) |
| **Xiang 2020** | 136.8±2.9 | 134.5±5.0 |
| **Qian 2020** | 139.6(137.6-142) | 137.85(135.3-139.38) |
| **Serum creatinine, µmol/L** | | |
| **Mao 2020 A** | 65.6(39.4-229.1) | 71.6(35.9-9435.0) |
| **Qian 2020** | 66(57-76) | 81.5(70.75-90.5) |
| **Wan 2020** | 66(55-79) | 63.5(51.5-74.3) |
| **Zheng 2020** | 48.3(38.8, 57.9) | 47.5(39.2, 64.2) |
| **Lo 2020** | 66(15) | 58(16) |
| **Feng 2020** | 65.46(52.96-76.66) | Severe: 70.9(54.67-84.1); Critical: 67.95(55.23-81.28) |
| **Li 2020 C, M(QR)** | 66.5(21.7) | 69(26.5) |
| **Xiang 2020** | 68.7±25.1 | 69.6±16.6 |
| **Serum creatinine ≥133 µmol/L** | | |
| **Guan 2020** | 6/614(1.0%) | 6/138(4.3%) |
| **Wan 2020(＞97 µmol/L)** | 3(3%) | 3(7.5%) |
| **Zheng 2020(＞87 µmol/L)** | 1(0.8%) | 1(3.3%) |
| **Serum amyloid A, mg/L** | | |
| **Zhang 2020 B** | 91.5(24.9-163.2) | 108.4(54.1-161.6) |
| **Xiang 2020** | 86.76±75.58 | 191.90±96.15 |
| **Serum amyloid A> 10 mg/L** | | |
| **Zhang 2020 B** | 29/34(85.3%) | 17/17(100.0%) |
| **Creatine kinase, U/L** | | |
| **Mao 2020 A** | 59.0(19.0- 1260.0) | 83.0(8.8- 12216.0) |

| | | |
|---|---|---|
| **Zhang 2020 B** | 83.0(56.0-112.0) | 66.0(38.5-144.0) |
| **Wan 2020** | 57(36.5-86.5) | 82(56.3-146.2) |
| **Zheng 2020** | 68.7(43.2-111.3) | 100.3(61.3-398.6) |
| **Feng 2020** | 80(55-138) | Severe: 98(57-154); Critical: 93(52-246) |
| **Xiang 2020** | 129.2±138.2 | 182.7±134.1 |
| | **Creatine kinase＞200 U/L** | |
| **Guan 2020** | 67/536(12.5%) | 23/121(19.0%) |
| **Zhang 2020 B** | 1/35(2.8%) | 3/25(12.0%) |
| **Wan 2020** | 3(3%) | 7(17.5%) |
| **Zheng 2020(＞190U/L)** | 8(6.1%) | 9(30%) |
| | **CK-MB, μ/L** | |
| **Feng 2020** | 12.75(10.07-15.95) | Severe: 14.11(11.31-19.25); Critical: 15.5(11.75-23) |
| | **Lactate dehydrogenase, U/L** | |
| **Mao 2020 A** | 215.0(2.5-908.0) | 302.0(2.2-880.0) |
| **Wan 2020** | 212(179.5-259) | 309(253.8-408.3) |
| **Zheng 2020** | 162.0(133.7, 208.5) | 226.2(193.5, 315.1) - |
| **Lo 2020** | 183(61) | 238(52) |
| **Feng 2020** | 236(192-314) | Severe: 307(228-401); Critical: 378(275-523) |
| **Li 2020 C, M(QR)** | 231(54) | 368(55) |
| **Xiang 2020** | 237.5±72.6 | 467.3±144.3 |
| | **Lactate dehydrogenase ≥250 U/L** | |
| **Guan 2020** | 205/551(37.2%) | 72/124(58.1%) |
| **Li 2020 A(Increased)** | 8(50%) | 6(66.7%) |
| **Wan 2020(＞250 U/L)** | 28(29%) | 30(75%) |
| **Zheng 2020(＞225 U/L)** | 23(17.6%) | 15(50%) |
| | **Procalcitonin, ng/mL** | |
| **Zhang 2020 B** | 0.05(0.03-0.1) | 0.1(0.06-0.3) |
| **Qian 2020** | 0.03(0-0.04) | 0(0) |
| **Wan 2020** | 0.04(0.03-0.06) | 0.11(0.08-0.16) |
| **Li 2020 B** | 0.038(0.028-0.068) | 0.086(0.054-0.215) |
| **Feng 2020** | 0.04(0.02-0.06) | Severe: 0.06(0.02-0.13); Critical: 0.07(0-0.18) |

| | | |
|---|---|---|
| **Li 2020 C, M(QR)** | 0.02(0.02) | 0.03(0.03) |
| **Xiang 2020** | 0.04±0.03 | 0.16±0.15 |
| | **Procalcitonin ≥0.5 ng/mL** | |
| **Guan 2020** | 19/516(3.7%) | 16/117(13.7%) |
| **Zhang 2020 B(>0.1 ng/mL)** | 16/68(23.5%) | 25/50(50.0%) |
| **Wan 2020** | 0 | 1(2.5%) |
| **Li 2020 B(Increased)** | 23(39.7%) | 21(84%) |
| | **Blood urea nitrogen, mmol/L** | |
| **Mao 2020 A** | 3.8(1.6-13.7) | 4.6(1.5-48.1) |
| **Li 2020 C, M(QR)** | 4.7(1.5) | 4.8(2.1) |
| | **Urea, mmol/L** | |
| **Qian 2020** | 3.83(3.25-4.4) | 5.19(4.66-6.14) |
| | **C-reactive protein, mg/L** | |
| **Mao 2020 A** | 9.4(0.4-126.0) | 37.1(0.1-212.0) |
| **Zhang 2020 B** | 28.7(9.5-52.1) | 47.6(20.6-87.1) |
| **Qian 2020** | 5.98(1.4-11.3) | 30.63(12.5-103.4) |
| **Wan 2020** | 7.7(1.9-31.1) | 91(52.7-136.3) |
| **Li 2020 B** | 9.59(2.07-29.89) | 89.2(47.88-134.64) |
| **Zheng 2020** | 15.4(5.8-24.9) | 52.2(28.8-75.1) |
| **Lo 2020** | 1.65(3.16) | 2.63(3.12) |
| **Xiang 2020** | 16.05±20.26 | 79.86±56.87 |
| | **C-reactive protein ≥ 10 mg/L** | |
| **Guan 2020** | 371/658(56.4%) | 110/135(81.5%) |
| **Zhang 2020 B(>3mg/L)** | 72/81(88.9%) | 53/55(96.4) |
| **Xu 2020 B(Decreased)** | 18(36%) | 8(16%) |
| **Li 2020 A(Increased)** | 9(56.3%) | 7(77.8%) |
| **Li 2020 B(Increased)** | 27(46.6%) | 23(92%) |
| **Zheng 2020(＞8mg/L)** | 91(69.5%) | 30(100%) |
| | **Others** | |
| | **ESR, mm/h** | |
| **Li 2020 A(Increased)** | 3(18.8%) | 5(55.6%) |

| | | |
|---|---|---|
| **Feng 2020** | 48(27-83) | Severe: 45(33-79); Critical: 58(39-72) |
| **Xiang 2020** | 41.23±23.24 | 75.25±24.45 |
| | **BNP, pg/ml** | |
| **Feng 2020** | 34.53(21.15-67.1) | Severe: 52.5(16.93-113.3); Critical: 49.9(34.45-120.4) |
| **Li 2020 A(Increased)** | 1(6.3%) | 2(22.3%) |
| | **Urea, mmol/L** | |
| **Feng 2020** | 4.6(3.6-5.59) | Severe: 4.8(3.96-5.84); Critical: 5.65(4.3-7.73) |
| **Xiang 2020** | 4.19±1.34 | 5.56±2.15 |
| | **Ca2+, mmol/L** | |
| **Feng 2020** | 2.05(1.98-2.16) | Severe: 2.03(1.89-2.07); Critical: 1.95(1.87-2.06) |
| **Qian 2020** | 2.17(2.07-2.25) | 2.01(1.86-2.01) |
| **Xiang 2020** | 2.0±0.1 | 1.8±0.1 |
| | **Cl-, mmol/L** | |
| **Qian 2020** | 103.4(101.28-105.23) | 101.4(99.28-104.5) |
| **Xiang 2020** | 97.3±17.0 | 98.7±5.9 |
| | **Direct bilirubin, µmol/L** | |
| **Feng 2020** | 3.9(3-5.5) | Severe: 4.5(3.4-6.7); Critical: 4.1(3.1-5.5) |
| **Xiang 2020** | 2.48±0.98 | 4.42±1.91 |
| | **Others** | |
| **Li 2020 A(Ferritin, Increased)** | 5(31.3%) | 4(44.5%) |
| **Li 2020 A(cTnI, Increased)** | 0 | 2(22.3%) |
| **Li 2020 A(Transaminase, Increased)** | 8(50%) | 5(55.6%) |
| **Li 2020 A(Renal function damage)** | 1(6.3%) | 2(22.3%) |
| **Li 2020 A(Electrolyte disturbance)** | 7(43.8%) | 7(77.8%) |
| **Feng 2020(eGFR - ml/min/1.73m$^2$)** | 108(92-128) | Severe: 102(89-118); Critical: 96(76-120) |
| **Feng 2020(Methemoglobin, ng/ml)** | 11.7(3.65-40.2) | Severe: 28.04(10.07-51.5); Critical: 52.05(29.8-107.63) |
| **Feng 2020(Troponin increased)** | 59/296(19.9%) | 27/88(30.7%) |
| **Feng 2020(Lactic acid, mmol/L)** | 2.73(2.22-3.22) | Severe: 3.09(2.37-3.62); Critical: 2.5(2.15-3.34) |
| **Wang 2020 C(pH＞7.45)** | 17(40.5%) | 30(55.6%) |
| **Wang 2020 C(Partial pressure of oxygen＜80mmHg)** | 4(9.5%) | 43(79.6%) |

| | | |
|---|---|---|
| **Li 2020 C, M(QR)(IL-6, ng/L)** | 17.9(25.2) | 84(148) |
| **Xiang 2020(Hematocrit, L/L)** | 0.41±0.04 | 0.41±0.03 |
| **Xiang 2020(Globulin, g/L)** | 26.95±3.35 | 31.25±7.29 |
| **Xiang 2020(Alkaline phosphatase, U/L)** | 15.7±15.7 | 80.1±40.7 |
| **Xiang 2020(Uric acid, umol/L)** | 254.0±74.9 | 240.2±83.9 |
| **Xiang 2020(Triglyceride, mmol/L)** | 1.17±1.04 | 1.01±0.48 |
| **Xiang 2020(Total cholesterol, mmol/L)** | 3.72±0.76 | 3.57±0.64 |
| **Xiang 2020(High density lipoprotein, mmol/L)** | 1.08±0.31 | 0.95±0.29 |
| **Xiang 2020(Low density lipoprotein, mmol/L)** | 2.36±0.73 | 2.30±0.64 |
| **Xiang 2020(Amylase, U/L)** | 61.6±21.8 | 112.0±105.1 |
| **Zheng 2020(oxygenation index)** | 373(345,400) | 259(247,278) |
| | **Treatment** | |
| | **Oxygen therapy** | |
| **Guan 2020** | 331(35.7%) | 123(71.1%) |
| **Wan 2020** | 58(61%) | 32(80%) |
| **Feng 2020(Nasal cannula or no oxygen therapy)** | 352(100%) | 16(12.9%) |
| **Feng 2020(High-flow nasal cannula)** | 0 | 31(25%) |
| | **Invasive mechanical ventilation** | |
| **Guan 2020** | 0 | 25(14.5%) |
| **Wan 2020** | 0 | 1(2.5%) |
| **Feng 2020** | 0 | 39(31.5%) |
| | **Non-invasive mechanical ventilation** | |
| **Guan 2020** | 0 | 56(32.4%) |
| **Wan 2020(Non-invasive ventilation or high-flow nasal cannula)** | 7(7.4%) | 27(67.5%) |
| **Feng 2020** | 0 | 34(27.4%) |
| | **CRRT** | |
| **Guan 2020** | 0 | 9(5.2%) |
| **Wan 2020** | 1(1%) | 4(10%) |
| | **ECMO** | |
| **Guan 2020** | 0 | 5(2.9%) |
| **Wan 2020** | 0 | 0 |

| | | |
|---|---|---|
| **Feng 2020** | 0 | 4(3.2%) |
| | **Antibiotic treatment** | |
| **Guan 2020** | 498(53.8%) | 139(80.3%) |
| **Wan 2020** | 24(25%) | 35(87.5%) |
| **Feng 2020** | 209(59.4%) | 110(88.7%) |
| | **Antiviral treatment** | |
| **Guan 2020** | 313(33.8%) | 80(46.2%) |
| **Wan 2020** | 95(100%) | 40(100%) |
| **Feng 2020** | 199(56.5%) | 87(70.2%) |
| **Bin 2020** | 23(80%) | 8(88.9%) |
| | **Corticosteroid therapy** | |
| **Guan 2020** | 127(13.7%) | 77(44.5%) |
| **Wan 2020** | 15(15.8%) | 21(52.5%) |
| **Feng 2020** | 47(13.4%) | 80(64.5%) |
| | **Intravenous immunoglobulin therapy** | |
| **Guan 2020** | 86(9.3%) | 58(33.5%) |
| | **Antifungal treatment** | |
| **Guan 2020** | 18(1.9%) | 13(7.5%) |
| **Feng 2020** | 2(0.6%) | 6(4.8%) |
| | **Traditional Chinese Medicine** | |
| **Wan 2020** | 87(91.5%) | 37(92.5%) |
| **Bin 2020** | 34(75.6%) | 8(88.9%) |

Continuous data are median(IQR) or mean(SD); Categorical data are n(N%). NR: not reported. ICU: intensive unit care; COPD: chronic obstructive pulmonary disease; HIV: human immunodeficiency virus; ARDS, acute respiratory distress syndrome; CRPT: continuous renal replacement therapy; ECMO: extracorporeal membrane oxygenation.

| Table S39 Summary table of univariable risk factors significantly ($p < 0{\cdot}05$)associated with severe condition/ICU admission | |
|---|---|
| **Epidemiological characteristics and comorbidities** | |
| **Subject** | **Numbers** |
| Male | (4:0)/14 |
| Middle and old age | (6:0)/2 |
| Former smoker | (1:0)/2 |
| Never smoking | (0:2)/0 |
| Former epicenter exposure | (0:1)/7 |
| Medical staff | (0:1)/2 |
| Any comorbidity | (6:0)/3 |
| Hypertension | (7:0)/4 |
| Cardiovascular disease | (2:0)/5 |
| Cerebrovascular disease | (1:0)/2 |
| Cardio-cerebrovascular disease | (1:0)/2 |
| Diabetes | (5:0)/6 |
| COPD | (5:0)/2 |
| Malignancy | (1:0)/3 |
| Immunosuppression | (1:0)/1 |
| **Signs and symptoms on admission** | |
| Fever | (1:1)/15 |
| Highest temperature > 39℃ | (2:0)/2 |
| Cough | (4:2)/10 |
| Sputum production | (5:0)/4 |
| Dyspnea | (10:0)/3 |
| Respiratory rate >24/min | (1:0)/ |
| Fatigue | (2:0)/9 |
| Myalgia | (2:0)/7 |
| Hemoptysis | (1:0)/3 |
| Nausea | (0:1)/5 |

| | |
|---|---|
| Sore throat | (0:1)/6 |
| Diarrhea | (2:0)/7 |
| Anorexia | (2:1)/2 |
| Rash | (1:0)/0 |
| Conjunctival congestion | (1:0)/0 |
| Impaired consciousness | (1:0)/0 |
| Chest pain | (1:0)/3 |
| **Radiologic and laboratory findings on admission** | |
| Bilateral involvement | (7:0)/4 |
| Local patchy opacity | (1:0)/2 |
| Interstitial abnormalities | (1:0)/1 |
| ⩾4 lobs involved | (1:0)/0 |
| Pleural effusion | (3:0)/3 |
| Consolidation | (3:0)/3 |
| Bronchial Wall Thickening | (2:0)/1 |
| Interlobular septa thickening | (2:0)/0 |
| Paving Stone Sign | (1:0)/1 |
| Intrathoracic lymph node enlargement | (1:0)/3 |
| Upper lobes involvement | (1:0)/3 |
| Lower lobes involvement | (1:0)/3 |
| No abnormality | (0:1)/3 |
| WBC count > 10 × $10^9$/L | (2:0)/2 |
| WBC count > 9.5 × $10^9$/L | (1:0)/2 |
| WBC count < 4 × $10^9$/L | (1:0)/4 |
| WBC count < 3.5 × $10^9$/L | (1:0)/1 |
| Lymphocyte count < 1.5× $10^9$/L | (4:0)/2 |
| Lymphocyte count < 0.8× $10^9$/L | (1:0)/0 |
| Platelet count ＜150× $10^9$/L | (1:0)/0 |
| Platelet count ＜125× $10^9$/L | (1:0)/0 |
| Oxyhemoglobin saturation, % (Decreased) | (1:0)/0 |

| | |
|---|---|
| $PO_2$ < 80mmHg | (1:0)/0 |
| ALT >40U/L | (1:0)/1 |
| AST >40U/L | (3:0)/0 |
| D-dimer > 0.5mg/L | (2:0)/0 |
| D-dimer > 0.243mg/L | (1:0)/0 |
| Total bilirubin > 17.1 mmol/L | (1:0)/1 |
| Serum creatinine ⩾ 133 µmol/L | (1:0)/0 |
| Creatine kinase > 200 U/L | (2:0)/1 |
| Lactate dehydrogenase ⩾ 250 U/L | (2:0)/1 |
| Lactate dehydrogenase > 225 U/L | (1:0)/0 |
| Procalcitonin ⩾ 0.5 ng/mL | (2:0)/0 |
| Procalcitonin ⩾ 0.1 ng/mL | (1:0)/0 |
| C-reactive protein⩾10 mg/L | (2:0)/2 |
| **Univariable risk factors associated with ICU admission significantly (p＜0·05)** | |
| **Subject** | **Numbers** |
| Medical staff | (1:0)/0 |
| Hospitalized patients | (1:0)/0 |
| Any comorbidity | (1:0)/1 |
| Hypertension | (1:0)/1 |
| Cerebrovascular disease | (1:0)/0 |
| Diabetes | (1:0)/1 |
| Cough | (1:0)/1 |
| Dyspnea | (2:0)/1 |
| Sore throat | (1:0)/0 |
| Dizziness | (1:0)/0 |
| Respiratory rate >24/min | (1:0)/0 |
| WBC count > 10 × $10^9$/L | (1:0)/0 |
| Hypersensitive troponin I > 28 pg/mL (99th percentile) | (1:0)/0 |
| Procalcitonin ⩾ 0.5 ng/mL | (2:0)/0 |
| In "Numbers (*a*:*b*)/*c*" column, *a* is number of studies where the subject was estimated as risk factor significantly (p<0.05), *b* is number where the subject was estimated as | |

not risk factor (protective factor) significantly ($p<0.05$), and c is number of the other studies with no significant connection either ($p>0.05$). COPD: Chronic obstructive pulmonary disease; WBC: White blood cell; ALT: Alanine aminotransferase; AST: Aspartate aminotransferase.

#### Table S40 Univariable risk factors significantly (p＜0·05) associated with severe condition

| Study | OR | 95% CI | P value |
|---|---|---|---|
| Epidemiological characteristics and clinical outcomes | | | |
| Male | | | |
| Mao 2020 A | 1·93 | [1·11-3·37] | 0·02 |
| Feng 2020 | 1.61 | [1.05-2.46] | 0.03 |
| Li 2020 C | 3.43 | [1.05-11.22] | 0.042 |
| Wang 2020 C | 2.43 | [1.06-5.57] | 0.036 |
| Age ⩾ 50, y | | | |
| Guan 2020 | 1·99 | [1·41,2·79] | 0·0001 |
| Mao 2020 A | 2·93 | [1·63,5·27] | 0·0003 |
| Zhang 2020 B | 3·07 | [1·36,6·93] | 0·007 |
| ⩾45, Tian 2020 | 3·95 | [1·87,8·36] | 0·0003 |
| ⩾65, Feng 2020 | 2.18 | [1.39-2.41] | ＜0.001 |
| Wang 2020 C (＞60) | 5.15 | [2.11-12.53] | ＜0.001 |
| Former smoker | | | |
| Guan 2020 | 4·15 | [1·72,10·00] | 0·002 |
| Never smoking | | | |
| Guan 2020 | 0·53 | [0·35-0·80] | 0·003 |
| Li 2020 A | 0.11 | [0.02-0.83] | 0.032 |
| Former epicenter exposure | | | |
| Wan 2020 | 0.42 | [0.20-0.89] | 0.024 |
| Medical staff | | | |
| Li 2020 A | 0.10 | [0.02-0.51] | 0.006 |
| Comorbidity | | | |
| Any comorbidity | | | |
| Guan 2020 | 2·38 | [1·69-3·37] | ＜0·00001 |
| Mao 2020 A | 1·89 | [1·08-3·31] | 0·03 |
| Zhang 2020 B | 3·31 | [1·53-7·15] | 0·002 |

| | | | |
|---|---|---|---|
| Feng 2020 | 2.28 | [1.50-3.46] | < 0.001 |
| Li 2020 B | 10.61 | [2.93-38.40] | < 0.001 |
| Wan 2020 | 12.44 | [5.20-29.78] | < 0.001 |
| Hypertension | | | |
| Guan 2020 | 2·01 | [1·35-3·99] | 0·0006 |
| Mao 2020 A | 3·22 | [1·67-6·18] | 0·0005 |
| Feng 2020 | 1.89 | [1.20-2.97] | 0.006 |
| Li 2020 C | 8.84 | [1.01-77.40] | 0.049 |
| Wang 2020 C | 3.32 | [1.43-7.70] | 0.005 |
| Xiang 2020 | 15.20 | [2.19-105.42] | 0.006 |
| Zheng 2020 | 8.07 | [3.04-21.37] | < 0.001 |
| Cardiovascular disease | | | |
| Guan 2020 | 3·28 | [1·48-7·29] | 0·004 |
| Feng 2020 | 2.50 | [1.27-4.92] | 0.008 |
| Cerebrovascular disease | | | |
| Feng 2020 | 3.37 | [1.27-8.93] | 0.015 |
| Cardio cerebrovascular disease | | | |
| Wan 2020 | 16.59 | [1.93-142,84] | 0.011 |
| Diabetes | | | |
| Guan 2020 | 3·18 | [1·95-5·19 | < 0·00001 |
| Li 2020 B | 47.43 | [2.58-870.76] | 0.009 |
| Wan 2020 | 8.90 | [2.27-34.99] | 0.002 |
| Wang 2020 C | 3.26 | [1.16-9.11] | 0.025 |
| Xiang 2020 | 27 | [1.17-620.53] | 0.039 |
| COPD | | | |
| Guan 2020 | 5·51 | [1·76-17·29] | 0·003 |
| Feng 2020 | 5.47 | [2.24-13.39] | < 0.001 |
| Li 2020 A | 12 | [1.07-134.11] | 0.044 |
| Li 2020 B | 10.86 | [1.15-102.77] | 0.038 |
| Wan 2020 | 23.55 | [1.24-448.35] | 0.036 |
| Malignancy | | | |

| | | | |
|---|---|---|---|
| Feng 2020 | 4.15 | [1.29-13.33] | 0.017 |
| Immunosuppression | | | |
| Feng 2020 | 7.35 | [1.41-38.40] | 0.018 |
| Signs and symptoms on admission | | | |
| Fever | | | |
| Mao 2020 A | 0·31 | [0·17-0·55] | < 0·0001 |
| Feng 2020 | 6.12 | [2.17-17.23] | < 0.001 |
| Highest temperature > 39℃ during hospital admission | | | |
| Guan 2020 | 1·61 | [1·00-2·59] | 0·05 |
| Zheng 2020 | 11.85 | [1.56-90.12] | 0.17 |
| Cough | | | |
| Mao 2020 A | 0·33 | [0·19-0·58] | 0·0001 |
| Zhang 2020 B | 2·75 | [1·11,6·82] | 0·03 |
| Feng 2020 (Dry cough) | 0.64 | [0.44-0.93] | 0.019 |
| Li 2020 B | 9.95 | [1.24-79.55] | 0.03 |
| Wan 2020 | 2.93 | [1.04-8.24] | 0.042 |
| Wang 2020 C | 2.71 | [1.07-6.84] | 0.035 |
| Sputum production | | | |
| Feng 2020 | 2.57 | [1.67-3.96] | < 0.001 |
| Li 2020 C | 3.66 | [1.05-12.79] | 0.042 |
| Li 2020 B | 4.88 | [1.51-15.79] | 0.008 |
| Wan 2020 | 3.82 | [1.13-12.87] | 0.031 |
| Xiang 2020 | 9.87 | [1.69-57.60] | 0.011 |
| Dyspnea | | | |
| Guan 2020 | 3·38 | [2·37-4·83] | < 0·00001 |
| Xu 2020 B (chest tightness or dyspnea) | 35·53 | [1·76,718·74] | 0·02 |
| Tian 2020 | 34·35 | [9·40,125·50] | < 0·00001 |
| Feng 2020 | 6.35 | [3.94-10.23] | < 0.001 |
| Lv 2020 | 36 | [1.71-757.79] | 0.021 |
| Li 2020 B | 10.89 | [2.07-57.20] | 0.005 |
| Tang 2020 | 9.93 | [1.90-51.72] | 0.006 |

| | | | |
|---|---|---|---|
| Tang 2020 | 8.58 | [2.36-31.16] | 0.001 |
| Wan 2020 | 157.04 | [9.12-2704.97] | < 0.001 |
| Zheng 2020 | 3.58 | [1.37-9.33] | 0.009 |
| | Respiratory rate > 24/min | | |
| Wan 2020 | 8.90 | [2.27-34.99] | 0.002 |
| | Fatigue | | |
| Xu 2020 B | 15 | [2·49,90·23] | 0·003 |
| Li 2020 C | 14 | [3.18-61.60] | < 0.001 |
| | Myalgia | | |
| Mao 2020 A A* | 4·79 | [1·80-12·71] | 0·002 |
| Wan 2020 | 2.53 | [1.17-5.47] | 0.018 |
| | Hemoptysis | | |
| Guan 2020 | 3·63 | [1·01-13·00] | 0·05 |
| | Nausea | | |
| Zhang 2020 B | 0·32 | [0·11,0·91] | 0·03 |
| | Sore throat | | |
| Wan 2020 | 0.04 | [0.002-0.61] | 0.021 |
| | Diarrhea | | |
| Li 2020 C | 226.11 | [11.48-4454.26] | < 0.001 |
| Wan 2020 | 8.67 | [2.83-26.50] | < 0.001 |
| | Anorexia | | |
| Mao 2020 A | 0·53 | [0·29-0·97] | 0·04 |
| Liu 2020 C | 5.13 | [1.70-15.43] | 0.004 |
| Wan 2020 | 35.99 | [1.97-655.75] | 0.016 |
| | Rash | | |
| Guan 2020 | 27·01 | [1·29-565·10] | 0·03 |
| | Conjunctival congestion | | |
| Guan 2020 | 4·36 | [1·16,16·40] | 0·03 |
| | Impaired consciousness | | |
| Mao 2020 A | 7·11 | [1·96-25·76] | 0·003 |
| | Chest pain | | |

| | | | |
|---|---|---|---|
| Li 2020 B | 10.86 | [1.14-102.77] | 0.038 |
| Radiologic and laboratory and findings | | | |
| Bilateral involvement of chest CT on admission | | | |
| Guan 2020 | 5·46 | [3·59,8·30] | < 0·0001 |
| Xu 2020 B (Bilateral upper lobes) | 17·60 | [2·06,150·03] | 0·009 |
| Xu 2020 B (Bilateral lower lobes) | 55·08 | [3·02,1003·70] | 0·007 |
| Feng 2020 | 3.07 | [1.42-6.63] | 0.004 |
| Li 2020 C | 113.67 | [6.07-2127.89] | 0.002 |
| Liu 2020 C | 28.78 | [1.65-501.60] | 0.021 |
| Zheng 2020 | 7.02 | [2.32-21.23] | < 0.001 |
| Local patchy shadowing of chest CT on admission | | | |
| Guan 2020 | 1·90 | [1·36,2·66] | 0·0002 |
| Interstitial abnormalities of chest CT on admission | | | |
| Guan 2020 | 2·56 | [1·71,3·83] | < 0·0001 |
| Number of lobs involved of chest CT on admission | | | |
| 4-5: Xu 2020 B | 14·12 | [1·66,119·11] | 0·02 |
| Pleural Effusion | | | |
| Feng 2020 | 4.76 | [2.07-10.92] | < 0.001 |
| Li 2020 B | 47.43 | [2.58-870.76] | 0.009 |
| Zhao 2020 | 4.81 | [1.32-17.54] | 0.017 |
| Consolidation | | | |
| Liu 2020 C | 51 | [2.79-932.53] | 0.008 |
| Li 2020 B | 6.39 | [1.72-23.72] | 0.006 |
| Xu 2020 B | 11.63 | [2.68-50.40] | 0.001 |
| Bronchial Wall Thickening | | | |
| Liu 2020 C | 57.07 | [10.78-302.11] | < 0.001 |
| Li 2020 B | 32.59 | [7.88-134.88] | < 0.001 |
| Interlobular septa thickening | | | |
| Tang 2020 | 3.58 | [1.00-12.79] | 0.049 |
| Xu 2020 B | 0.15 | [0.06-0.39] | < 0.001 |
| Paving Stone Sign | | | |

| | | | |
|---|---|---|---|
| Li 2020 B | 3.34 | [1.26-8.88] | 0.016 |
| Intrathoracic lymph node enlargement | | | |
| Li 2020 B | 47.43 | [2.58-870.76] | 0.009 |
| Upper lobes involvement | | | |
| Xu 2020 B | 17.60 | [2.06-150.03] | 0.009 |
| Lower lobes involvement | | | |
| Xu 2020 B | 55.08 | [3.02-1003.70] | 0.007 |
| No radiologic abnormality of chest CT on admission | | | |
| Guan 2020 | 0·31 | [0·15,0·62] | 0·001 |
| White blood cell count > 10 × $10^9$/L | | | |
| Guan 2020 | 2·54 | [1·43-4·52] | 0·002 |
| Zhang 2020 B ( > 9.5 × $10^9$/L) | 5·90 | [1·81,19·20] | 0·003 |
| Feng 2020 | 3.78 | [2.07-6.93] | < 0.001 |
| White blood cell count < 4× $10^9$/L | | | |
| Guan 2020 | 4·01 | [2·84-5·68] | < 0·00001 |
| Wan 2020 ( < 3.5× $10^9$/L) | 34.13 | [10.44-111.54] | < 0·001 |
| Lymphocyte count < 1.5× $10^9$/L | | | |
| Guan 2020 | 5·47 | [2·51-11·91] | < 0·0001 |
| Feng 2020 | 4.04 | [2.59-6.31] | < 0.001 |
| Li 2020 B | 8.67 | [2.28-32.87] | 0.002 |
| Wan 2020 | 6.56 | [2.72-15.78] | < 0.001 |
| Zheng 2020 ( < 0.8× $10^9$/L) | 2.69 | [1.17-6.18] | 0.02 |
| Platelet count < 150× $10^9$/L | | | |
| Guan 2020 | 2·96 | [2·07-4·22] | < 0·00001 |
| Wan 2020 ( < 125×$10^9$/L) | 3.27 | [1.30-8.24] | 0.012 |
| Oxyhemoglobin saturation, % | | | |
| Li 2020 B (Decreased) | 8.33 | [2.48-27.93] | < 0.001 |
| $PO_2$ < 80mmHg | | | |
| Wang 2020 C | 37.14 | [10.91-126.39] | < 0.001 |
| Alanine aminotransferase > 40 U/L | | | |
| Guan 2020 | 1·59 | [1·04-2·43] | 0·03 |

| | | | |
|---|---|---|---|
| Aspartate aminotransferase > 40 U/L | | | |
| Guan 2020 | 2·92 | [1·97-4·34] | < 0·00001 |
| Wan 2020 | 3.20 | [1.37-7.45] | 0.007 |
| Zheng 2020 | 8.07 | [3.04-21.37] | < 0.001 |
| D-dimer > 0.5mg/L | | | |
| Guan 2020 | 1·94 | [1·27-2·97] | 0·002 |
| Zhang 2020 B ( > 0.243mg/L) | 3·96 | [1·56,10·05] | 0·004 |
| Wang 2020 C (Increased) | 3.64 | [1.54-8.61] | 0.003 |
| Total bilirubin > 17.1 mmol/L | | | |
| Guan 2020 | 1·39 | [0·78-2·47] | 0·26 |
| Serum creatinine ⩾ 133 μmol/L | | | |
| Guan 2020 | 4·61 | [1·46-14·51] | 0·009 |
| Creatine kinase > 200 U/L | | | |
| Wan 2020 | 6.51 | [1.59-26.64] | 0.009 |
| Zheng 2020 | 6.59 | [2.29-19.00] | < 0.001 |
| Lactate dehydrogenase ⩾ 250 U/L | | | |
| Guan 2020 | 2·34 | [1·57-3·47] | < 0·0001 |
| Wan 2020 ( > 250U/L) | 7.18 | [3.10-16.64] | < 0.001 |
| Zheng 2020 ( > 225 U/L) | 4.70 | [2.02-10.94] | < 0.001 |
| Procalcitonin ⩾ 0.5 ng/mL | | | |
| Guan 2020 | 4·14 | [2·06-8·33] | < 0·0001 |
| Zhang 2020 B ( > 0.1 ng/mL) | 3·25 | [1·48,7·15] | 0·003 |
| Li 2020 B | 7.99 | [2.43-26.30] | < 0.001 |
| C-reactive protein⩾10 mg/L | | | |
| Guan 2020 | 3·55 | [2·22-5·66] | < 0·00001 |
| Li 2020 B | 13.20 | [2.85-61.24] | 0.001 |

OR=odds ratio. COPD=chronic obstructive pulmonary disease.

#### Table S41 Epidemiological and clinical characteristics of the confirmed patients with ICU admission or not

| | Non-ICU admission | | ICU admission | |
|---|---|---|---|---|
| | **Huang 2020 A** | **Wang 2020 A** | **Huang 2020 A** | **Wang 2020 A** |
| **Epidemiological characteristics** | | | | |
| **Total** | 28/41 (68.3%) | 102/138 (73.9%) | 13/41 (31.7%) | 36/138 (26.1%) |
| **Male** | 19 (68%) | 53 (52.0%) | 11 (85%) | 22 (61.1%) |
| **Female** | 9 (32%) | 49 (48.0%) | 2 (15%) | 14 (38.9%) |
| **Age Median (IQR)** | 49 (41-57.5) | 51 (37-62) | 49 (41-61) | 66 (57-78) |
| **Huanan seafood market exposure** | 18 (64%) | 7 (6.9%) | 9 (69%) | 5 (13.9%) |
| **Current smoker** | 3 (11%) | NR | 0 | NR |
| **Time from onset of symptoms to first hospital admission** | NR | 6.0 (3.0-7.0) | NR | 8.0 (4.5-10.0) |
| **Medical staff** | NR | 39 (38.2%) | NR | 1 (2.8%) |
| **Hospitalized patients** | NR | 8 (7.8%) | NR | 9 (25.0%) |
| **Clinical outcomes** | | | | |
| **Discharge** | 21 (75%) | 38 (37.3%) | 7 (54%) | 9 (25%) |
| **Staying in hospital** | 6 (21%) | 74 (72.5%) | 0 (transferred to the general wards)<br>1 (8%) (still in ICU) | 10 (27.8%) (transferred to the general wards)<br>11 (30.6%) (still in ICU) |
| **Death** | 1 (4%) | 0 | 5 (38%) | 6 (16.7%) |
| **Comorbidity** | | | | |
| **Any comorbidity** | 8 (29%) | 38 (37.3) | 5 (38%) | 26 (72.2%) |
| **Hypertension** | 4 (14%) | 22 (21.6%) | 2 (15%) | 21 (58.3%) |
| **Diabetes** | 7 (25%) | 6 (5.9%) | 1 (8%) | 8 (22.2%) |
| **Cardiovascular disease** | 3 (11%) | 11 (10.8%) | 3 (23%) | 9 (25.0%) |
| **Cerebrovascular disease** | NR | 1 (1.0%) | NR | 6 (16.7%) |
| **COPD** | 0 | 1 (1.0%) | 1 (8%) | 3 (8.3%) |
| **Malignancy** | 1 (4%) | 6 (5.9%) | 0 | 4 (11.1%) |

| | | | | |
|---|---|---|---|---|
| **Chronic liver disease** | 1 (4%) | 4 (3.9%) | 0 | 0 |
| **Chronic kidney disease** | NR | 2 (2.0%) | NR | 2 (5.6%) |
| **Immunodeficiency/HIV** | NR | 2 (2.0%) | NR | 0 |
| | | **Signs and symptoms on admission** | | |
| **Fever** | 27 (96%) | 100 (98.0%) | 13 (100%) | 36 (100%) |
| **Highest temperature＞39℃** | 11 (39%) | NR | 3 (23%) | NR |
| **Highest temperature＜37.5℃** | 1 (4%) | NR | 0 | NR |
| **Cough** | 11 (39%) (dry cough mostly) | 61 (59.8%) (dry cough) | 11 (85%) (dry cough mostly) | 21 (58.3%) (dry cough) |
| **Sputum production** | 11 (39%) | 29 (28.4%) | 5 (38%) | 8 (22.2%) |
| **Dyspnea** | 10/27 (37%) | 20 (19.6%) | 12 (92%) | 23 (63.9%) |
| **Fatigue** | 11 (39%) (fatigue or myalgia) | 67 (65.7%) | 7 (54%) (fatigue or myalgia) | 29 (80.6%) |
| **Myalgia** | | 36 (35.3%) | | 12 (33.3%) |
| **Headache** | 3/25 (12%) | 6 (5.9%) | 0 | 3 (8.3%) |
| **Hemoptysis** | 1/26 (4%) | NR | 1 (8%) | NR |
| **Diarrhea** | 1/25 (4%) | 8 (7.8%) | 0 | 6 (16.7%) |
| **Sore throat** | NR | 12 (11.8%) | NR | 12 (33.3%) |
| **Nausea or vomiting** | NR | Nausea: 10 (9.8%); Vomiting: 2 (2.0%) | NR | Nausea: 4 (11.1%), Vomiting: 3 (8.3%) |
| **Dizziness** | NR | 5 (4.9%) | NR | 8 (22.2%) |
| **Anorexia** | NR | 31 (30.4%) | NR | 24 (66.7%) |
| **Abdominal pain** | NR | 0 | NR | 3 (8.3%) |
| **Time from onset of symptoms to shortness of breath** | NR | 2.5 (0.0-7.3) | NR | 6.5 (3.0-10.8) |
| **Time from onset of symptoms to shortness of ARDS** | NR | 8.0 (6.3-11.3) | NR | 8.0 (6.0-12.0) |
| **Respiratory rate** | 4 (14%) > 24/min | 20 (19-21) | 8 (62%) > 24/min | 20 (16-25) |
| | | **Complications** | | |
| **ARDS** | 1 (4%) | 5 (4.9%) | 11 (85%) | 22 (61.1%) |
| **Acute cardiac injury** | 1 (4%) | 2 (2.0%) | 4 (31%) | 8 (22.2%) |
| **Acute kidney injury** | 0 | 2 (2.0%) | 3 (23%) | 3 (8.3%) |
| **Shock** | 0 | 1 (1.0%) | 3 (23%) | 11 (30.6%) |

| | | | | |
|---|---|---|---|---|
| **Arrhythmia** | NR | 7 (6.9%) | NR | 16 (44.4%) |
| **RNAaemia** | 4 (14%) | NR | 2 (15%) | NR |
| **Secondary infection** | 0 | NR | 4 (31%) | NR |
| | | **Radiologic findings on admission** | | |
| **Bilateral involvement of chest CT on admission** | 27 (96%) | 102 (100%) | 13 (100%) | 36 (100%) |
| **Local patchy shadowing of chest CT on admission** | NR | 0 | NR | 0 |
| **Ground glass opacity of chest CT on admission** | NR (bilateral ground-glass opacities on chest CT scans for most patients) | 102 (100%) | NR (bilateral ground-glass opacities on chest CT scans for most patients) | 36 (100) |
| | | **Laboratory and findings on admission** | | |
| **White blood cell count, × $10^9$/L** | 5.7 (3.1–7.6) | 4.3 (3.3-5.4) | 11.3 (5.8–12.1) | 6.6 (3.6-9.8) |
| **White blood cell count > 10 × $10^9$/L** | 5/27 (19%) | NR | 7 (54%) | NR |
| **White blood cell count < 4× $10^9$/L** | 9/27 (33%) | NR | 1 (8%) | NR |
| **Neutrophil count, × $10^9$/L** | 4.4 (2.0–6.1) | 2.7 (1.9-3.9) | 10.6 (5.0–11.8) | 4.6 (2.6-7.9) |
| **Monocyte count, $10^9$/L** | NR | 0.4 (0.3-0.5) | NR | 0.4 (0.3-0.5) |
| **Lymphocyte count, × $10^9$/L** | 1.0 (0.7–1.1)<br>< 1× $10^9$/L: 15 (54%) | 0.9 (0.6-1.2) | 0.4 (0.2–0.8)<br>< 1× $10^9$/L: 11 (85%) | 0.8 (0.5-0.9) |
| **Hemoglobin, g/L** | 130.5 (120.0–140.0) | NR | 122.0 (111.0–128.0) | NR |
| **Platelet count, × $10^9$/L** | 149.0 (131.0–263.0)<br>< 100× $10^9$/L: 1/27 (4%)<br>≥100× $10^9$/L: 26/27 (96%) | 165 (125-188) | 196.0 (165.0–263.0)<br>< 100× $10^9$/L: 1 (8%)<br>≥100× $10^9$/L: 12 (92%) | 142 (119-202) |
| **Prothrombin time, s** | 10.7 (9.8–12.1) | 12.9 (12.3-13.4) | 12.2 (11.2–13.4) | 13.2 (12.3-14.5) |
| **Activated partial thromboplastin time, s** | 27.7 (24.8–34.1) | 31.7 (29.6-33.5) | 26.2 (22.5–33.9) | 30.4 (28.0-33.5) |
| **D-dimer, mg/L** | 0.5 (0.3–0.8) | 166 (101-285) | 2.4 (0.6–14.4) | 414 (191-1324) |
| **Albumin, g/L** | 34.7 (30.2–36.5) | NR | 27.9 (26.3–30.9) | NR |
| **Alanine aminotransferase, U/L** | 27.0 (19.5–40.0) | 23 (15-36) | 49.0 (29.0–115.0) | 35 (19-57) |

| | | | | |
|---|---|---|---|---|
| **Aspartate aminotransferase, U/L** | 34.0 (24.0–40.5) | 29 (21-38) | 44.0 (30.0–70.0) | 52 (30-70) |
| | > 40 U/L: 7 (25%) | | > 40 U/L: 8 (62%) | |
| | ⩽40 U/L: 21 (75%) | | ⩽40 U/L: 5 (38%) | |
| **Total bilirubin, mmol/L** | 10.8 (9.4–12.3) | 9.3 (8.2-12.8) | 14.0 (11.9–32.9) | 11.5 (9.6-18.6) |
| **Potassium, mmol/L** | 4.1 (3.8–4.6) | NR | 4.6 (4.0–5.0) | NR |
| **Sodium, mmol/L** | 139.0 (137.5–140.5) | NR | 138.0 (137.0–139.0) | NR |
| **Serum creatinine, μmol/L** | 73.3 (57.5–84.7) | 71 (58-84) | 79.0 (53.1–92.7) | 80 (66-106) |
| | > 133 μmol/L: 2 (7%) | | > 133 μmol/L: 2 (15%) | |
| **Creatine kinase–MB, U/L** | NR | 13 (10-14) | NR | 18 (12-35) |
| **Creatine kinase, U/L** | 133.0 (61.0–189.0) | 87 (54-121) | 132.0 (82.0–493.0) | 102 (62-252) |
| | > 185 U/L 7/27 (26%) | | > 185 U/L: 6 (46%) | |
| | ⩽185 U/L 20/27 (74%) | | ⩽185 U/L: 7 (54%) | |
| **Lactate dehydrogenase, U/L** | 281.0 (233.0–357.0) | 212 (171-291) | 400.0 (323.0–578.0) | 435 (302-596) |
| | > 245 U/L: 17/27 (63%) | | > 245 U/L: 12 (92%) | |
| | ⩽ 245 U/L: 10/27 (37%) | | ⩽ 245 U/L: 1 (8%) | |
| **Hypersensitive troponin I, pg/mL** | 3.5 (0.7–5.4) | 5.1 (2.1-9.8) | 3.3 (3.0–163.0) | 11.0 (5.6-26.4) |
| | > 28 pg/mL (99th percentile): 1 (4%) | | > 28 pg/mL (99th percentile): 4 (31%) | |
| **Procalcitonin, ng/mL** | 0.1 (0.1–0.1) | NR | 0.1 (0.1–0.4) | NR |
| | ⩾0.5 ng/mL: 0/27 | ⩾0.5 ng/mL: 22 (21.6%) | ⩾0.5 ng/mL: 3/12 (25%) | ⩾0.5 ng/mL: 27 (75.0%) |
| | ⩾0.1 to <0.5 ng/mL: 6/27 (22%) | | ⩾0.1 to <0.5 ng/mL: 3/12 (25%) | |
| **Blood urea nitrogen, mmol/L** | NR | 4.0 (3.1-5.1) | NR | 5.9 (4.3-9.6) |
| | | **Treatment** | | |
| **Oxygen therapy** | 26 (93%) | 102 (100%) | 1 (8%) | 4 (11.11%) |
| **Invasive mechanical ventilation** | 0 | 0 | 2 (15%) | 17 (47.22%) |
| **Non-invasive mechanical ventilation** | 2 (7%) | 0 | 8 (62%) | 15 (41.7%) |
| **CRRT** | 0 | 0 | 3 (23%) | 2 (5.56%) |
| **ECMO** | 2 (7%) | 0 | 2 (15%) | 4 (11.1%) |
| **Antibiotic treatment** | 28 (100%) | NA | 13 (100%) | NA |

| | | | | |
|---|---|---|---|---|
| **Antiviral treatment** | 26 (93%) | 90 (88.2%) | 12 (92%) | 34 (94.4%) |
| **Corticosteroid therapy** | 3 (11%) | 36 (35.3%) | 6 (46%) | 26 (72.2%) |

Continuous data are median (IQR) or mean (SD); Categorical data are n (N%). NR: not reported. ICU: intensive unit care; COPD: chronic obstructive pulmonary disease; HIV: human immunodeficiency virus; ARDS, acute respiratory distress syndrome; CRPT: continuous renal replacement therapy; ECMO: extracorporeal membrane oxygenation.

**Table S42 Univariable risk factors significantly ($p < 0·05$)associated with ICU admission**

| Study | OR | 95% CI | P value |
|---|---|---|---|
| **Epidemiological characteristics and clinical outcomes** | | | |
| **Medical staff** | | | |
| **Wang 2020 A** | 0·05 | [0·01-0·35] | 0·003 |
| **Hospitalized patients** | | | |
| **Wang 2020 A** | 3·92 | [1·38-11·13] | 0·01 |
| **Comorbidity** | | | |
| **Any comorbidity** | | | |
| **Wang 2020 A** | 4·38 | [1·90-10·07] | 0·0005 |
| **Hypertension** | | | |
| **Wang 2020 A** | 5·09 | [2·26-11·48] | <0·0001 |
| **Cerebrovascular disease** | | | |
| **Wang 2020 A** | 3·74 | [2·42-5·79] | <0·00001 |
| **Diabetes** | | | |
| **Wang 2020 A** | 4·57 | [1·46-14·28] | 0·0009 |
| **Signs and symptoms on admission** | | | |
| **Cough** | | | |
| **Huang 2020 A** | 8·50 | [1·57-45·92] | 0·01 |
| **Dyspnea** | | | |
| **Huang 2020 A** | 20·40 | [2·30-181·26] | 0·007 |
| **Wang 2020 A** | 7·25 | [3·14-16·76] | <0·00001 |
| **Sore throat** | | | |
| **Wang 2020 A** | 3·75 | [1·50-9·39] | 0·005 |
| **Dizziness** | | | |
| **Wang 2020 A** | 5·54 | [1·68-18·29] | 0·005 |
| **Respiratory rate >24 breaths/ min** | | | |
| **Huang 2020 A** | 9·60 | [2·06-44·74] | 0·004 |
| **Radiologic and laboratory and findings** | | | |
| **White blood cell count $> 10 \times 10^9$/L** | | | |

| | | | |
|---|---|---|---|
| **Huang 2020 A** | 5·13 | [1·19-22·11] | 0·03 |
| **Hypersensitive troponin I ＞28 pg/mL (99th percentile)** | | | |
| **Huang 2020 A** | 12·00 | [1·18-121·81] | 0·04 |
| **Procalcitonin ≥ 0.5 ng/mL** | | | |
| **Huang 2020 A** | 20·26 | [0·96-429·41] | 0·05 |
| **Wang 2020 A** | 8·93 | [3·83-20·78] | <0·00001 |

OR=odds ratio.
